# Genetic modifiers of psychiatric, motor, and cognitive symptoms in Huntington’s disease

**DOI:** 10.64898/2026.06.16.26355789

**Authors:** Qingqin S Li, Regeneron Genetic Center, Seung Kwak, Cristina Sampaio, Thomas F Vogt, Jim Rosinski

## Abstract

The Enroll-HD natural history platform provides rich longitudinal phenotypes enabling genome-wide analyses across diverse clinical domains. Psychiatric symptoms are a major source of morbidity in Huntington’s disease (HD), yet the genetic architecture underlying their onset is poorly understood. We analyzed ∼18,000 people with HD (PwHD) to define genetic determinants of ages at psychiatric, motor, and cognitive symptom onset, and HD diagnosis. GWAS meta-analysis recapitulated 11 established modifiers of motor onset and identified a novel locus spanning *RAB3B*/*ZFYVE9* associated with age at violent/aggressive behavior onset. Exome-wide analyses in Enroll-HD participants implicated rare variants in *FAN1*, *PMS1*, *POLD1*, and *HTT*. Several HD modifiers of motor and cognitive symptom onset (*MSH3*, *FAN1*, *HTT*) also influenced psychiatric symptom onset, whereas *PMS1* and *POLD1* showed significant association with motor symptom onset. Psychiatric polygenic scores predicted psychiatric symptom onset, revealing a hybrid architecture combining psychiatric liability in general population with HD- or repeat expansion disease (RED)-specific pathways.

## Introduction

Psychiatric symptoms and behavioral disturbances, core clinical features of Huntington’s disease (HD), are among the most burdensome manifestations of HD, that can emerge years before motor diagnosis and contribute substantially to functional decline, caregiver burden, and reduced quality of life. Large observational cohorts show that depression, irritability, anxiety, apathy, and obsessive - compulsive behaviors occur at markedly elevated rates in people with HD (PwHD), that is individuals with an expanded CAG repeat in their huntingtin (*HTT*) gene^1,2^, though conflicting evidence exists^3^. Several symptoms including apathy and obsessive-compulsive behaviors increase in frequency and severity with disease progression^1,2,4^. Longitudinal analyses further demonstrate that apathy and related motivational deficits worsen over time independent of depression, underscoring their role as intrinsic manifestations of HD neuropathology rather than psychosocial sequelae^4^. Collectively, these findings indicate that psychiatric symptoms represent early, prevalent, and clinically meaningful components of HD, with trajectories that may reflect underlying genetic and neurobiological modifiers of disease manifestation. Despite their clinical importance, the biological mechanisms that shape the timing and manifestation of psychiatric symptoms in HD remain poorly understood.

Previous common variant genetic studies in HD have focused primarily on modifiers of motor onset and time to various clinical landmarks in motor, cognitive, and functional domains, identifying loci - most prominently genes involved in DNA repair and handling, and specifically the DNA mismatch repair (MMR) pathway such as *PMS1, MLH1*, *MSH3*, *PMS2*, *MLH3*, *FAN1*, *LIG1*, and *POLD1. C*is-variants in *HTT* (CAACAG-dup, CAA/CCA-loss, CAA-loss, and CCA-loss), and other non-MMR genes, such as *TCERG1, RRM2B, CCDC82, MED15,* and *CCDC134* /*SREBF2* /*SHISA8* /*TNFRSF13C* /*CENPM* /*SMIM45* /*SEPTIN3*^5^, are also implicated. The MMR genes (such as *MSH3*, *PMS2*, *MLH3*, and *FAN1)* influence disease course through effects on somatic instability (SI) of the expanded CAG repeat, while additional genes such as *MSH2 /MSH6, ATAD5* /*ADAP2* /*TEFM,* or variants in 5’ UTR from *HTT* are associated with SI in blood, but have not yet been associated with time to reach clinical landmarks^5^. Previous whole exome sequencing study has additionally implicated the rare variants from *FAN1* and their biological link to SI and HD progression^6^. Whether these same pathways contribute to psychiatric symptom onset is not fully understood, although it is reasonable to hypothesize that SI is required for pathology in vulnerable cell types including those involved in manifesting psychiatric symptoms. For mild cognitive impairment and dementia, previous GWAS has linked Alzheimer’s disease (AD) disease susceptibility gene *APOE* to neuropsychiatric symptoms^7^. At the same time, psychiatric symptoms in HD resemble those seen in primary psychiatric disorders, although this is usually manifested as subthreshold symptoms and not a full-blown episode. Polygenic scores (PGS) of psychiatric disorders have been associated with the presence of psychiatric symptoms in HD, including depression and psychosis^8^. Further study is needed to systematically evaluate these two hypotheses.

As a monogenic neurodegenerative disorder with a known causal pathogenic trinucleotide repeat expansion, HD offers a rare opportunity to examine how general psychiatric risk interacts with disease-specific molecular pathology within individuals. Deeply phenotyped large cohorts such as Enroll-HD and REGISTRY^9,10^ enable longitudinal assessment of motor, cognitive, and six psychiatric symptom domains, allowing comparison of genetic association across phenotypes and examination of potentially shared versus domain-preferential genetic effects. In this study, we aim to investigate the genetic contributions of both common and rare variants in HD to age at psychiatric, motor, and cognitive symptom onset using genome-wide association study (GWAS), PGS association analysis, and exome-wide association study (ExWAS).

## Results

Onset of motor symptoms was reported in ∼72-83% of study participants, while emergence of cognitive impairment was reported in ∼40-53% of study participants. In addition, onset of apathy, depression, irritability, perseverative obsessive behaviors, violent or aggressive behaviors, and psychosis was reported in ∼ 47-60%, ∼55-72%, ∼51-67%, ∼32-53%, ∼29-39%, and ∼11% of participants, respectively (**Supplementary Table 1**). The domain associated with initial symptom onset was also reported by the participant, family, and rater (**Supplementary Note, Supplementary Table 2)**.

### GWAS Results

Meta-analyses of three non-overlapping cohorts (by assay and study) identified a total of 35 independent genome wide significant association signals (r^2^<0.1) across 12 genomic regions and 11 traits (**Figures 1 and 2**, **Table 1**, **Supplementary Tables 3-6, Supplementary Figure 1**). The analyses implicated known HD modifiers identified by the GeM-HD consortium^5,11^: *HTT*, MMR genes (*PMS1*, *MLH1*, *MSH3*, *PMS2*, *FAN1*, *LIG1*, *POLD1*) and non-MMR genes (*TCERG1* and *RRM2B*) and one additional novel locus in the chromosome 1 genomic region spanning *RAB3B and ZFYVE9* (**Supplementary Figure 2**). Most of the association signals were observed in time to motor symptom onset and time to age at HD diagnosis, while subsets of those signals were also detected by the psychiatric and cognitive phenotypes. The novel locus near *RAB3B/ZFYVE9* was identified from estimated age of onset of violent or aggressive behavior symptoms. *HTT*, *MSH3*, and *FAN1* were associated with estimated age of onset of cognitive and apathy symptoms. *HTT* was associated with estimated age of onset of irritability, and *MSH3* was associated with estimated age of onset of irritability, perseverative obsessive behaviour, and violent or aggressive behavior symptoms. The association signals for previously reported modifiers are also reported (**Supplementary Table 7** and **Supplementary Figure 3**). Among the known modifier genes, there were a few novel independent signals for *MSH3* and *FAN1,* respectively (**Supplementary Table 4** and **Supplementary Figure 4 and 5**). *MSH3* variants in LD with 5AM1 or 5AM3 and a single *FAN1* variant in LD with 15AM2 reached genome wide significance threshold and were located in *MSH3* and *FAN1* super-enhancer regions (personal communication with Matthew Baffuto and Nathaniel Heintz, **Supplementary Table 8**)^12^, suggesting a potential regulatory mechanism for functional follow up. 5:80789582:G:A, 15:30688680:G:GT, and 15:30700921:A:G appeared to be independent signals not in strong LD with other lead variants previously reported. For the novel signals from chromosome 1 for onset of violent or aggressive behavior, both lead variants were in the introns of *RAB3B* or *ZFYVE9,* respectively (**Supplementary Figure 2**). In the genomic region between *RAB3B* and *ZFYVE9,* there are other variants in LD with the lead variants in genes such as basic transcription factor 3 like 4 (*BTF3L4*), thioredoxin domain containing 12 (*TXNDC12*), and KTI12 chromatin associated homolog (*KTI12*).

**Figure 1.**
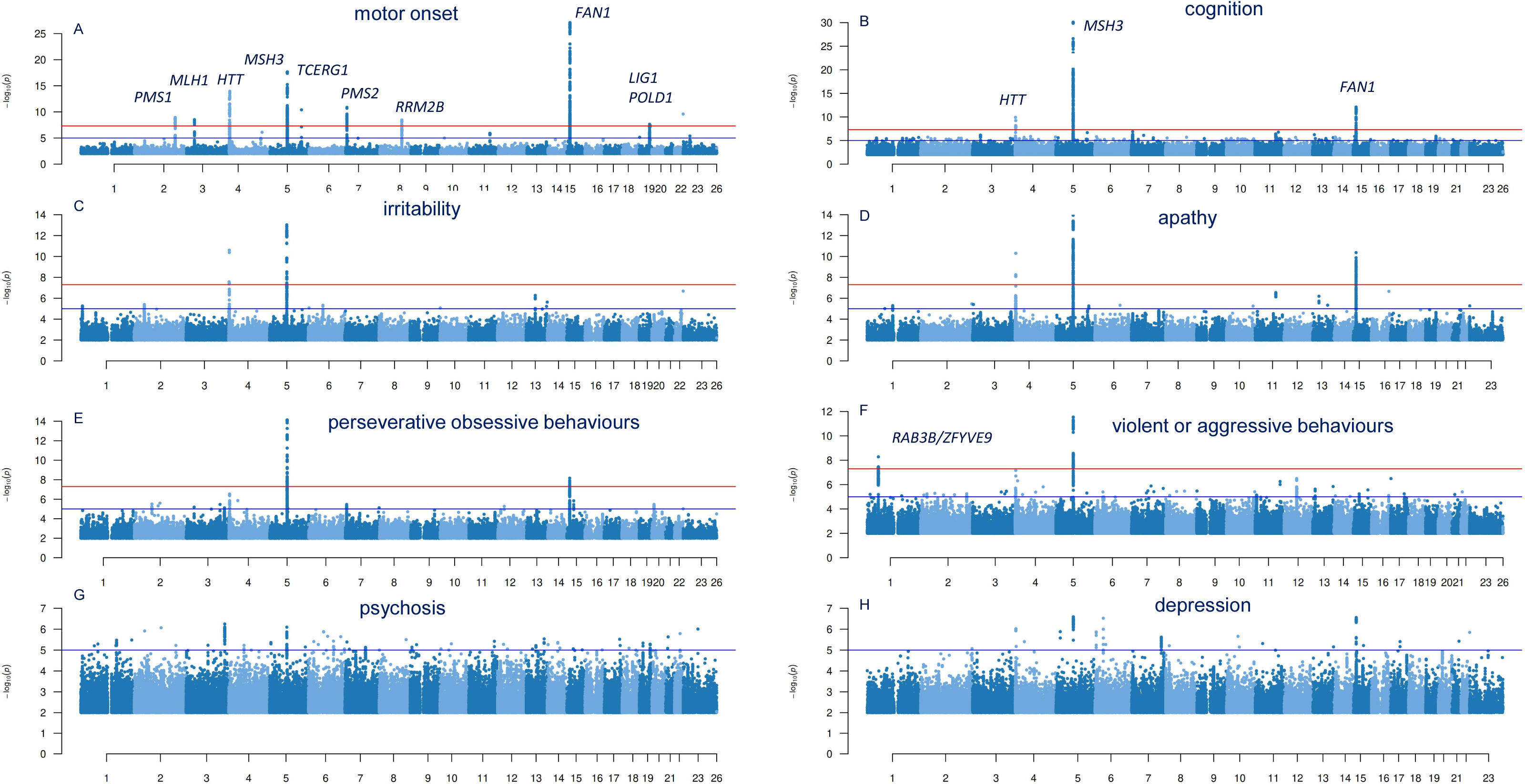
Manhattan plots of meta-analysis results from time-to-event mixed-effects survival (frailty) models evaluating genetic modifiers of clinical symptom onset. The red horizontal line indicates the genome-wide significance threshold (5×10⁻□), and the blue line marks the suggestive threshold (1×10⁻□). Signals at *MSH3*, *HTT*, and *FAN1* represent modifiers shared across motor, cognitive, and psychiatric onset, alongside additional trait-preferential loci.

**Figure 2.**
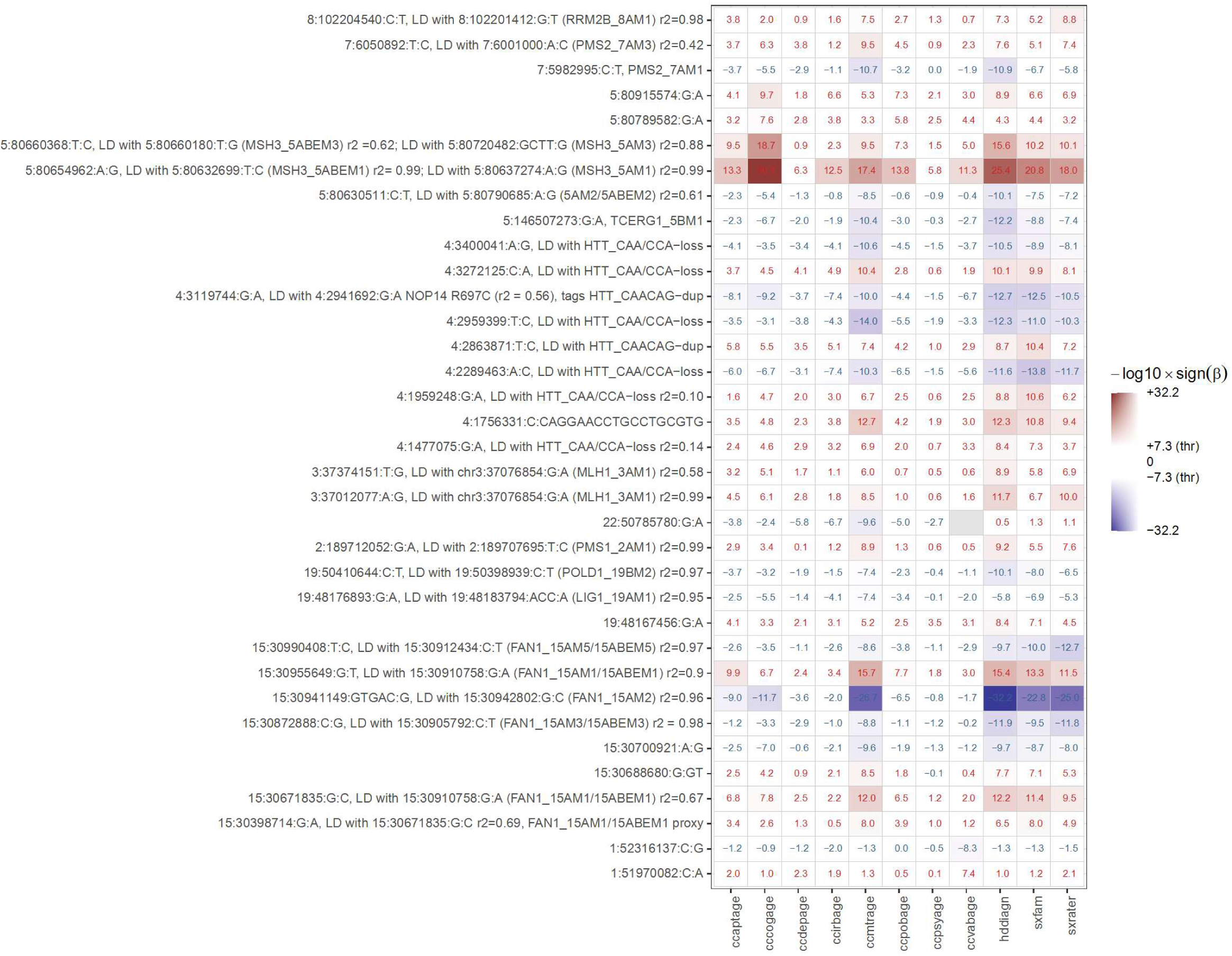
Meta-analysis results for 35 genome wide significant independent signals (linkage disequilibrium (LD) r2 < 0.1, corresponding to **Supplementary Table 4**) from 12 genomic regions across 11 traits. Most overlap with previously reported signals, while *MSH3* and *FAN1* had additional independent signals as illustrated via linkage disequilibrium (LD) structure of the *MSH3* and *FAN1* locus (**Supplementary Figures 4 and 5**). Note that the GWAS results for subject-reported age at HD diagnosis were noisy and were excluded from LD clumping.

**Table 1.**
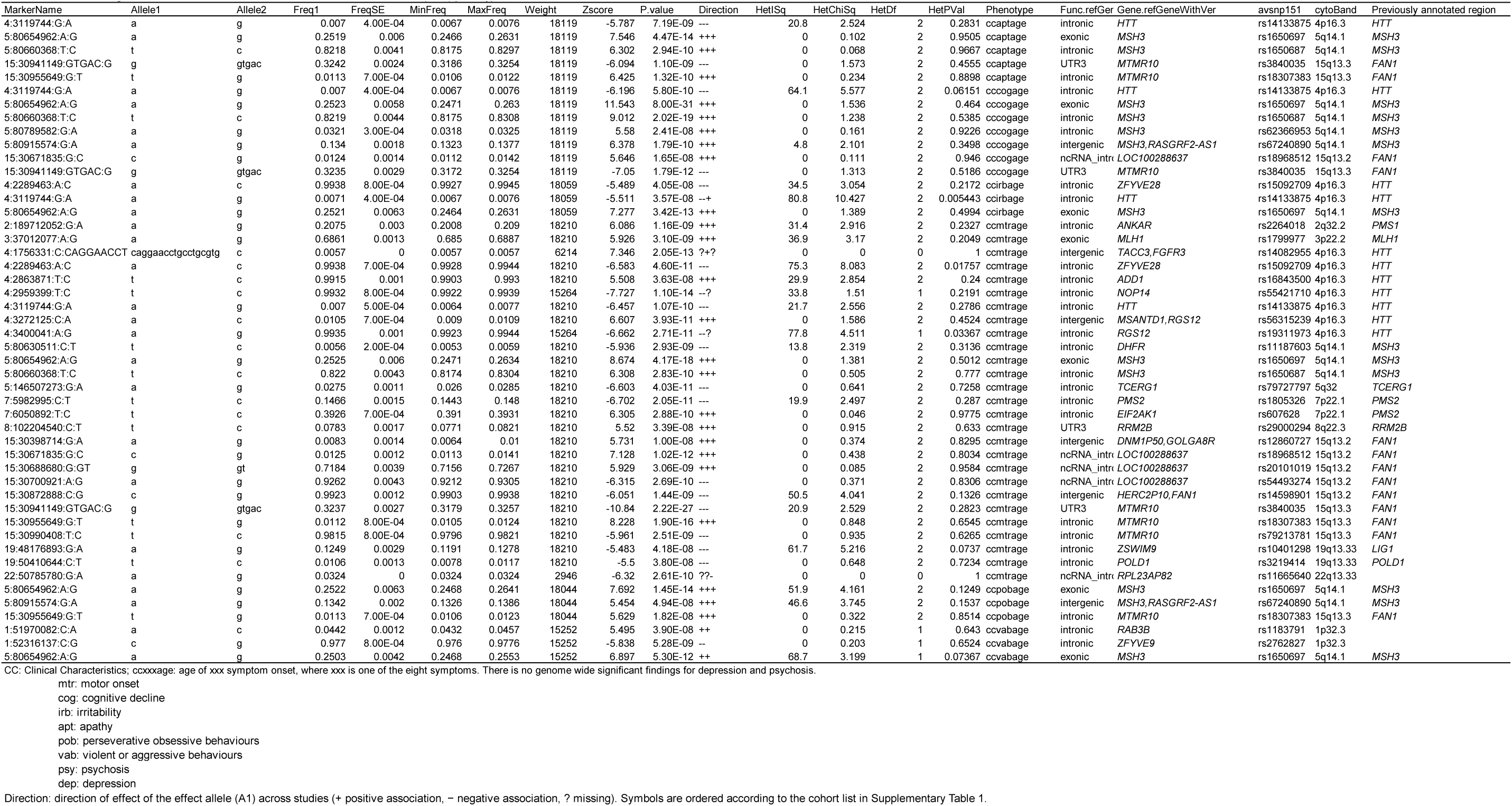
Genome wide significant lead variants for 8 HD Clinical Characteristic Questionnaire (CCQ) phenotypes.

### PGS association with 12 HD phenotypes

Among 13 PGSs derived from psychiatric, neurological, and cognitive / behavioral traits, depressed affect^13^ was associated with onset of all HD signs except motor onset and age at HD diagnosis, with hazard ratio > 1, suggesting the genetic burden of depressed affect is associated with earlier age of onset (**Table 2**, **Supplementary Tables 9 and 10**). The strongest association was with age at depressive symptom onset (p = 1.61×10^−21^). Likewise, PGS derived from major depressive disorder (MDD)^14^ was associated with 11 phenotypes (except age at HD diagnosis estimated by clinician); again, the strongest association was with age at depressive symptom onset (p = 3.57 x 10^−34^), and the association with age at motor onset was on the weaker end (p = 1.97×10^−5^). As expected, schizophrenia^15^ PGS was associated with age at psychosis symptom onset (p = 1.43×10^−14^). Both observations along with other PGS associations supported the notion that the genetic liability to psychiatric conditions/traits derived from the general population influenced age of onset of psychiatric signs. However, the PGS for AD^16^ and PD^17^ was not associated with any phenotype examined, including age of onset of cognitive and motor symptoms. Strikingly, education attainment (EA)^18^ PGS was negatively associated with the reported age of emergence of violent or aggressive behavior (p = 9.15 x 10^−21^), irritability (p = 9.49 x 10^−16^), apathy (p = 1.14×10^−13^), and perseverative obsessive behavior symptoms (p = 6.28 x 10^−10^).

**Table 2.** Polygenic score (PGS) association with HD phenotypes using Cox regression model adjusting for CAG length, sex, and the first two principal components representing population substructure.

| Base | Phenotype | N | Event | coef | exp(coef) | P | CI |
| --- | --- | --- | --- | --- | --- | --- | --- |
| **Age at apathy symptom onset** |  |  |  |  |  |  |  |
| MDD | ccaptage | 9011 | 5378 | 0.276 | 1.317 | 1.99E-14 | 1.23-1.41 |
| EA | ccaptage | 9011 | 5378 | -0.303 | 0.738 | 1.14E-13 | 0.68-0.8 |
| depressed_affect | ccaptage | 9011 | 5378 | 0.466 | 1.594 | 7.44E-12 | 1.39-1.82 |
| neuroticism | ccaptage | 9011 | 5378 | 0.403 | 1.496 | 3.11E-11 | 1.33-1.68 |
| depression | ccaptage | 9011 | 5378 | 0.591 | 1.806 | 2.90E-08 | 1.47-2.23 |
| worry | ccaptage | 9011 | 5378 | 0.3 | 1.349 | 7.87E-06 | 1.18-1.54 |
| intelligence | ccaptage | 9011 | 5378 | -0.177 | 0.838 | 1.91E-05 | 0.77-0.91 |
| insomnia | ccaptage | 9011 | 5378 | 0.201 | 1.222 | 3.15E-05 | 1.11-1.34 |
| CP | ccaptage | 9011 | 5378 | -0.16 | 0.852 | 5.29E-05 | 0.79-0.92 |
| **Age at cognitivesymptom onset** |  |  |  |  |  |  |  |
| MDD | cccogage | 8994 | 4362 | 0.253 | 1.288 | 1.86E-10 | 1.19-1.39 |
| neuroticism | cccogage | 8994 | 4362 | 0.305 | 1.356 | 6.05E-06 | 1.19-1.55 |
| depressed_affect | cccogage | 8994 | 4362 | 0.32 | 1.377 | 2.11E-05 | 1.19-1.6 |
| CP | cccogage | 8994 | 4362 | -0.17 | 0.844 | 1.28E-04 | 0.77-0.92 |
| intelligence | cccogage | 8994 | 4362 | -0.168 | 0.846 | 3.21E-04 | 0.77-0.93 |
| **Age at depressive symptom onset** |  |  |  |  |  |  |  |
| MDD | ccdepage | 9021 | 6493 | 0.398 | 1.489 | 3.57E-34 | 1.4-1.59 |
| depressed_affect | ccdepage | 9021 | 6493 | 0.587 | 1.798 | 1.61E-21 | 1.59-2.03 |
| neuroticism | ccdepage | 9021 | 6493 | 0.501 | 1.65 | 9.29E-20 | 1.48-1.84 |
| depression | ccdepage | 9021 | 6493 | 0.73 | 2.075 | 7.14E-14 | 1.71-2.51 |
| worry | ccdepage | 9021 | 6493 | 0.379 | 1.46 | 6.91E-10 | 1.29-1.65 |
| BIP | ccdepage | 9021 | 6493 | 0.095 | 1.1 | 5.36E-09 | 1.07-1.14 |
| SCZ | ccdepage | 9021 | 6493 | 0.063 | 1.066 | 1.87E-07 | 1.04-1.09 |
| EA | ccdepage | 9021 | 6493 | -0.165 | 0.848 | 8.51E-06 | 0.79-0.91 |
| intelligence | ccdepage | 9021 | 6493 | -0.14 | 0.87 | 2.58E-04 | 0.81-0.94 |
| insomnia | ccdepage | 9021 | 6493 | 0.16 | 1.174 | 2.90E-04 | 1.08-1.28 |
| **Age at irritability symptom onset** |  |  |  |  |  |  |  |
| MDD | ccirbage | 8986 | 6012 | 0.299 | 1.348 | 1.40E-18 | 1.26-1.44 |
| depressed_affect | ccirbage | 8986 | 6012 | 0.527 | 1.694 | 1.52E-16 | 1.49-1.92 |
| neuroticism | ccirbage | 8986 | 6012 | 0.466 | 1.593 | 3.72E-16 | 1.42-1.78 |
| EA | ccirbage | 8986 | 6012 | -0.309 | 0.734 | 9.49E-16 | 0.68-0.79 |
| depression | ccirbage | 8986 | 6012 | 0.701 | 2.017 | 4.31E-12 | 1.65-2.46 |
| worry | ccirbage | 8986 | 6012 | 0.339 | 1.404 | 7.94E-08 | 1.24-1.59 |
| intelligence | ccirbage | 8986 | 6012 | -0.196 | 0.822 | 5.82E-07 | 0.76-0.89 |
| insomnia | ccirbage | 8986 | 6012 | 0.225 | 1.252 | 8.93E-07 | 1.14-1.37 |
| CP | ccirbage | 8986 | 6012 | -0.181 | 0.834 | 1.41E-06 | 0.78-0.9 |
| SCZ | ccirbage | 8986 | 6012 | 0.049 | 1.05 | 1.31E-04 | 1.02-1.08 |
| **Age at motor symptom onset** |  |  |  |  |  |  |  |
| MDD | ccmtrage | 9050 | 7237 | 0.134 | 1.143 | 1.97E-05 | 1.08-1.22 |
| **Age at perseverative obsessive behaviour symptom onset** |  |  |  |  |  |  |  |
| MDD | ccpobage | 8954 | 4728 | 0.255 | 1.291 | 2.56E-11 | 1.2-1.39 |
| EA | ccpobage | 8954 | 4728 | -0.268 | 0.765 | 6.28E-10 | 0.7-0.83 |
| neuroticism | ccpobage | 8954 | 4728 | 0.387 | 1.472 | 2.21E-09 | 1.3-1.67 |
| depressed_affect | ccpobage | 8954 | 4728 | 0.402 | 1.496 | 2.93E-08 | 1.3-1.72 |
| depression | ccpobage | 8954 | 4728 | 0.611 | 1.843 | 6.85E-08 | 1.48-2.3 |
| SCZ | ccpobage | 8954 | 4728 | 0.075 | 1.078 | 1.27E-07 | 1.05-1.11 |
| worry | ccpobage | 8954 | 4728 | 0.342 | 1.408 | 1.66E-06 | 1.22-1.62 |
| CP | ccpobage | 8954 | 4728 | -0.197 | 0.821 | 2.93E-06 | 0.76-0.89 |
| BIP | ccpobage | 8954 | 4728 | 0.083 | 1.087 | 1.41E-05 | 1.05-1.13 |
| intelligence | ccpobage | 8954 | 4728 | -0.173 | 0.841 | 8.73E-05 | 0.77-0.92 |
| insomnia | ccpobage | 8954 | 4728 | 0.194 | 1.214 | 1.69E-04 | 1.1-1.34 |
| **Age at psychosis symptom onset** |  |  |  |  |  |  |  |
| SCZ | ccpsyage | 9069 | 1035 | 0.233 | 1.263 | 1.43E-14 | 1.19-1.34 |
| MDD | ccpsyage | 9069 | 1035 | 0.434 | 1.543 | 1.11E-07 | 1.31-1.81 |
| neuroticism | ccpsyage | 9069 | 1035 | 0.63 | 1.878 | 5.46E-06 | 1.43-2.46 |
| depressed_affect | ccpsyage | 9069 | 1035 | 0.659 | 1.934 | 2.24E-05 | 1.43-2.62 |
| **Age at violent or aggressive behaviour symptom onset** |  |  |  |  |  |  |  |
| EA | ccvabage | 9031 | 3520 | -0.47 | 0.625 | 9.15E-21 | 0.57-0.69 |
| depressed_affect | ccvabage | 9031 | 3520 | 0.729 | 2.073 | 2.67E-18 | 1.76-2.44 |
| neuroticism | ccvabage | 9031 | 3520 | 0.626 | 1.87 | 6.07E-17 | 1.62-2.17 |
| MDD | ccvabage | 9031 | 3520 | 0.364 | 1.44 | 2.22E-16 | 1.32-1.57 |
| depression | ccvabage | 9031 | 3520 | 1.007 | 2.736 | 2.17E-14 | 2.11-3.54 |
| intelligence | ccvabage | 9031 | 3520 | -0.371 | 0.69 | 5.11E-13 | 0.62-0.76 |
| CP | ccvabage | 9031 | 3520 | -0.342 | 0.71 | 3.38E-12 | 0.65-0.78 |
| SCZ | ccvabage | 9031 | 3520 | 0.092 | 1.096 | 2.53E-08 | 1.06-1.13 |
| worry | ccvabage | 9031 | 3520 | 0.439 | 1.551 | 1.04E-07 | 1.32-1.82 |
| insomnia | ccvabage | 9031 | 3520 | 0.275 | 1.316 | 4.21E-06 | 1.17-1.48 |
| **Age at HD diagnosis** |  |  |  |  |  |  |  |
| MDD | hddiagn | 8748 | 6895 | 0.143 | 1.154 | 8.63E-06 | 1.08-1.23 |
| MDD | sxfam | 7960 | 6462 | 0.136 | 1.146 | 4.35E-05 | 1.07-1.22 |
| EA | sxfam | 7960 | 6462 | -0.134 | 0.875 | 3.21E-04 | 0.81-0.94 |

<sup>a</sup>HR: hazard ratio, i.e. exp(coef)
MDD: major depressive disorder
EA: education attainment
CP: cognitive performance
PGC summary statistics were downloaded from https://pgc.unc.edu/for-researchers/download-results/. Additional summary statistics were downloaded from https://cncr.nl/research/summary_statistics/.
CC: Clinical Characteristics; cxxxxage: age of xxx symptom onset, where xxx is one of the eight symptoms. There is no genome wide significant findings for depression and psychosis.
mtr: motor onset
cog: cognitive decline
irb: irritability
apt: apathy
pob: perseverative obsessive behaviours
vab: violent or aggressive behaviours
psy: psychosis

### ExWAS Results

Across 12 phenotypes, seven genes reached study-wide significance, defined by a significant Cauchy test after multiple testing correction (p < 3.95×10⁻□), mask-level p ≤ 3.29×10⁻□, and minor allele count (MAC) ≥ 20 (**Supplementary Tables 11 and 12; Figure 3; Supplementary Figures 6 and 7**).

**Figure 3.**
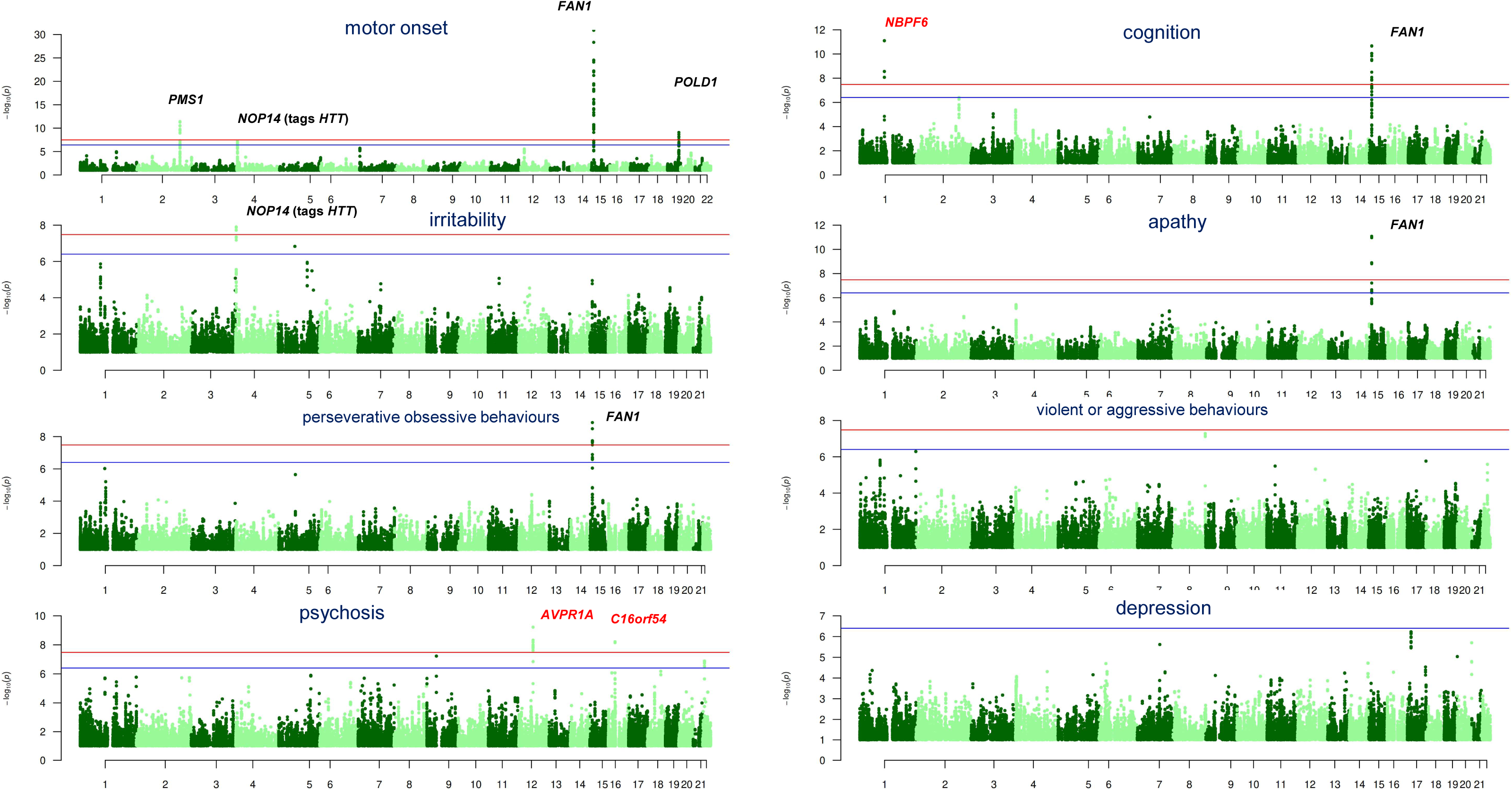
Manhattan plots of rare variant gene-level test from time-to-event mixed-effects survival (frailty) models evaluating genetic modifiers of symptom onset. Mask-level (minimum MAC of 20 (A), 10 (B), and 5 (C)) association p-values are plotted. Red line denotes mask-level *p* < 0.05/(2*3*3*4*# of genes tested ∼ 3.29 x 10^−8^) and the blue line marks the Cauchy p-value threshold (p < 0.05(2*3*# of genes tested).

Four loci - *FAN1*, *PMS1*, *POLD1*, and *HTT* - represent high-confidence findings, as they overlap with previously implicated HD modifier genes associated with age at clinical landmarks and/or somatic instability^5^ (**Supplementary Tables 13–16; Supplementary Figures 8–11**). These genes show associations across multiple phenotypes and converge on DNA repair and somatic instability, supporting these pathways as central drivers of disease progression.

*FAN1* exhibited the strongest and most consistent associations, including time to motor, cognitive, apathy, and perseverative/obsessive symptom onset, as well as multiple definitions of age at HD diagnosis. Associations were highly significant (Cauchy p-values 1.53×10⁻³□ to 6.88×10⁻¹□ for time to motor onset) and were not solely driven by known hastening modifiers (R507H, R377W), but also by additional rare variants (R145H, P654L) and ultra-rare missense and pLoF variations (**Supplementary Table 13**).

*PMS1* and *POLD1* were significantly associated with time to motor onset and age at HD diagnosis, with evidence for allelic heterogeneity. For *PMS1*, associations (Cauchy p-values 8.61×10⁻¹¹ to 1.78×10⁻□) were driven by delaying rare SNVs (including T75I and R26K) and ultra-rare missense and pLoF variants; notably, T75I has previously been associated with decreased repeat expansion^19^ (**Supplementary Table 14**). For *POLD1*, associations (Cauchy p-values 1.68×10⁻□ to 1.11×10⁻□) reflect contributions from delaying rare SNVs (including R30W, Q710H, and R875H) as well as ultra-rare missense and pLoF variants (**Supplementary Table 15**). *POLD1* was additionally associated with age at diagnosis using family-reported measures.

Evidence for the *HTT* locus arises from *NOP14* and *SH3BP2* signals tagging the *HTT* non-canonical CAACAG duplication. *NOP14* was associated with time to irritability onset and age at HD diagnosis, with the signal largely driven by R697C, which tags the *HTT* non-canonical CAACAG duplication^6^ (Cauchy p-values 2.85×10^−□^ to 4.28×10^−□^; **Supplementary Table 16**). Consistent with this, *SH3BP2* emerged as an additional high-confidence signal when stop-loss variants were included as pLoF variants, with the driving variant (X562W) also tagging the CAACAG duplication (**Supplementary Table 17; Supplementary Figure 12**). In both cases, when conditioning on *HTT* alleles, the association for the respective variants (R697C or X562W) became insignificant, suggesting that the *HTT* non-canonical alleles are the causal signal (**Supplementary Table 16 D** and **Supplementary Table 17 D).**

These signals at MMR genes and the *HTT* locus are consistent with findings from a parallel and co-submitted companion study^20^, with differences attributable to analytical pipelines, and further support somatic expansion as a key determinant of clinical variability.

Two additional genes reaching significance (**Supplementary Table 11**) map to segmental duplication or repeat regions and require cautious interpretation with respect to genotype quality and causal inference (*NBPF6*, **Supplementary Table 18; Supplementary Figure 13**; *C16orf54*, deprioritized after manual review).

*AVPR1A* was associated with age of psychosis onset, driven primarily by a single rare coding variant (F308L; HR ∼2.75, p = 8.69×10⁻□) with minor contribution from ultra-rare variants (**Supplementary Table 19**). *AVPR1A* encodes a vasopressin receptor implicated in social behavior and stress response, with relevance to neuropsychiatric phenotypes and prior investigation as a therapeutic target for irritability and aggression in HD^21,22^.

Additional genes meeting a relaxed study-wide threshold (Cauchy p < 3.95×10⁻□, mask-level p ≤ 10⁻□, MAC ≥ 5) are reported in **Supplementary Table 11**. Notably, DNA damage response genes *CHTF8* (Cauchy p-values 8.94×10⁻□ to 1.21×10⁻□; MAC = 13; **Supplementary Table 20; Supplementary Figure 14, Supplementary Results**) and *PSME4* (Cauchy p-value = 2.34×10⁻□; MAC = 7; **Supplementary Table 21; Supplementary Figure 15, Supplementary Results**) emerged as candidates supported by ultra-rare variation. These, together with broader pathway enrichment signals - including protein homeostasis and ubiquitin–proteasome function, RNA processing and nucleolar stress, vesicle trafficking (such as *BIN1* and *TMED1*) and synaptic signaling, immune response, and lipid metabolism—highlight additional biological processes that may contribute to HD modification (**Supplementary Tables 11 and 12**). Detailed gene-level and variant-level results are provided in **Supplementary Tables 22–30 and Supplementary Figures 16–25**.

Finally, a suggestive association at *ZFAND2A* (Cauchy p-values 1.16×10⁻□ to 2.34×10⁻□; MAC = 4) was observed for both motor onset and age at diagnosis (**Supplementary Table 31; Supplementary Figure 26**), further implicating proteasome-associated stress response pathways.

A more detailed description of the ExWAS results is provided in the **Supplementary Results**.

### Pathway analysis results

Consistent with prior studies, genome-wide significant loci for motor symptom onset were strongly enriched for MMR pathway genes. This signal was recapitulated in MAGMA pathway analysis^23^ using gene-level association results derived from GWAS, where the top REACTOME term (“Diseases of Mismatch Repair”) and a Gene Ontology cellular component term (“mismatch repair complex”) reached Bonferroni-adjusted significance (p = 7.23×10⁻□ and 4.09×10⁻□, respectively; **Supplementary Table 32**). Notably, the REACTOME gene set is small (n = 5) and includes *MSH3* (p_gene_ *=* 1.36×10^−13^), *PMS2* (p_gene_ *=* 3.28×10^−9^), *MLH1* (p_gene_ *=* 0.0003), *MSH6* (p_gene_ *=* 0.005), and *MSH2* (p_gene_ *=* 0.44), indicating that this enrichment is largely driven by high-confidence GWAS loci.

In contrast, larger gene sets showed broader but less pronounced enrichment. For example, the Gene Ontology molecular function term “DNA damage sensor activity” (n = 20) achieved a nominal p-value of 1.10×10⁻□ and Bonferroni-corrected significance (p = 0.02; **Supplementary Table 32**). Similarly, the KEGG mismatch repair pathway (n = 22) showed nominal enrichment (p = 3.22×10⁻³), suggesting involvement of a wider set of DNA damage response (DDR) and MMR genes but with more diffuse statistical support across the pathway. Importantly, MAGMA pathway analysis evaluates the full distribution of gene-level association statistics rather than relying solely on genome-wide significant loci. Thus, enrichment observed in larger gene sets reflects coordinated, sub-threshold signal across multiple genes rather than dominance by a small number of highly significant loci.

Consistent with this, gene set enrichment analysis, which similarly evaluates the full distribution of gene-level association statistics, applied to ExWAS results identified enrichment of the KEGG_MEDICUS_REFERENCE_MISMATCH_REPAIR pathway with modest significance (q = 0.04), driven by a broader collection of MMR genes (*POLE*, *POLD2*, *RFC1*, *POLD3*, *POLE2*, *MLH3*, *LIG1*, *MSH3*, *POLD1*, *PMS1*; **Supplementary Table 33**; **Supplementary Figure 27**). Within this pathway (n = 26), several genes, including *MSH3* (Cauchy SKAT-O p = 3.78×10⁻³, **Supplementary Table 34**), *MLH3*, and *LIG1*, showed nominal evidence of association at the gene level, indicating that ExWAS captures additional MMR-related signals beyond genome-wide significant loci *PMS1* and *POLD1*.

To further interpret these findings in a biological context, we applied network-based reprioritization (NetWAS)^24^, which integrates gene-level association statistics (derived using MAGMA based on GWAS results) with tissue-specific interaction networks to prioritize genes based on both statistical evidence and functional connectivity. NetWAS analysis highlighted MMR and broader DDR pathways, including genes not reaching significance in GWAS, reinforcing the central role of DNA repair processes while extending the signal to a wider network of repair-associated genes (**Supplementary Table 35**; see **Supplementary Methods and Results**), which requires experimental validation.

ExWAS results across phenotypes were also tabulated for MMR genes identified in GWAS and for selected top-ranked DDR genes from the NetWAS reprioritization analysis, providing a cross-method view of pathway-level signals (**Supplementary Tables 36 and 37**).

Together, these analyses indicate that the strongest genetic effects are concentrated in a small set of core MMR genes, while complementary approaches that leverage genome-wide signal distributions extend this finding to a broader, but more modestly supported, network of DNA damage response pathways.

## Discussion

The Enroll-HD natural history platform which continues to add rich phenotypic data provides the motivation and value of WGS data generation for diverse genotype-phenotype association analyses. Our present study provides the first genetic dissection of reported emergence of psychiatric signs in HD; the associations observed suggest a hybrid genetic architecture that integrates general psychiatric liability with repeat expansion disease (RED^25^, including HD)-specific burden. Onset of cognitive and motor signs, on the other hand, do not appear to share genetic liability with that of other dementia conditions such as AD and PD. Given that the earliest brain regions affected—entorhinal cortex (medial temporal lobe) and hippocampus for AD, and dorsal motor nucleus of the vagus (DMV) in the brainstem for PD—these findings might be anticipated. Psychiatric PGS predicted earlier reported age of onset of multiple psychiatric signs, suggesting that the same common-variant liability influencing primary psychiatric disorders may also contribute to psychiatric vulnerability in HD. At the same time, several established HD motor modifiers—most prominently *MSH3*, *FAN1*, and *HTT*—also influenced psychiatric signs, supporting a model in which psychiatric symptoms reflect both shared and disease-specific genetic mechanisms. Even though currently there is no genome wide significant common variant associations for depression and psychosis, there are suggestive association signals for *MSH3* (p = 4.72×10^−7^ and p = 1.59×10^−6^ for depression and psychosis, respectively, for variant 5:80654962:A:G, which is in LD with 5:80632699:T:C (5ABEM1, r^2^= 0.99, **Supplementary Figure 4**) and 5:80637274:A:G (5AM1, r^2^=0.94, **Supplementary Figure 4**)) and *HTT* (p = 8.34 x 10^−5^ for depression and 4:3272125:C:A, which is in weak LD with CAA/CCA-loss, r^2^ = 0.16, **Supplementary Table 4**), respectively. This conclusion is consistent with the notion from neuroanatomy that HD symptoms arise from dysfunction in three partially distinct cortico-striatal loops - sensorimotor, associative, and limbic^26^. Shared genetic modifiers—primarily mismatch-repair genes influencing somatic CAG expansion—affect vulnerable cells contributing to all circuitry loops. The relative strength of common variant associations differs by phenotype, with *FAN1* showing stronger effects for motor onset, and *MSH3* demonstrating stronger associations for cognitive and psychiatric outcomes. Additional rare variant modifiers also show domain preferential associations, with *PMS1* and *POLD1* significantly associated with motor phenotypes and *FAN1* influencing motor, cognitive, and psychiatric domains, acknowledging potential differences in statistical power across phenotypes. This architecture explains why HD symptoms are partly shared and partly distinct at both circuit and genetic levels.

The contribution of general psychiatric liability to symptom onset in HD is notable, as it suggests that psychiatric symptoms in a monogenic neurodegenerative disorder are shaped in part by the same polygenic architecture that influences risk for depression, irritability, psychosis, and related traits in the general population. This finding aligns with the NIH / NIMH Research Domain Criteria (RDoC) framework^27^ and growing recognition that psychiatric symptoms are dimensional features cutting across diagnostic categories sharing substantial genetic liability. HD provides a window to observe these effects in a monogenic genetic background. The observation that psychiatric polygenic scores are linked with estimated symptom emergence even in the presence of a highly penetrant monogenic mutation underscores the pervasive influence of common-variant psychiatric risk across diverse biological contexts.

In parallel, our results demonstrate that HD-specific somatic instability pathways also contribute to psychiatric symptom onset. *MSH3* and *FAN1*, two of the most robust modifiers of motor onset, were associated with earlier onsets of multiple psychiatric symptoms, including apathy, irritability, perseverative behavior and aggression. These findings extend the role of somatic instability beyond motor and cognitive domains and suggest that instability-driven neuronal dysfunction may influence brain regions and circuits relevant to psychiatric symptom manifestation. The involvement of *HTT*-tagging variants further supports the idea that mechanisms intrinsic to the expanded CAG repeat and time to clinical landmarks contribute to psychiatric vulnerability^5,28^. Furthermore, across REDs, psychiatric symptoms are also prevalent for myotonic dystrophy (DM)^29^, fragile X (FX) syndrome^30^, and spinocerebellar ataxia (SCA)^31^. Together, these cross-disease patterns hypothesize a unified model in which MMR-driven somatic expansion amplifies repeat toxicity in interconnected striatal, cerebellar, and limbic circuits, yielding convergent psychiatric phenotypes across HD, DM1, FX, and SCAs.

Importantly, not all HD modifiers exhibited cross-domain effects. *PMS1* and *POLD1* (for both common and rare variants), as well as *PMS2* and *MLH1* (for common variants), strongly influenced motor onset and age at HD diagnosis but showed no detectable effects on psychiatric symptom onset. There is a trend association between *PMS2* and *MLH1* common variants and cognitive symptom onset (**Figure 2**). We cannot exclude the possibility that the lack of/weak association reflects limited statistical power at this stage. The coupling of MutSβ (MSH2-MSH3)-MutLγ (MLH1-MLH3) has been well appreciated, with MutSβ-MutLγ-driven repeat expansion biochemically reconstituted using purified human proteins, whereas MutLβ (MLH1-PMS1) has not been shown to exhibit comparable endonuclease activity in repeat expansion assays; however, in HD mouse and HD cell models *Pms1*/*PMS1* loss-of-function alleles can halt *HTT* CAG SI^32–36^. Blood-based SI GWAS nevertheless also implicated other MMR pathway players (i.e. MutLα (MLH1-PMS2 via PMS*2*) and MutSα (MSH2-MSH6)), in addition to *MSH3, MLH3*, and *FAN1* which also plays an important role in repeat instability. No direct SI genetic study has been conducted in the central nervous system due to limited sample size. Late clinical landmark GWAS, which captures the cumulative effects of repeat expansion and downstream mHTT toxicity, implicated both MutLα and MutLβ with association signals stronger for *PMS1* and *PMS2* than *MLH3*^5^. In the present study, the previously reported suggestive *MLH3* association with age at a score of 6 on the 13-point Total Functional Capacity scale (TFC6) was only nominally significant in the motor symptom-onset GWAS (p = 0.01 for 14:75140342:G:A). By contrast, in *in vivo* CRISPR–Cas9 genome-editing experiments in HD mouse models, the effect of *Mlh3* appeared stronger than that of *Pms1*^35^. Notably, in this study, *PMS1* rare variants are suggestively associated with cognitive symptom onset (**Supplementary Table 36**), although genome wide association signals for *PMS1*, *MLH1*, and *PMS2*, respectively, have been reported for late cognitive landmarks such as age at 25% quantile of Symbol Digit Modalities Test (SDMT) and Stroop Word^5^. This difference suggests that the genetic architecture of HD is not uniformly shared across symptom domains and that distinct biological pathways contributing to motor versus psychiatric and cognitive manifestations may be context or stage dependent. The presence of both shared and domain-preferential modifiers provides a mechanistic explanation for the well-recognized clinical heterogeneity of HD and highlights that targeting *MSH3* and *FAN1* may modify the disease course across all three symptom domains.

Rare variant analyses further broadened the landscape of potential modifiers, identifying additional genes involved in DNA damage response, proteostasis, vesicle trafficking, and synaptic function. Although some of these findings require replication due to modest allele counts, they converge on biological pathways that have been implicated in HD pathogenesis and psychiatric disorders more broadly. The potential association of *RAB3B* with violent or aggressive behavior onset, for example, aligns with the role of RAB family proteins in synaptic vesicle cycling and neurotransmitter release^37,38^. It is interesting to note that several rare variant modifier candidates *BIN1*^39,40^ and *TMED1*^41^ are involved in vesicle trafficking. These emerging pathways suggest that psychiatric symptoms in HD may arise from disruptions in neuronal signaling and cellular stress responses that intersect with, but are not limited to, somatic instability mechanisms. Among the known modifiers, the association signals for *MSH3* and *FAN1* are complex with few independent novel signals. In a model of HD pathogenesis, transcriptional regulation of MMR genes determines the cell types vulnerable to somatic instability, followed by DNA methylation stabilization of a toxicity effect of mutant HTT protein via interaction with MED15 and TCERG1, which regulate enhancer activity and transcriptional elongation^12^. The variants in the super-enhancer region of *MSH3* and *FAN1,* in LD with 5AM1, 5AM3 or 15AM2, respectively, offer a direct testable hypothesis on how these variants could alter the super-enhancer activity. Future rare variant analysis focusing on regulome, meQTL analysis, and long-range chromatin interaction in vulnerable brain regions and cell types could further identify modifier genes of interest and shed light on additional regulatory mechanisms.

In aggregate, these findings support a model where (1) *HTT* CAG instability is necessary for pathogenic phenotypes and reflected in the associations for DNA repair and handling modifier genes with psychiatric symptoms, and (2) psychiatric symptoms in HD reflect the combined influence of general psychiatric liability and HD-specific molecular pathology. This hybrid architecture provides a conceptual bridge between psychiatric genetics and neurodegeneration, illustrating how common-variant psychiatric risk can interact with disease-specific mechanisms to shape symptom manifestation. The results also underscore the importance of considering psychiatric symptoms as intrinsic components of HD biology rather than secondary consequences of motor decline, or psychological stress from knowing the risk of carrying the expanded allele of the *HTT* CAG repeat or being informed of one’s HD diagnosis.

Several limitations should be acknowledged. Psychiatric symptom onset is inherently more variable and subject to greater measurement noise than motor onset, and ascertainment may differ across raters and cohorts, though one could argue the same for motor and neurocognitive and the need for more objective assessment. Although our sample size is large, rare variant analyses remain underpowered for many genes, and replication in independent sequencing datasets or increasing the sample size of the current study (since Enroll-HD is still ongoing) will be essential. Additionally, while polygenic scores capture a portion of common-variant liability, they do not fully represent the genetic architecture of psychiatric disorders and symptoms. Future work integrating transcriptomic, epigenetic, and imaging data may help clarify the mechanisms linking genetic modifiers to emergence of psychiatric and other HD symptoms, as well as disease progression.

In conclusion, this study provides an initial utilization of WGS data from 18K Enroll-HD participants, as well as the first systematic evaluation of the genetic architecture underlying psychiatric symptom onset in HD and identifies both shared and domain-preferential genetic pathways.

## Methods

### Clinical cohorts and Clinical phenotypes analyzed

Whole genome sequencing (WGS) data was generated for the samples from Enroll-HD^9^ cohort (n= 18,825) as described in Gelfman et al., 2026^20^. Additional SNP array genotyping data was generated previously for a portion of the Enroll-HD and the EHDN REGISTRY samples^5,10^. The clinical studies were carried out in accordance with the ethical principles outlined in the Declaration of Helsinki, Good Clinical Practices guidelines, and applicable regulatory requirements. The study protocols were approved by the local, regional, or central Institutional Review Board (IRB) or Independent Ethics Committee (IEC) or Research Ethic Board (REB) overseeing the respective clinical sites. All participants provided written informed consent before enrollment.

We examined age at onset of HD symptoms including motor, cognitive, and six psychiatric symptoms, as well as age at HD diagnosis (4 ascertainments). Estimated age of onset of psychiatric symptoms, cognitive impairment, and HD-related motor symptoms were collected using the HD Clinical Characteristics Questionnaire (CCQ). The HD CCQ is a clinician-completed instrument that captures retrospectively reported history and estimated age at emergence of symptoms based on all available information, including participant, observer reports, medical records, and clinical rating scales. HD CCQ was previously used in the REGISTRY study^42^. A publicly hosted version of the questionnaire is available at <u>Zenodo</u> and <u>Enroll-HD-AnnotatedCRF_2020-10-R2.pdf</u>. Among the 8 symptoms assessed, only motor symptoms needed to be HD-specific. For the age at HD diagnosis, the first symptom leading to HD diagnosis can be motor, cognitive, or psychiatric. Among the 4 ascertainment approaches, age at HD diagnosis variable hddiagn represented when the HD diagnosis was communicated to the patients, while the other variables (sxrater, sxfam, and sxsubj) represented the age of HD diagnosis estimated by clinical rater, family member, and patient, respectively. Phenotypes from Enroll-HD Periodic Dataset version 7 (PDS7) were used for all analysis except for Enroll-HD cohort genotyped by microarray, which used PDS6. Phenotypes from REGISTRY-RDS-2018-07-R2 were used for the REGISTRY cohort analysis. Participants with CAG length, based on PDS6/7 as determined by PCR fragment size assay, greater than 35 were included in the genetic analysis.

### Variant annotation

Variant mapping and annotation were based on the GRCh38 Human Genome reference sequence and all annotated transcripts in RefSeq Gene using ANNOVAR software^43^. Variants were categorized as putative loss-of function (pLoF) if annotated as stop gained, start loss, frameshift, or splice site variants (i.e., within 2-bp away from an exon/intron boundary that captures the core splice site dinucleotides). Missense variants were assigned an aggregated predicted deleteriousness score by a weighted sum of deleteriousness predictions from 40+ algorithms precomputed in dbNSFP version 4.7a^44^ including AlphaMissense^45^ and ESM1b^46^, but also includes other deep learning models such as PrimateAI^47^, EVE^48^, DANN^49^, classical ML such as REVEL^50^, evolutionary / constraint algorithms such as GERP++^51^, CADD (hybrid)^52,53^, and rule-based / clinical algorithm such as ClinVar^54,55^. A full list of algorithms as well deleteriousness cutoff and weighting are provided in the **Supplementary Note**.

### Genetic Data QC and Statistical Analyses

Array-based genotyping data had previously undergone quality control and imputation prior to the present analyses^5^. For the whole-genome sequencing dataset, we excluded 45 individuals whose X-chromosome inbreeding coefficient (FHET) fell outside ±0.20, consistent with standard sex-check quality control procedures. Genetic ancestry outliers were removed based on EIGENSTRAT^56,57^ as implemented in RICOPILI^58^, that uses principal component analysis to model population structure. As a result, 17,724 relatively genetic homogeneous samples were retained for downstream analysis. At the variant level, SNPs with a genotyping missingness rate greater than 10% were excluded. Hardy–Weinberg equilibrium (HWE) was evaluated separately in controls and cases using exact tests, and variants with HWE p-values less than 10^−100^ and 10^−150^ were filtered out.

Genome-wide association studies (GWAS) cohorts were constructed by study and genotyping platform, including 1) Enroll-HD participants previously genotyped^5^; 2) New Enroll-HD with WGS data; 3) REGISTRY participants previously genotyped^5^. We analyzed array-based genotypes and WGS data from more than 18,000 individuals across two Enroll-HD^9^ cohorts (previously genotyped by array and newly sequenced) and the EHDN REGISTRY^10^ cohort (previously genotyped using microarray technology). GWAS of time to multiple age of symptom onset phenotypes and time to age at HD diagnosis (hddiagn, sxrater, sxfam, and sxsubj) using mixed effect survival/frailty model^59^, were performed within each cohort, followed by inverse-variance meta-analysis using METAL^60^. Linkage disequilibrium (LD) clumping of the meta-analyzed summary statistics was conducted jointly using PLINK^61,62^ to identify index variants across traits. Additional network-based gene reprioritization is described in **Supplementary Methods**.

For rare variant analyses, we used the Enroll-HD WGS dataset and performed gene-based tests, treating each gene as the analysis unit. We evaluated putative loss-of-function (pLoF) and missense variants both separately and jointly across multiple masking strategies defined by maximum alternate allele frequency (AAF) thresholds of 2%, 1%, 0.1%, and 0.01%. Two weighting schemes were applied. (1) A deleteriousness-informed scheme in which pLoF variants were assigned a weight of 50, while missense variants were weighted using aggregated deleteriousness predictions (maximum weight 40 if all algorithms predicted a deleterious effect, additional details provided in **Supplementary Method**); (2) An inverse-frequency weighting scheme, in which missense variants predicted as benign by all algorithms were excluded; this also enabled direct calculation of the raw minor allele count (MAC) for each mask. Three gene-based association tests were conducted: a burden test (assuming consistent effect direction for variants within a gene) ^63,64^, SKAT (a variance-component test allowing bidirectional effects)^65^, and SKAT-O (a hybrid of burden and SKAT)^66^. For each gene test, results for the three variant categories and 4 AAF bins were aggregated using an omnibus Cauchy combination method, yielding combined statistics for all 12 masks.

Both common-variant (GWAS) and rare-variant (ExWAS) analyses were conducted using the time-to-event mixed-effects frailty model implemented in SAIGE^67^/SAIGE-GENE^68^, which likely improved power by incorporating censored data. In Step 1 (Saige v1.5.0), a null Cox proportional hazards mixed model was fitted, including covariates as fixed effects and a random effect based on the genetic relationship matrix (GRM) to account for relatedness and population structure. In Step 2 (Saige v1.4.0), individual or sets of variants were tested for association with time to event using score tests conditional on the fitted null model. All analyses were adjusted for CAG repeat length, sex, and the first two principal components derived from common variants. To further investigate the relationship between chromosome 4 signals and *HTT* cis-variants, conditional analysis was performed using data described earlier^5^ where *HTT* non-canonical/canonical alleles were characterized using MiSeq (n = 7,553), the variant of interest was fitted in a linear regression model (using age at HD diagnosis for illustration purpose among the subsamples with onset events) in the presence and absence of *HTT* alleles (as a categorical variable and the canonical allele was used as a reference).

For GWAS, we applied the conventional genome-wide significance threshold of 5×10⁻□. For ExWAS, the Cauchy omnibus p-value served as the primary criterion for gene prioritization. To control for multiple testing under low-MAC conditions, where fraility model test statistics may be inflated, three post-ExWAS QC were applied to weed out the likely inflated test statistics: 1) some of the associations were driven by a single outlier subject, the analyses were re-run removing this single outlier subject, which refined the association signals; 2) When there was no rare variant for a given gene, and saddle point approximation was applied for a given UR (ultra-rare collapsed marker) and there was a large difference in p-values between saddle point approximation and normal approximation, the gene-level Saige results was close to the variant-level results with normal approximation, the gene-level test statistics was considered to be inflated and therefore filtered out; 3) Visualization of the relationship between age of onset (or last age observed in the study if censored) vs. CAG length using the R package ggplot2 (v.4.0.0)^69^ and smaller number of associations was marked as low confidence. We defined an ExWAS genome-wide significance threshold of Cauchy p-value to be 3.95×10⁻□ (0.05 divided by 2 weighting schemes × 3 tests × number of genes), and additionally required that at least one mask with *p* < ∼3.29×10⁻□ (0.05 divided by 2 schemes × 3 tests × 3 annotation categories × 4 AAF bins × number of genes) with MAC ≥ 20 (tier 1) or mask with *p* < 1×10⁻^6^ and MAC ≥ 5 (tier 2). Manhattan plots of GWAS and ExWAS results were generated using the R packages ggplotify (v.0.1.2)^70^, qqman (v.0.1.9)^71^, cowplot (v.1.2.0)^72^.

### Polygenic score association with HD phenotypes

PGSs derived from 13 summary statistics (for non-unique traits including psychiatry conditions and traits schizophrenia^15^, major depressive disorder^14^, bipolar disorder^73^, insomnia^74^, neuroticism^13^, neurological conditions including AD and Parkinson’s disease (PD)^16,17^, and cognitive / behavioral traits intelligence, education attainment, and cognitive performance^18,75,76^) were tested for association with HD phenotypes in cohort 1 using LDpred2^77^. A full list of summary statistics used to derive PGS is available in **Supplementary Table S38**. Association p values less than 0.05/(12*13) ∼ 3.21 x 10^−4^ were considered significant.

### Pathway enrichment analysis

Time to motor symptom onset was used as an exemplary phenotype to perform pathway enrichment analysis as the number of significant findings was expected to be the greatest. Multi-marker Analysis of GenoMic Annotation (MAGMA v1.10)^23^ as implemented in Functional Mapping and Annotation (FUMA v1.8.3)^78^ and R package clusterProfiler v4.14.6^79^ were used to perform pathway enrichment analysis for GWAS and ExWAS results, respectively. Both gene-set analyses were performed for curated gene sets and GO terms obtained from the Human Molecular Signatures Database (MSigDB^80,81^, v2023.1.Hs for FUMA and v2025.1.Hs for clusterProfiler). MAGMA performed a multiple regression of SNP Z-scores, using the LD matrix to account for correlation, and produced a gene-level Z-statistic. For ExWAS, minimal p-value across all masks for the SKAT-O results was used as gene-level p-value, signed (directionality from the burden test) −log(p) was used to rank the genes. Both MAGMA and clusterProfiler gene-set analyses used the full distribution of p-values.

## Data Availability

The data used in this study is available from http://www.enroll-hd.org/. A data use agreement is needed to obtain the data used in this study.

## Code availability

All software used in this study is publicly available. EIGENSOFT; RICOPILI; PLINK; SAIGE; METAL; FUMA; NetWAS; LocalZoom, LDpred2; Haploview; ANNOVAR; cowplot, ggplot2, ggplotify, qqman, clusterProfiler R packages.

*Regeneron Genetics Center^2,3^

## RGC Management & Leadership Team

Aris Baras, Gonc□alo Abecasis, Adolfo Ferrando, Giovanni Coppola, Andrew Deubler, Luca A Lotta, John D Overton, Jeffrey G Reid, Alan Shuldiner, Katherine Siminovitch, Jason Portnoy, Marcus B Jones, Lyndon Mitnaul, Alison Fenney, Jonathan Marchini, Manuel Allen Revez Ferreira, Maya Ghoussaini, Mona Nafde, William Salerno, Cristen Willer, Lourdes Crane, Niek Verweij, Eric Jorgenson, and Joseph Pickrell.

## Sequencing & Lab Operations

John D Overton, Christina Beechert, Erin Fuller, Laura M Cremona, Eugene Kalyuskin, Hang Du, Caitlin Forsythe, Zhenhua Gu, Kristy Guevara, Michael Lattari, Alexander Lopez, Kia Manoochehri, Prathyusha Challa, Manasi Pradhan, Raymond Reynoso, Ricardo Schiavo, Maria Sotiropoulos Padilla, Chenggu Wang, Sarah E Wolf, Hang Du, Kristy Guevara.

## Genome Informatics & Data Engineering

Jeffrey G Reid, Mona Nafde, Manan Goyal, George Mitra, Sanjay Sreeram, Rouel Lanche, Vrushali Mahajan, Sai Lakshmi Vasireddy, Gisu Eom, Krishna Pawan Punuru, Sujit Gokhale, Shehroze Aamer, Pooja Mule, Mudasar Sarwar, Muhammad Aqeel, Xiaodong Bai, Lance Zhang, Sean O’Keeffe, Razvan Panea, Evan Edelstein, Devika Torvi, Ayesha Rasool, William Salerno, Evan K Maxwell, Boris Boutkov, Alexander Gorovits, Ju Guan, Alicia Hawes, Olga Krasheninina, Samantha Zarate, Adam J Mansfield, Lukas Habegger, Stephen Tahan, Naveen Karumuri.

## Analytical Genetics and Data Science

Gonc□alo Abecasis, Manuel Allen Revez Ferreira, Joshua Backman, Kathryn Burch, Adrian Campos, Liron Ganel, Sheila Gaynor, Benjamin Geraghty, Arkopravo Ghosh, Christopher Gillies, Lauren Gurski, Eric Jorgenson, Tyler Joseph, Michael Kessler, Jack Kosmicki, Adam Locke, Priyanka Nakka, Jonathan Marchini, Karl Landheer, Olivier Delaneau, Maya Ghoussaini, Anthony Marcketta, Joelle Mbatchou, Jonathan Ross, Carlo Sidore, Eli Stahl, Timothy Thornton, Rujin Wang, Kuan-Han Wu, Bin Ye, Blair Zhang, Andrey Ziyatdinov, Yuxin Zou, Jingning Zhang, Kyoko Watanabe, Mira Tang, Frank Wendt, Suganthi Balasubramanian, Suying Bao, Kathie Sun, Chuanyi Zhang, Sean Yu, Aaron Zhang, David Corrigan, Dhruv Shidhaye, Chen Wang, Keyrun Adhikari, Alexander Lachmann, Anna Alkelai, Mark Weiner, Julian Stamp, Joseph Pickrell, Ammar Tareen.

## Therapeutic Area Genetics

Adolfo Ferrando, Giovanni Coppola, Luca A. Lotta, Alan Shuldiner, Katherine Siminovitch, Brian Hobbs, Jon Silver, William Palmer, Rita Guerreiro, Amit Joshi, Antoine Baldassari, Cristen Willer, Sarah Graham, Ernst Mayerhofer, Erola Pairo Castineira, Mary Haas, Niek Verweij, George Hindy, Jonas Bovijn, Tanima De, Luanluan Sun, Olukayode Sosina, Arthur Gilly, Peter Dornbos, Moeen Riaz, Manav Kapoor, Gannie Tzoneva, Veera Rajagopal, Sahar Gelfman, Vijay Kumar, Jacqueline Otto, Jose Bras, Silvia Alvarez, Jessie Brown, Hossein Khiabanian, Joana Revez, Kimberly Skead, Jae Soon Sul, Lei Chen, Sam Choi, Amy Damask, Nan Lin, Charles Paulding, Sameer Malhotra, Joseph Herman, Jacob McPadden, David Blair, Joshua Motelow, Julie Horowitz, Wenmin Zhang.

## Research Program Management & Strategic Initiatives

Marcus B Jones, Michelle G LeBlanc, Nadia Rana, Jennifer Rico-Varela, Jaimee Hernandez, Larizbeth Romero, Ashley Paynter.

## Senior Partnerships & Business Operations

Randi Schwartz, Lourdes Crane, Alison Fenney, Jody Hankins, Anna Han, Samuel Hart, Ryan Smith, Sarah Murphy.

## Business Operations & Administrative Coordinators

Ann Perez-Beals, Gina Solari, Johannie Rivera-Picart, Michelle Pagan, Sunilbe Siceron.

Affiliations:

^2^Regeneron Genetics Center, Tarrytown, NY, USA.

^3^Contact information

## Acknowledgements

Biosamples and data used in this work were generously provided by the participants in the Enroll-HD and REGISTRY study and made available by CHDI Foundation, Inc. Enroll-HD is a clinical research platform and longitudinal observational study for Huntington’s disease families intended to accelerate progress towards therapeutics; it is sponsored by CHDI Foundation, a nonprofit biomedical research organization exclusively dedicated to collaboratively developing therapeutics for HD. Enroll-HD and the previous contributing REGISTRY study from the European Huntington Disease Network would not be possible without the vital contribution of the research participants and their families. We also thank those individuals who contributed to the collection of the Enroll-HD data, listed at https://enroll-hd.org/enrollhd_documents/Enroll-HD_Acknowledgement-List_20250821.pdf. Lastly, we would like to thank Drs. James F. Gusella and Docia Demmin for critical review of the manuscript and helpful feedback, and Dr. Simon Noble for editorial input and assistance in improving the manuscript.

## Author Contributions

TFV, JR, and SK initiated the proposal and sample approval for WGS of Enroll-HD in collaboration with Regeneron. Regeneron performed the WGS and initial genetic data processing (sequence alignment and variant callings) and QC. QSL performed the data analysis and wrote the first draft of the manuscript. All authors reviewed, contributed, and approved the final submitted draft of manuscript.

