## Supplementary Note for "Genetic modifiers of psychiatric, motor, and cognitive symptoms in Huntington’s disease"

### Supplemental Methods

#### Variant annotations

Missense variants were assigned an aggregated predicted deleteriousness score by a weighted sum of deleteriousness predictions from 40+ algorithms precomputed in dbNSFP version 4.7a ^1^ including AlphaMissense ^2^ and ESM1b ^3^, but also includes other deep learning models such as PrimateAI ^4^, EVE ^5^, DANN ^6^, LINSIGHT ^7^, GenoCanyon ^8^, VARITY ^9^, MetaRNN ^10^, classical ML such as REVEL ^11^, ClinPred ^12^, DEOGEN2 ^13^, BayesDel ^14^, MetaLR ^15^, MetaSVM ^15^, FATHMM ^16^, PROVEAN ^17^, MutationAssessor ^18^, MutationTaster ^19^, M‑CAP ^20^, PolyPhen2 ^21^, evolutionary / constraint algorithms such as GERP++ ^22^, phyloP ^23^, phastCons ^24^, SiPhy ^25^, bStatistic ^26^, fitCons ^27^, Eigen ^28^, CADD (hybrid) ^29,30^, and rule‑based / clinical algorithm such as ClinVar ^31,32^.

An aggregated score (del_pred_weighted, also used as a weight in rare variant gene-based analysis) is derived using the deleteriousness cutoff and weight of different algorithms so that algorithms from the same algorithm family are not overweight.

ann <- ann %>%

rowwise() %>%

mutate(del_pred_weighted = sum(GERP.._RS>=2, AlphaMissense_pred %in% c("A"), CADD_phred >= 15, Eigen.phred_coding >= 15, GenoCanyon_score >= 0.7, integrated_fitCons_score >= 0.7,

DANN_score >= 0.9, LINSIGHT >= 0.7, phyloP100way_vertebrate >= 2, phyloP470way_mammalian >= 2, phyloP17way_primate >= 1.5,

phastCons100way_vertebrate >= 0.7, phastCons470way_mammalian >= 0.7, phastCons17way_primate >= 0.7, SiPhy_29way_pi>= 0.8,

bStatistic >= 0.7,

CLNSIG2 %in% c("Pathogenic/Likely_pathogenic"),

LIST.S2_pred %in% c("D"), ESM1b_pred %in% c("D"), ClinPred_pred %in% c("D"),

EVE_score >= 0.7, DEOGEN2_pred %in% c("D"),

PrimateAI_pred %in% c("D"), MPC_score >=2, REVEL_score >= 0.75, MutPred_score >= 0.75,

M.CAP_pred %in% c("D"), MetaRNN_pred %in% c("D"),

MetaLR_pred %in% c("D"), MetaSVM_pred %in% c("D"),

PROVEAN_pred %in% c("D"), FATHMM_pred %in% c("D"),

MutationAssessor_pred %in% c("M", "H"),

MutationTaster_pred %in% c("D","A"), LRT_pred %in% c("D"), na.rm= TRUE) +

sum(VARITY_R_score >= 0.7, VARITY_ER_score >= 0.7, VARITY_R_LOO_score >= 0.7,

VARITY_ER_LOO_score >= 0.7)/4 +

sum(SIFT_pred %in% c("D"), SIFT4G_pred %in% c("D"))/2 +

sum(BayesDel_noAF_pred %in% c("D"),BayesDel_addAF_pred %in% c("D"))/2 +

sum(Polyphen2_HDIV_pred %in% c("D"), Polyphen2_HVAR_pred %in% c("D"))/2 +

sum(fathmm.MKL_coding_pred %in% c("D"), fathmm.XF_coding_pred %in% c("D"))/2)

#### Network-based gene prioritization (NetWAS)

To further assess functional coherence of genetic associations, we performed network-based gene reprioritization using NetWAS^33^. Gene-level P-values derived from GWAS using Multi-marker Analysis of GenoMic Annotation (MAGMA v1.10)^34^ as implemented in Functional Mapping and Annotation (FUMA v1.8.3)^35^ were used as input, and genes with nominal association (p < 10^-4^) were defined as positive examples. NetWAS was run using the HumanBase (GIANT) brain and basal ganglion tissue-specific network, and genes were ranked based on their network connectivity to nominally associated genes. Resulting top 50 ranked gene lists were used for over-representation analysis using https://www.gsea-msigdb.org/gsea/msigdb/human/annotate.jsp and for downstream interpretation.

### Supplemental Results

#### Clinical characteristics of the Enroll-HD cohort

Informant’s judgement of initial major symptoms is largely consistent between rater, family members, and the study participants, with 50-60% of participants reporting motor ~20% of participants reporting psychiatric symptoms, and ~7-11% reporting cognitive symptoms, and ~10-18% reporting mixed symptoms being the initial major symptom (**Supplementary Table S39**).

**Supplementary Table 39**. Clinical characteristics of Enroll-HD cohort

| N (%) | motor | cognitive | psychiatric | oculomotor | other | Mixed |
| --- | --- | --- | --- | --- | --- | --- |
| Rater | 9047 (53.9%) | 1252 (7.5%) | 3275 (19.5%) | 34 (0.2%) | 70 (0.4%) | 3111 (18.5%) |
| Family | 9027 (56.4%) | 1346 (8.4%) | 3484 (21.8%) | 26 (0.2%) | 146 (0.9%) | 1964 (12.3%) |
| Subject | 9802 (58.7%) | 1850 (11.1%) | 3146 (18.8%) | 26 (0.2%) | 213 (1.3%) | 1674 (10.0%) |

#### ExWAS results

Among the 7 genes passing study wide significant threshold (significant Cauchy test after multiple testing correction (*p* < 3.95x 10^-7^) and significant mask-level p-value <= 3.29 x 10^-8^ & minor allele count (MAC) >= 20) across 12 phenotypes (**Supplementary Tables 11 and 12**, **Figure 3,** **Supplementary Figures 6 and 7**), four (*FAN1*, *PMS1*, *POLD1*, and *NOP14* (**Supplementary Tables 13-16**, **Supplementary Figures 8-11**) genes overlapped with previously implicated HD modifier genes associated with age at various clinical landmarks and somatic instability. In this study, these genes were associated with multiple phenotypes and were deemed high confidence findings. Rare variants in *FAN1* were associated with time to motor, cognitive, apathy, and perseverative obsessive behaviour symptom onset, and time to HD diagnosis, while *PMS1* and *POLD1* were significantly associated with time to motor onset and time to age at HD diagnosis. *POLD1* was additionally associated with time to age at HD diagnosis. Significant *FAN1* association (Cauchy p-values 1.53x10^-30^ to 6.88x10^-15^ for time to motor onset) was not only driven by known modifiers R507H (15AM1^36^, p-value_SPA_ = 1.10x10^-11^, hazard ratio (HR) = 1.64) and R377W (15AM3^36^, p-value_SPA_ = 1.09x19^-8^, HR = 1.72), but also other rare SNVs (R145H, p-value_SPA_ = 9.14x10^-3^, HR = 1.56; and P654L, p-value_SPA_ = 1.15x10^-3^, HR = 2.85) and ultra-rare pLoF and missense variants (**Supplementary Table 13**). Significant *PMS1* association (Cauchy p-values 8.61x10^-11^ to 1.78x10^-4^ for time to motor onset) was driven by rare SNVs (T75I, p-value_SPA_ = 1.19x19^-8^, HR = 0.33; R26K, p-value_SPA_=8.45x10^-3^, HR = 0.86) and ultra-rare missense and pLoF variants (**Supplementary Table 14**). T75I was also previously reported to be associated with decreased expansion of chr19:14.8 Mb AAAG repeats^37^. Significant *POLD1* association (Cauchy p-values 1.68x10^-8^ to 1.11x10^-5^ for time to motor symptom onset) was driven by both rare SNVs (R30W, p-value_SPA_ = 1.84x19^-8^, HR = 0.73; Q710H, p-value_SPA_=2.19x10^-2^, HR = 2.05; R875H, p-value_SPA_=3.44x10^-2^, HR = 0.86) and ultra-rare missense and pLoF variants (**Supplementary Table 15**).

*NOP14* was associated with time to irritability symptom onset and time to age at HD diagnosis. Significant *NOP14* association (Cauchy p-values 2.85x10^-7^ to 4.28x10^-4^) was driven by rare variant R697C (p-value_SPA_ = 1.03x19^-7^, HR = 0.63), which tags the *HTT* non-canonical CAACAG-duplication^38^ (**Supplementary Table 16**). When including stop-loss variants as pLoF variants (as implemented in an earlier version of the analysis with full results not reported except *SH3BP2*), *SH3BP2* gene association was an additional high confidence finding (**Supplementary Table 17**). The variant X562W driving the association of *SH3BP2* association also tags the *HTT* non-canonical CAACAG-duplication (**Supplementary Table** **17 and Supplementary Figure 12**). The three MMR genes and *HTT* locus are identical to those reported in Gelfman et al., 2026^39^ with difference due to the analytical pipeline deployed.

Two additional genes in **Supplementary Table 11** were in segmental duplication/repeat regions and warrant a closer evaluation with respect to genotype quality and causal vs. passenger effect (*NBPF6* in **Supplementary Table 18**, **Supplementary Figure 13**, the other (*C16orf54*) deprioritized during manual review). An additional gene *AVPR1A* was associated with age of psychosis onset, which was primarily driven by a single rare coding variant F308L (hazard ratio ~ 2.75, p-value_SPA_ = 8.69 x 10^-7^) with few ultra rare variants boosting the gene-level association signal (**Supplementary Table 19**). *AVPR1A* encodes a vasopressin‑responsive GPCR that plays key roles in social behavior and stress regulation, with prominent relevance to neuropsychiatric traits^40^. Antagonist of the vasopressin 1a receptor has been studied for irritability and aggressive behavior in HD^41^.

Additional genes meeting a slightly relaxed study wide significant threshold (significant Cauchy test after multiple testing correction (*p* < 3.95x 10^-7^) and significant mask-level p-value <= 10^-6^ and MAC >= 5) across 12 phenotypes are also reported in **Supplementary Table 11**. Genes involved in the adjacent DNA damage response (DDR) such as *CHTF8* (Cauchy p-values ranging from 8.94x10^-8^ to 1.21x10^-6^, MAC=13, driven by 9 ultra-rare missense variants with alternative allele frequency (AAF) less than 0.001, **Supplementary Table 20, Supplementary Figure 14**) and *PSME4* (Cauchy p-value = 2.34x10^-8^, MAC = 7, driven by 6 ultra-rare pLoF variants with AAF less than 0.0001, **Supplementary Table 21, Supplementary Figure 15**) can be further evaluated in similar experiments interrogating the roles of MMR genes in somatic instability. *CHTF8* encodes a short protein that forms part of the Ctf18 replication factor C (RFC) complex. The heteroheptameric RFC complex plays a role in sister chromatid cohesion and may load the replication clamp proliferating cell nuclear antigen (PCNA) onto DNA during DNA replication and repair, while *PSME4* encodes a proteasome activator involved in chromatin-bound proteasome function and DNA damage response. Further validation requires an increase in sample size or direct studies evaluating gene knock-out or effects of missense variants in MMR pathway modulation. Additional themes emerged from this list of candidate modifiers include protein homeostasis, ubiquitin–proteasome system, and autophagy; ribosome biogenesis, RNA processing, and nucleolar stress; vesicle trafficking, synaptic function, and receptor signaling; immune / inflammatory signaling and cellular stress response; and lipid metabolism & membrane composition (**Supplementary Tables 11 and 12**). Detailed gene-level, variant-level (single variant or collapsed ultra-rare pseudo-marker) are presented for each gene in **Supplementary Tables 22-30** and **Supplementary Figures 16-25**.

A suggestive association signal in the Manhattan plot in both motor symptom onset and age at HD diagnosis was seen on chromosome 7 harboring the gene for *ZFAND2A* (Cauchy p-values 1.16x10^-6^ to 2.34x10^-6^, MAC = 4, **Supplementary Table 31, Supplementary Figure 26**). Interestingly, *ZFAND2A* encodes zinc finger AN1-type containing 2A and is predicted to be in nucleus and part of proteasome complex. It is induced by HSF1 responding to proteotoxic stress.

#### NetWAS network reprioritization results

Network-based reprioritization (NetWAS)^33^ integrates gene-level association statistics with tissue-specific gene interaction networks to prioritize genes based on both their statistical evidence and their functional connectivity. This approach enables identification of biologically coherent pathways and highlights genes that may not reach genome-wide significance individually but are supported through network context. NetWAS supported convergence of associated genes within DNA repair and repair-related pathways.

##### Brain-specific network

Top NetWAS-ranked genes using gene-level GWAS results of age of motor symptom onset included MMR genes (*MSH2* and *MLH1*), canonical DNA damage response pathways including *RAD50*, *BRCA1*, *FANCG*, and *ATM*, and replication / repair synthesis (*POLD1*, MCMs) (**Supplementary Table 35**). It is worth noting that *MSH2* did not reach genome wide significance in motor symptom onset GWAS but is a candidate gene for the genome wide significant association signal for somatic instability supported by both GWAS and gene KO experiment ^36,42^. Even though functional evidence indicates that canonical DDR signaling components such as ATM are not required for somatic expansion ^42^, a ginsenoside compound K (CK) was previously shown to have a protective effect in R6/2 HD mice model, via suppressing the activation of ATM/AMPK and reducing neuronal toxicity and mHTT aggregation^43^. The NetWAS signals may reflect the broader replication‑independent, repair‑associated replication‑like stress context in which mismatch repair–mediated expansion occurs, but it is essential to test the hypothesis in experimental systems.

Network-based analyses using a shared brain tissue framework revealed substantial convergence of both motor and cognitive phenotypes on replication-associated mismatch repair pathways, while still supporting domain-preferential modifier effects (**Supplementary Table 35**). Over-representation analysis results for top 50 NetWAS ranked genes are available in **Supplementary Table 40**.

##### Basal ganglion-specific network

NetWAS analysis of age of cognitive symptom onset highlights a distinct network enriched for transcriptional regulation, RNA processing, chromatin remodeling, and single-strand repair processes, including *PARP2* (**Supplementary Tables 35 and 40)** compared to using brain-specific netweork, suggesting that striatum, a major component of the basal ganglia and vulnerable region for HD patients, revealed the RNA processing aspect better than the generic brain-specific network. It is worth noting that *TCERG1* was a significant GWAS modifier for motor and not for cognitive onset yet. NetWAS analysis for motor and violent/aggressive behavior using basal ganglion-specific network likewise also identified RNA processing as a key converging theme among top 50 NetWAS ranked genes (**Supplementary Table 40**).

### Supplementary Tables

Suppl Table 1. Sample size and event rate for the GWAS and ExWAS cohorts

Suppl Table 2 Judgement of initial major symptoms

Suppl Table 3. Genome wide significant lead variants for 4 age of HD diagnosis traits

Suppl Table 4. GWAS meta-analysis results for 35 genome wide significant independent signals (linkage disequilibrium (LD) r^2^ < 0.1, corresponding to Supp Table 1) from 12 genomic regions across 11 traits (excluding sxsubj). Some are novel, while others overlap with previously reported signals.

Suppl Table 5. LD friends for 35 genome wide significant independent signals (linkage disequilibrium (LD) r^2^ < 0.1, corresponding to Supp Table 4) from 12 genomic regions across 11 traits (excluding sxsubj).

Suppl Table 6. GWAS meta-analysis results for all genome wide significant signals and additional variants of interest from 12 genomic regions across 11 traits (excluding sxsubj).

Suppl Table 7. GWAS meta-analysis results for genome wide significant signals previously reported by GeM-HD or Hujoel et al., 2026

Suppl Table 8. SNPs in the *MSH3* and *FAN1* super-enhancer regions reaching genome wide significance threshold

Suppl Table 9. Polygenic score (PGS) association with HD phenotype using Cox regression model adjusting for CAG length, sex, and the first two principal component representing population substructure.

Suppl Table 10. Polygenic score (PGS) association with HD phenotype using linear model adjusting for CAG length, sex, and the first two principal component representing population substructure.

Suppl Table 11. Summary of ExWAS rare variant gene-level test results from time‑to‑event mixed‑effects survival (frailty) models evaluating genetic modifiers of symptom onset and/or HD diagnosis.

Suppl Table 12. ExWAS rare variant gene-level test results from time‑to‑event mixed‑effects survival (frailty) models evaluating genetic modifiers of symptom onset and HD diagnosis. Both mask-level association results and omnibus Cauchy test are reported. The filter criteria are that one of the six tests (3 tests and 2 weighting schemes) in a given gene had Cauchy p-value < 0.05/(2*3*# of genes tested ~ 3.95x 10-7) and at least one of the masks within the gene had p-value less than 0.000001 and minimum MAC ≥ 5. Pvalue is SKAT-O results.

Suppl Table 13. *FAN1* ExWAS association results for motor symptom onset (ccmtrage)

Suppl Table 14. *PMS1* ExWAS association results for motor symptom onset (ccmtrage)

Suppl Table 15. *POLD1* ExWAS association results for motor symptom onset (ccmtrage)

Suppl Table 16. *NOP14* ExWAS association results for time to age at HD diagnosis (hddiagn)

Suppl Table 17. *SH3BP2* ExWAS association results for motor symptom onset (ccmtrage)

Suppl Table 18. *NBPF6* ExWAS association results for cognitive symptom onset (cccogage)

Suppl Table 19. *AVPR1*A ExWAS association results for psychosis symptom onset (ccpsyage)

Suppl Table 20. *CHTF8* ExWAS association results for onset of irritability symptom onset (ccirbage)

Suppl Table 21. *PSME4* ExWAS association results for onset of irritability symptom onset (ccirbage)

Suppl Table 22. *DGAT2* ExWAS association results for age of depression symptom onset (ccdepage)

Suppl Table 23. *LY6L* ExWAS association results for age of violent and aggressive behavior symptom onset (ccvabage)

Suppl Table 24. *NKG7* ExWAS association results for age of HD diagnosis (sxrater)

Suppl Table 25. *BIN1* ExWAS association results for onset of depression symptom onset (ccdepage)

Suppl Table 26. *TMEM171* ExWAS association results for onset of irritability symptom onset (ccirbage)

Suppl Table 27. *TMED1* ExWAS association results for onset of apathy symptom onset (ccaptage)

Suppl Table 28. *SPATA25* ExWAS association results for onset of apathy symptom onset (ccaptage)

Suppl Table 29. *PTCHD4* ExWAS association results for age of violent and aggressive behavior symptom onset (ccvabage)

Suppl Table 30. *DUSP8* ExWAS association results for age of apathy symptom onset (ccaptage)

Suppl Table 31. *ZFAND2A* ExWAS association results for age of HD diagnosis (hddiagn)

Suppl Table 32. MAGMA pathway enrichment analysis for time to motor symptom onset GWAS meta-analysis (Top 64)

Suppl Table 33. Gene set enrichment analysis against the Human Molecular Signatures Database (MSigDB)_v2025.1.Hs c2cp.kegg (canonical pathway KEGG) for time to motor symptom onset ExWAS using SKAT-O results (minimal p-value across all masks is used as gene-level p-value) signed -log(p) is used to rank the genes (Top 30)

Suppl Table 34. *MSH3* ExWAS association results for age of motor symptom onset (ccmtrage)

Suppl Table 35. Top 50 NetWAS-ranked genes for age of onset for motor/cognitive/one representative psychiatric symptoms

Suppl Table 36. ExWAS association results for selected MMR and DDR genes across 12 phenotypes with at least one association with nominal association p-value less than 0.05

Suppl Table 37. ExWAS association full results for selected MMR and DDR genes across 12 phenotypes

Suppl Table 38. References for summary statistics used in PGS analyses

Suppl Table 39. Clinical characteristics of Enroll-HD cohort

Suppl Table 40. Over-represented gene sets for top 50 NetWAS-ranked genes
