## Supplementary Figures for "Genetic modifiers of psychiatric, motor, and cognitive symptoms in Huntington’s disease"

### Slide 1
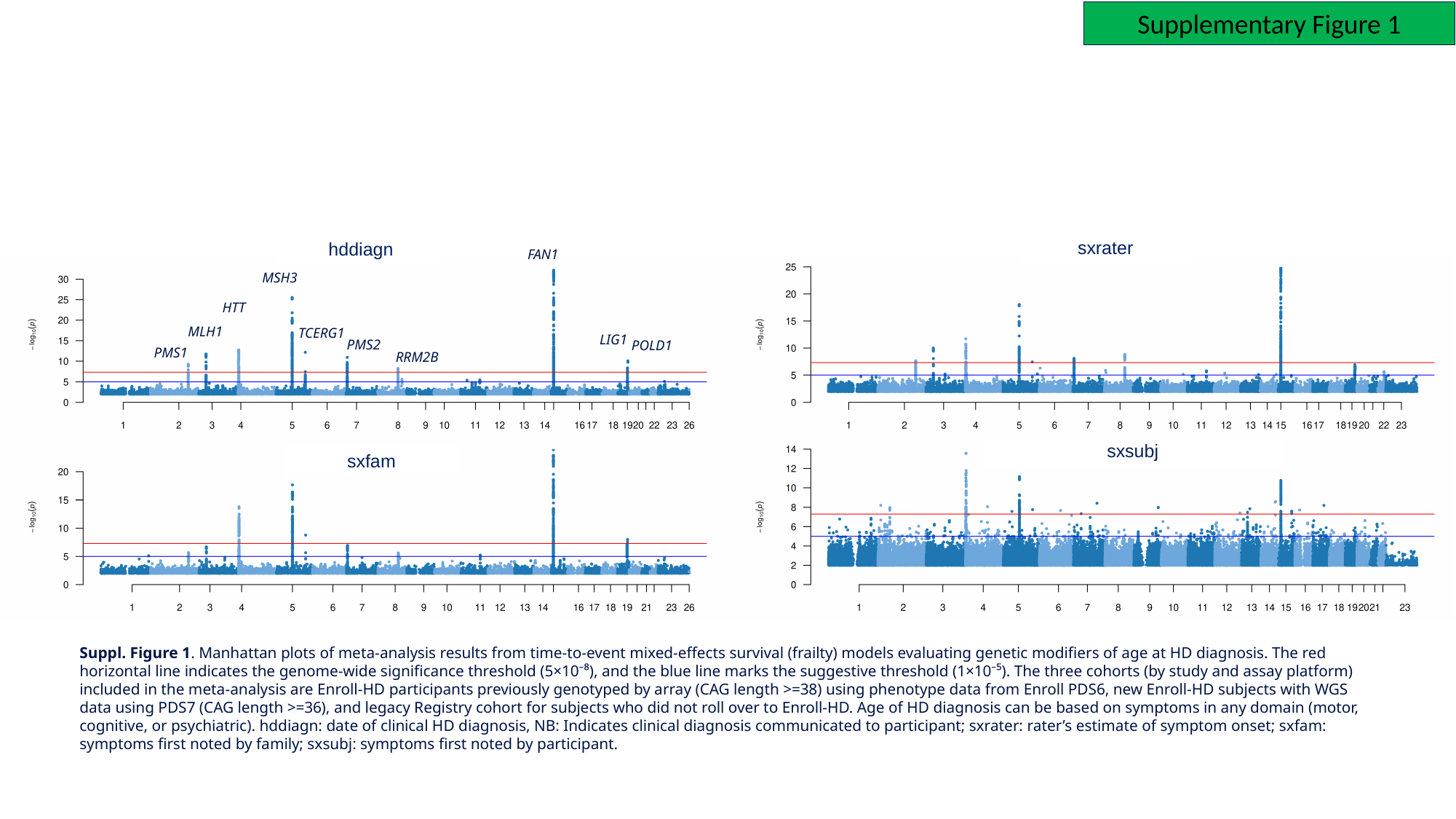

Supplementary Figure 1
sxrater
hddiagn
FAN1
MSH3
HTT
MLH1
TCERG1
LIG1
PMS2
POLD1
PMS1
RRM2B
sxsubj
sxfam
Suppl. Figure 1. Manhattan plots of meta-analysis results from time‑to‑event mixed‑effects survival (frailty) models evaluating genetic modifiers of age at HD diagnosis. The red horizontal line indicates the genome‑wide significance threshold (5×10⁻⁸), and the blue line marks the suggestive threshold (1×10⁻⁵). The three cohorts (by study and assay platform) included in the meta-analysis are Enroll-HD participants previously genotyped by array (CAG length >=38) using phenotype data from Enroll PDS6, new Enroll-HD subjects with WGS data using PDS7 (CAG length >=36), and legacy Registry cohort for subjects who did not roll over to Enroll-HD. Age of HD diagnosis can be based on symptoms in any domain (motor, cognitive, or psychiatric). hddiagn: date of clinical HD diagnosis, NB: Indicates clinical diagnosis communicated to participant; sxrater: rater’s estimate of symptom onset; sxfam: symptoms first noted by family; sxsubj: symptoms first noted by participant.

### Slide 2
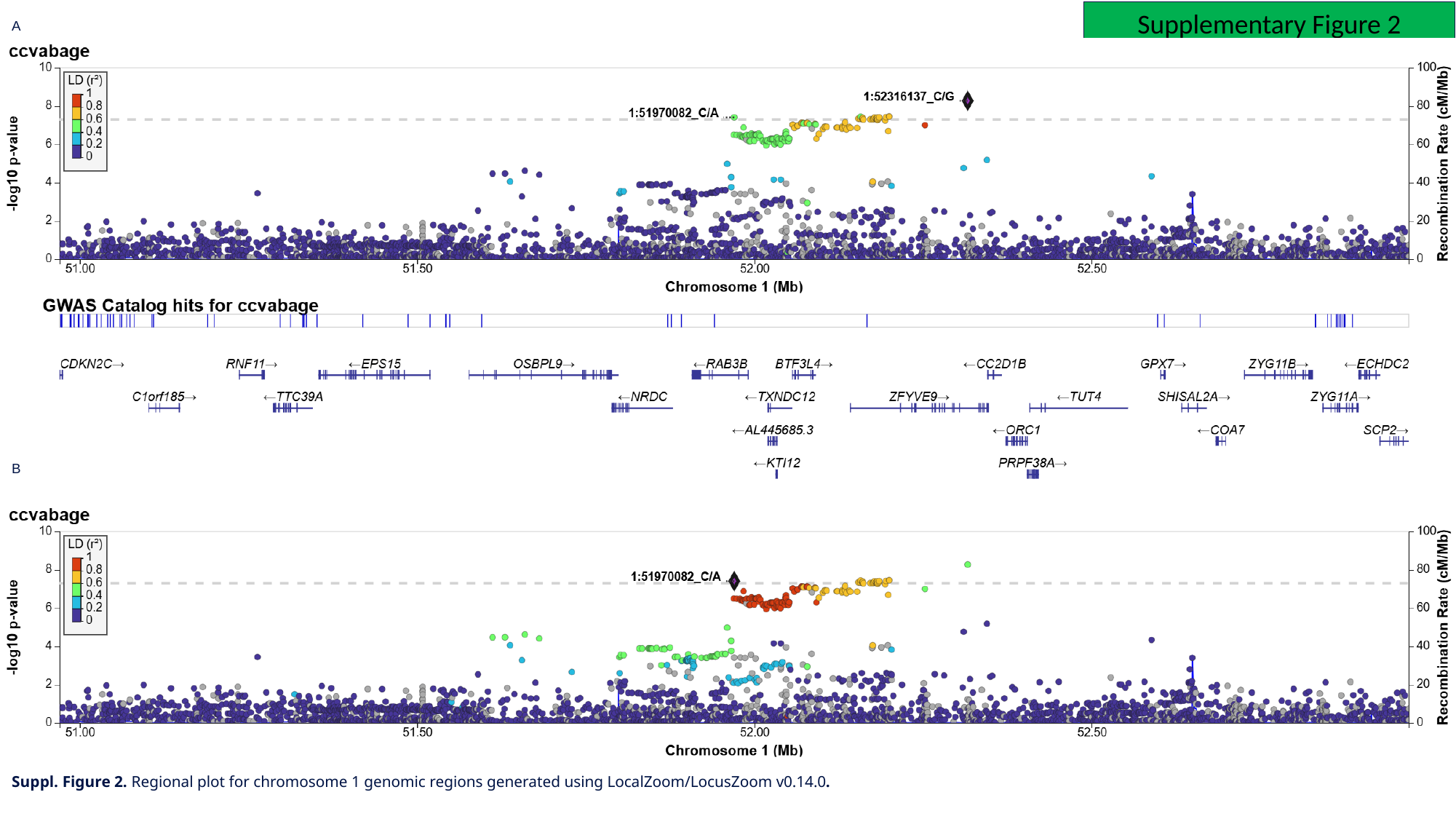

Supplementary Figure 2
A
B
Suppl. Figure 2. Regional plot for chromosome 1 genomic regions generated using LocalZoom/LocusZoom v0.14.0.

### Slide 3
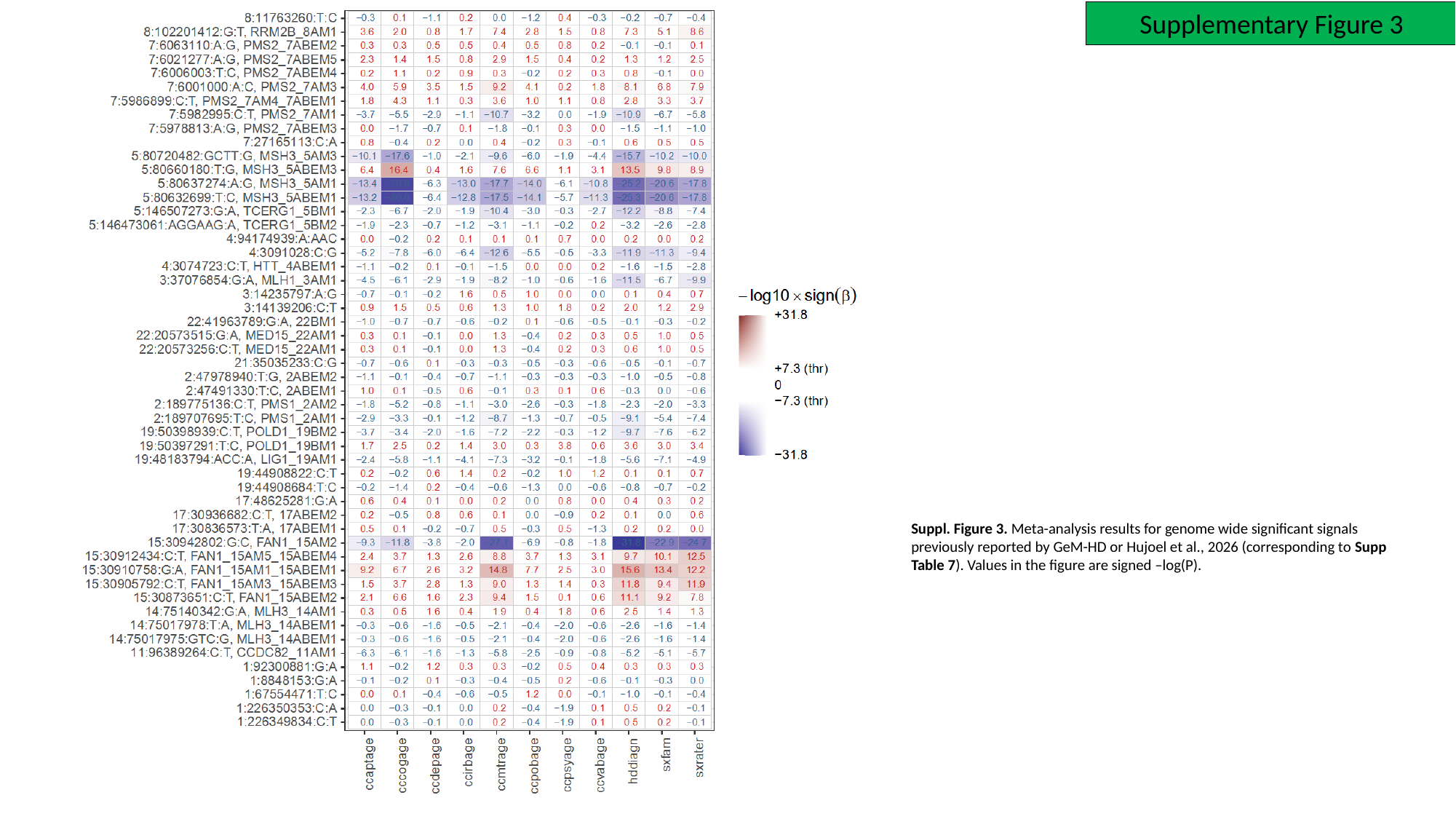

Supplementary Figure 3
Suppl. Figure 3. Meta-analysis results for genome wide significant signals previously reported by GeM-HD or Hujoel et al., 2026 (corresponding to Supp Table 7). Values in the figure are signed –log(P).

### Slide 4
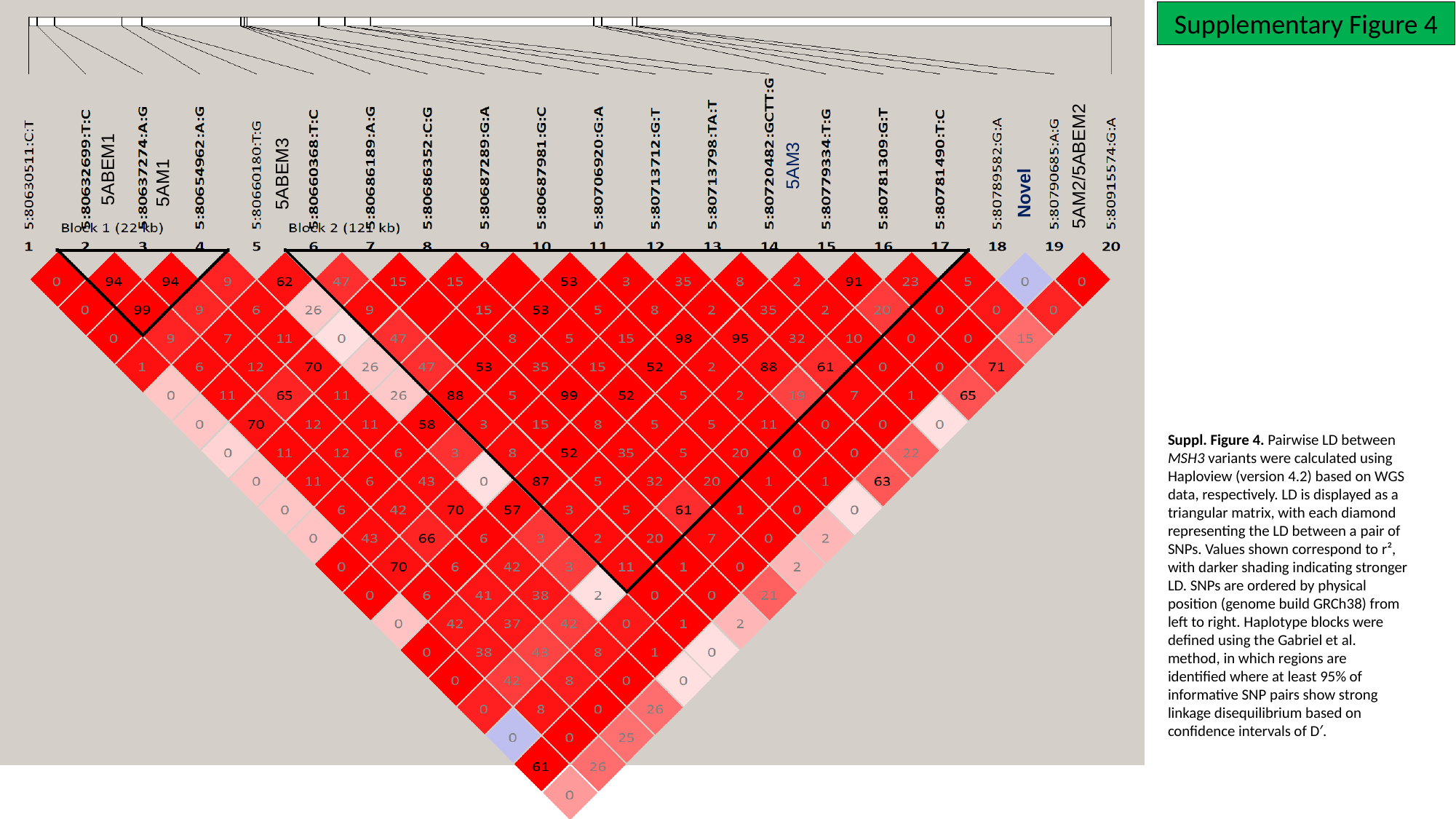

Supplementary Figure 4
Novel
5AM2/5ABEM2
5AM3
5ABEM1
5AM1
5ABEM3
Suppl. Figure 4. Pairwise LD between MSH3 variants were calculated using Haploview (version 4.2) based on WGS data, respectively. LD is displayed as a triangular matrix, with each diamond representing the LD between a pair of SNPs. Values shown correspond to r², with darker shading indicating stronger LD. SNPs are ordered by physical position (genome build GRCh38) from left to right. Haplotype blocks were defined using the Gabriel et al. method, in which regions are identified where at least 95% of informative SNP pairs show strong linkage disequilibrium based on confidence intervals of D′.

### Slide 5
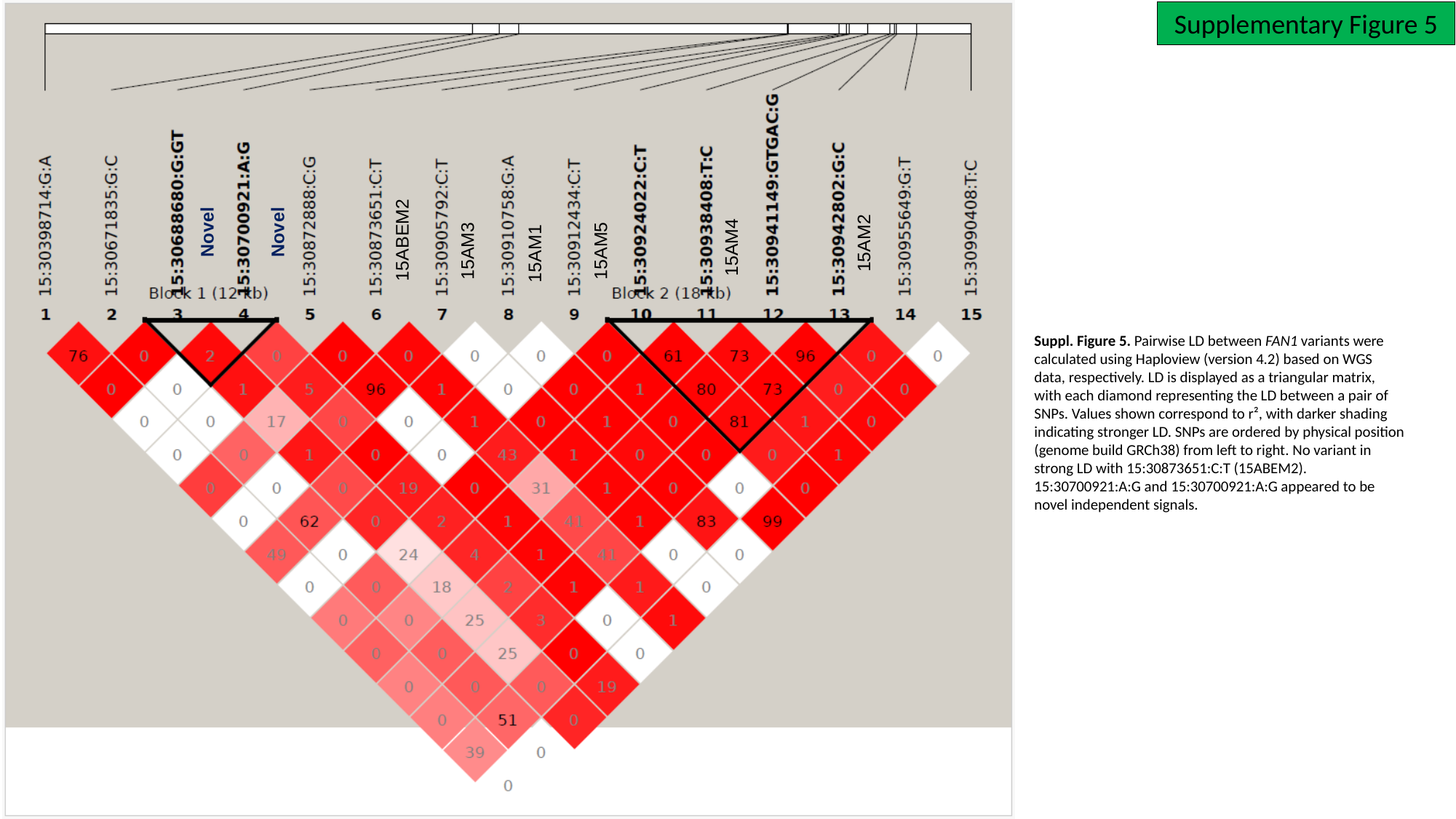

Supplementary Figure 5
Novel
Novel
15ABEM2
15AM2
15AM4
15AM5
15AM3
15AM1
Suppl. Figure 5. Pairwise LD between FAN1 variants were calculated using Haploview (version 4.2) based on WGS data, respectively. LD is displayed as a triangular matrix, with each diamond representing the LD between a pair of SNPs. Values shown correspond to r², with darker shading indicating stronger LD. SNPs are ordered by physical position (genome build GRCh38) from left to right. No variant in strong LD with 15:30873651:C:T (15ABEM2). 15:30700921:A:G and 15:30700921:A:G appeared to be novel independent signals.

### Slide 6
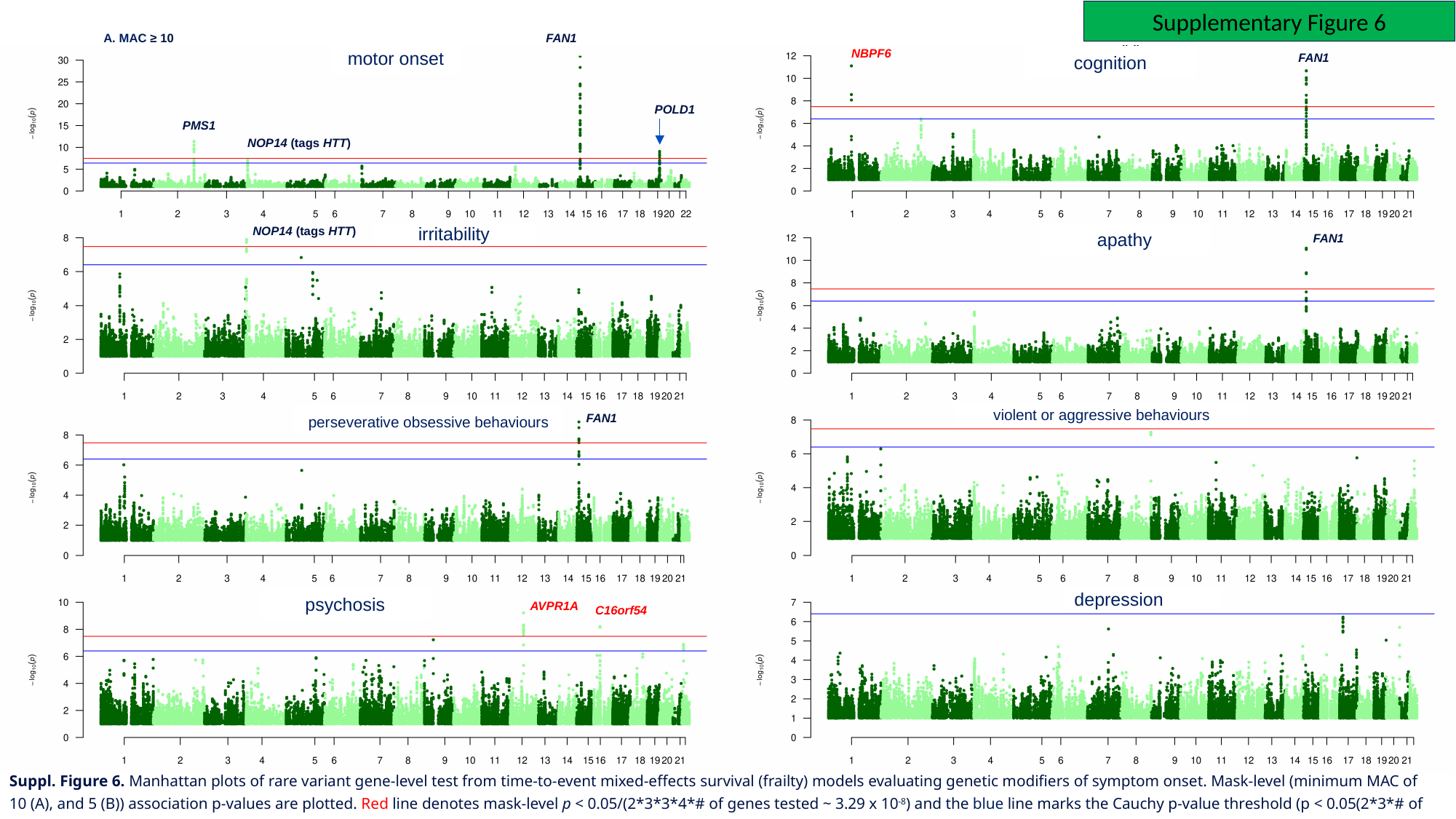

Supplementary Figure 6
A. MAC ≥ 10
FAN1
NBPF6
motor onset
FAN1
cognition
POLD1
PMS1
NOP14 (tags HTT)
NOP14 (tags HTT)
irritability
apathy
FAN1
violent or aggressive behaviours
perseverative obsessive behaviours
FAN1
psychosis
depression
AVPR1A
C16orf54
Suppl. Figure 6. Manhattan plots of rare variant gene-level test from time‑to‑event mixed‑effects survival (frailty) models evaluating genetic modifiers of symptom onset. Mask-level (minimum MAC of 10 (A), and 5 (B)) association p-values are plotted. Red line denotes mask-level p < 0.05/(2*3*3*4*# of genes tested ~ 3.29 x 10^-8^) and the blue line marks the Cauchy p-value threshold (p < 0.05(2*3*# of genes tested).

### Slide 7
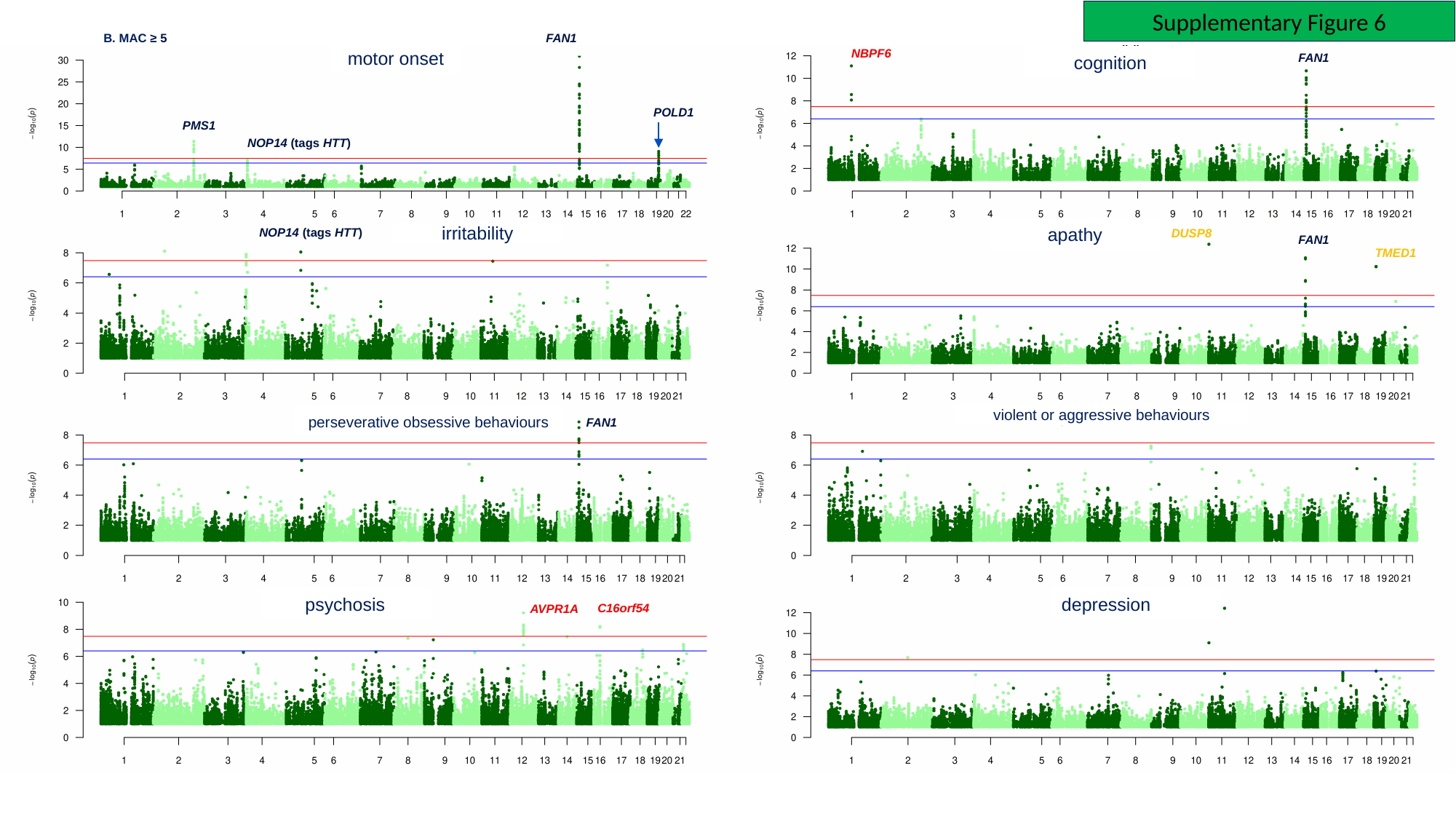

Supplementary Figure 6
B. MAC ≥ 5
FAN1
NBPF6
motor onset
FAN1
cognition
POLD1
PMS1
NOP14 (tags HTT)
apathy
NOP14 (tags HTT)
DUSP8
irritability
FAN1
TMED1
violent or aggressive behaviours
perseverative obsessive behaviours
FAN1
psychosis
depression
C16orf54
AVPR1A

### Slide 8
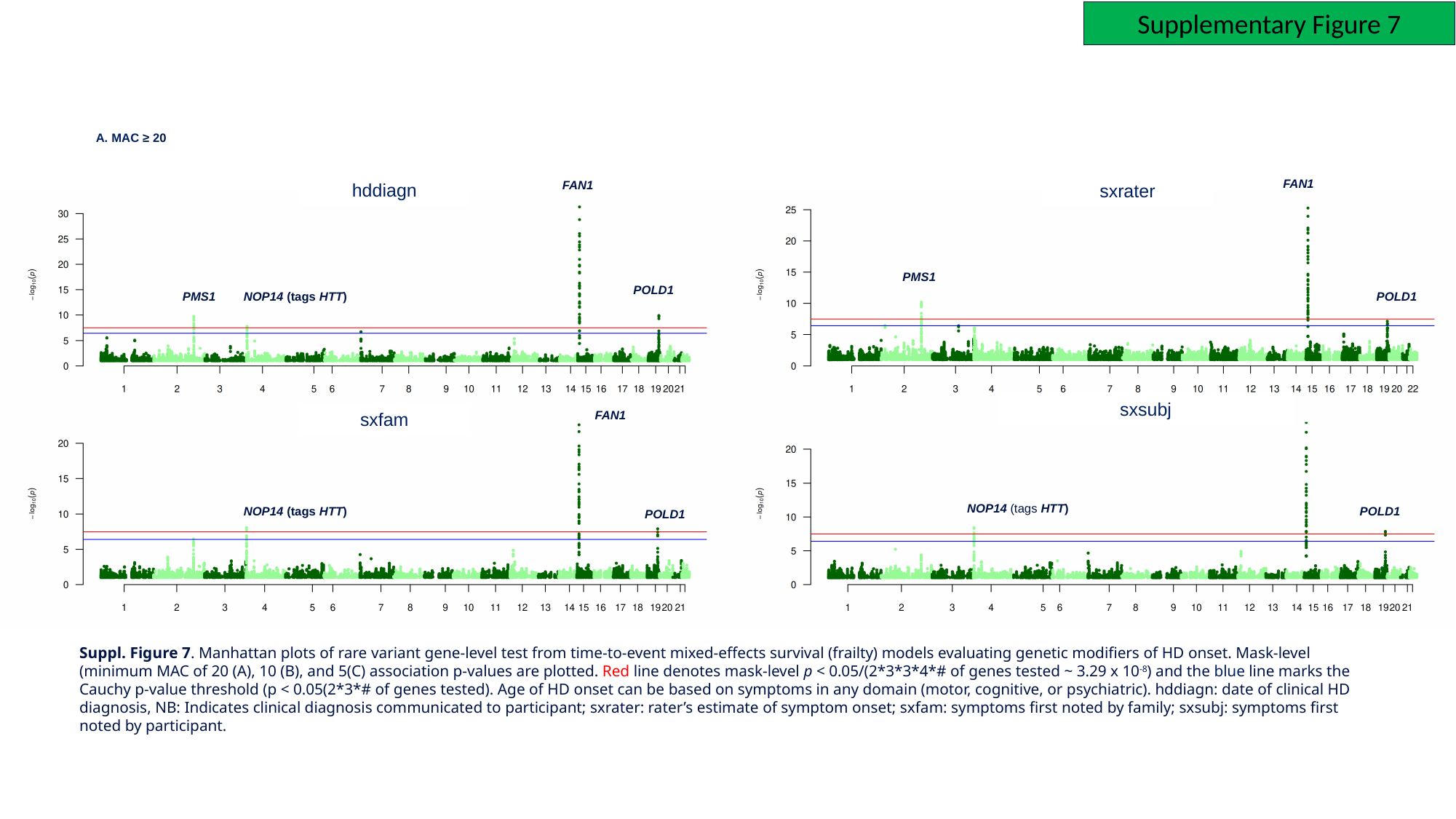

Supplementary Figure 7
A. MAC ≥ 20
FAN1
FAN1
hddiagn
sxrater
PMS1
POLD1
PMS1
NOP14 (tags HTT)
POLD1
sxsubj
FAN1
sxfam
NOP14 (tags HTT)
NOP14 (tags HTT)
POLD1
POLD1
Suppl. Figure 7. Manhattan plots of rare variant gene-level test from time‑to‑event mixed‑effects survival (frailty) models evaluating genetic modifiers of HD onset. Mask-level (minimum MAC of 20 (A), 10 (B), and 5(C) association p-values are plotted. Red line denotes mask-level p < 0.05/(2*3*3*4*# of genes tested ~ 3.29 x 10^-8^) and the blue line marks the Cauchy p-value threshold (p < 0.05(2*3*# of genes tested). Age of HD onset can be based on symptoms in any domain (motor, cognitive, or psychiatric). hddiagn: date of clinical HD diagnosis, NB: Indicates clinical diagnosis communicated to participant; sxrater: rater’s estimate of symptom onset; sxfam: symptoms first noted by family; sxsubj: symptoms first noted by participant.

### Slide 9
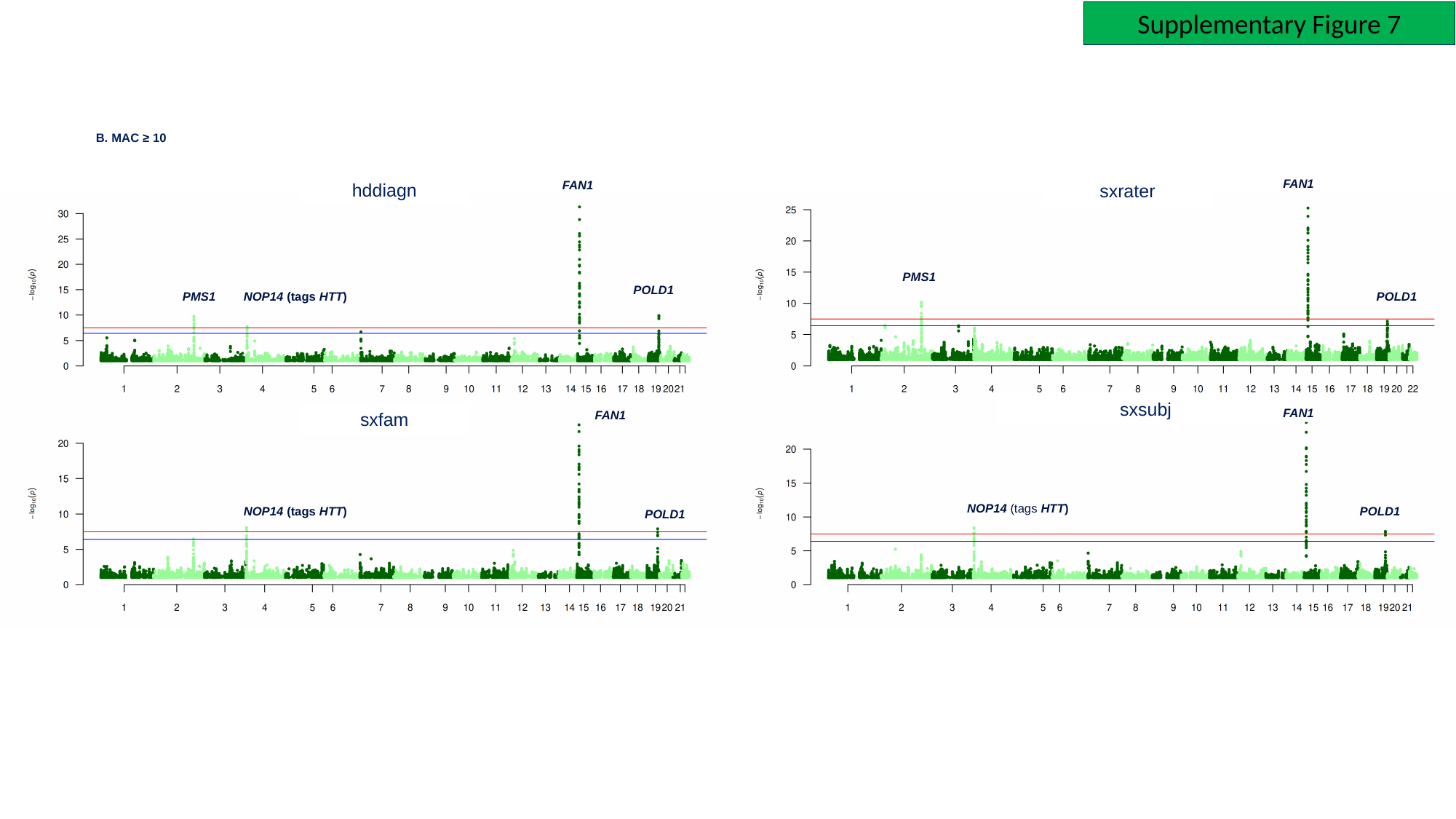

Supplementary Figure 7
B. MAC ≥ 10
FAN1
FAN1
hddiagn
sxrater
PMS1
POLD1
PMS1
NOP14 (tags HTT)
POLD1
sxsubj
FAN1
FAN1
sxfam
NOP14 (tags HTT)
NOP14 (tags HTT)
POLD1
POLD1

### Slide 10
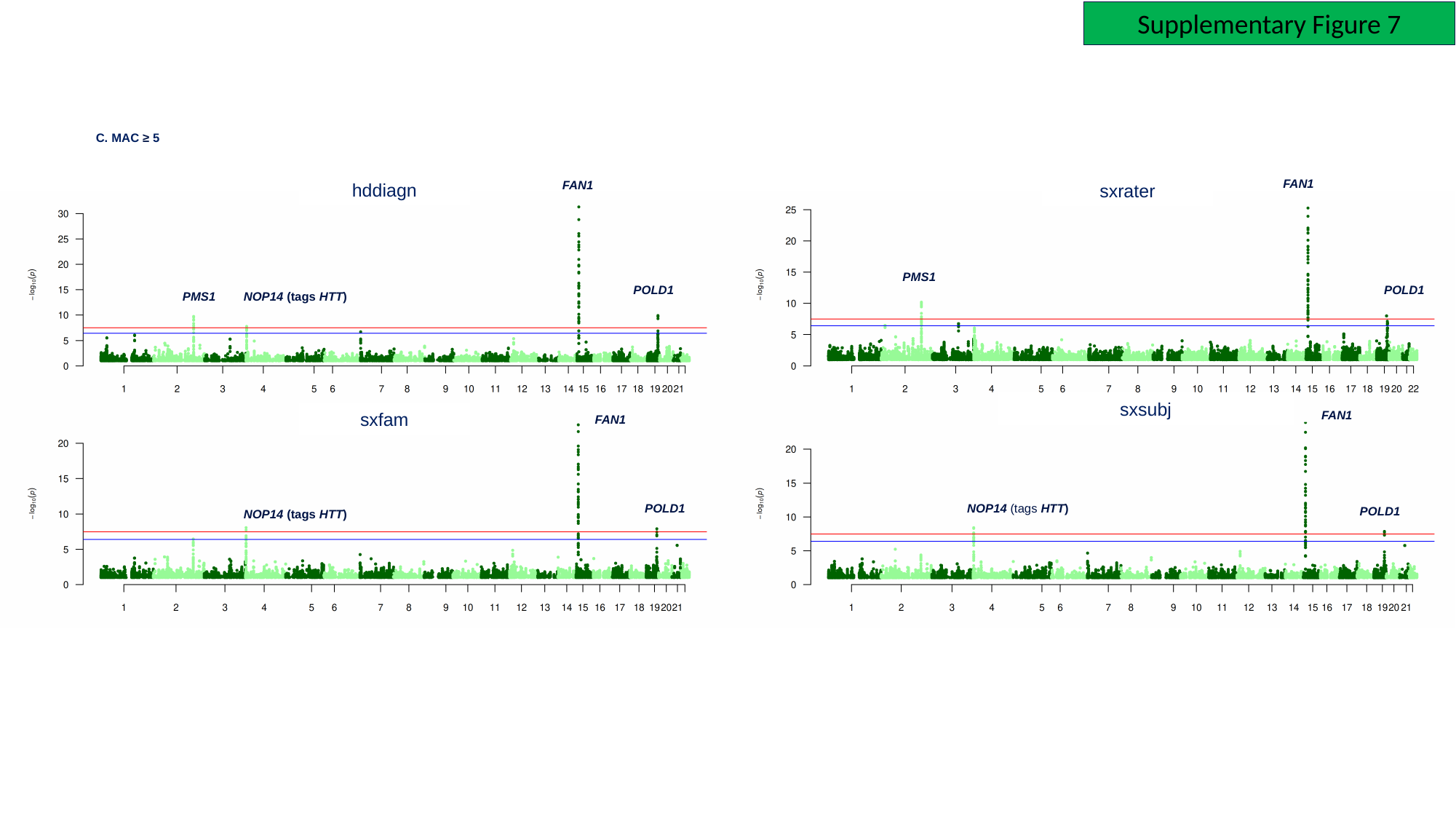

Supplementary Figure 7
C. MAC ≥ 5
FAN1
FAN1
hddiagn
sxrater
PMS1
POLD1
POLD1
PMS1
NOP14 (tags HTT)
sxsubj
FAN1
sxfam
FAN1
POLD1
NOP14 (tags HTT)
POLD1
NOP14 (tags HTT)

### Slide 11
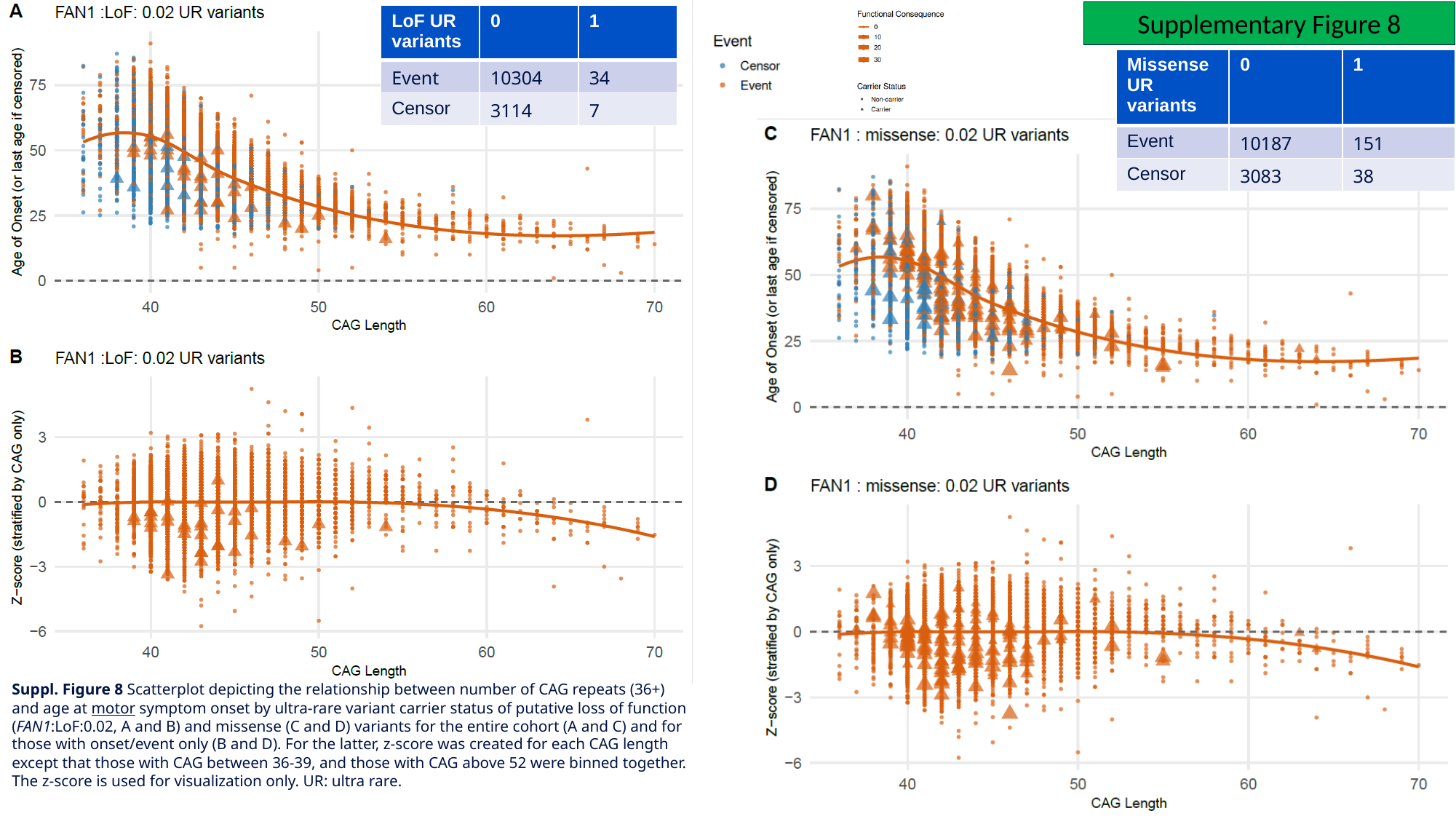

Supplementary Figure 8
| LoF UR variants | 0 | 1 |
| --- | --- | --- |
| Event | 10304 | 34 |
| Censor | 3114 | 7 |
| Missense UR variants | 0 | 1 |
| --- | --- | --- |
| Event | 10187 | 151 |
| Censor | 3083 | 38 |
Suppl. Figure 8 Scatterplot depicting the relationship between number of CAG repeats (36+) and age at motor symptom onset by ultra-rare variant carrier status of putative loss of function (FAN1:LoF:0.02, A and B) and missense (C and D) variants for the entire cohort (A and C) and for those with onset/event only (B and D). For the latter, z-score was created for each CAG length except that those with CAG between 36-39, and those with CAG above 52 were binned together. The z-score is used for visualization only. UR: ultra rare.

### Slide 12
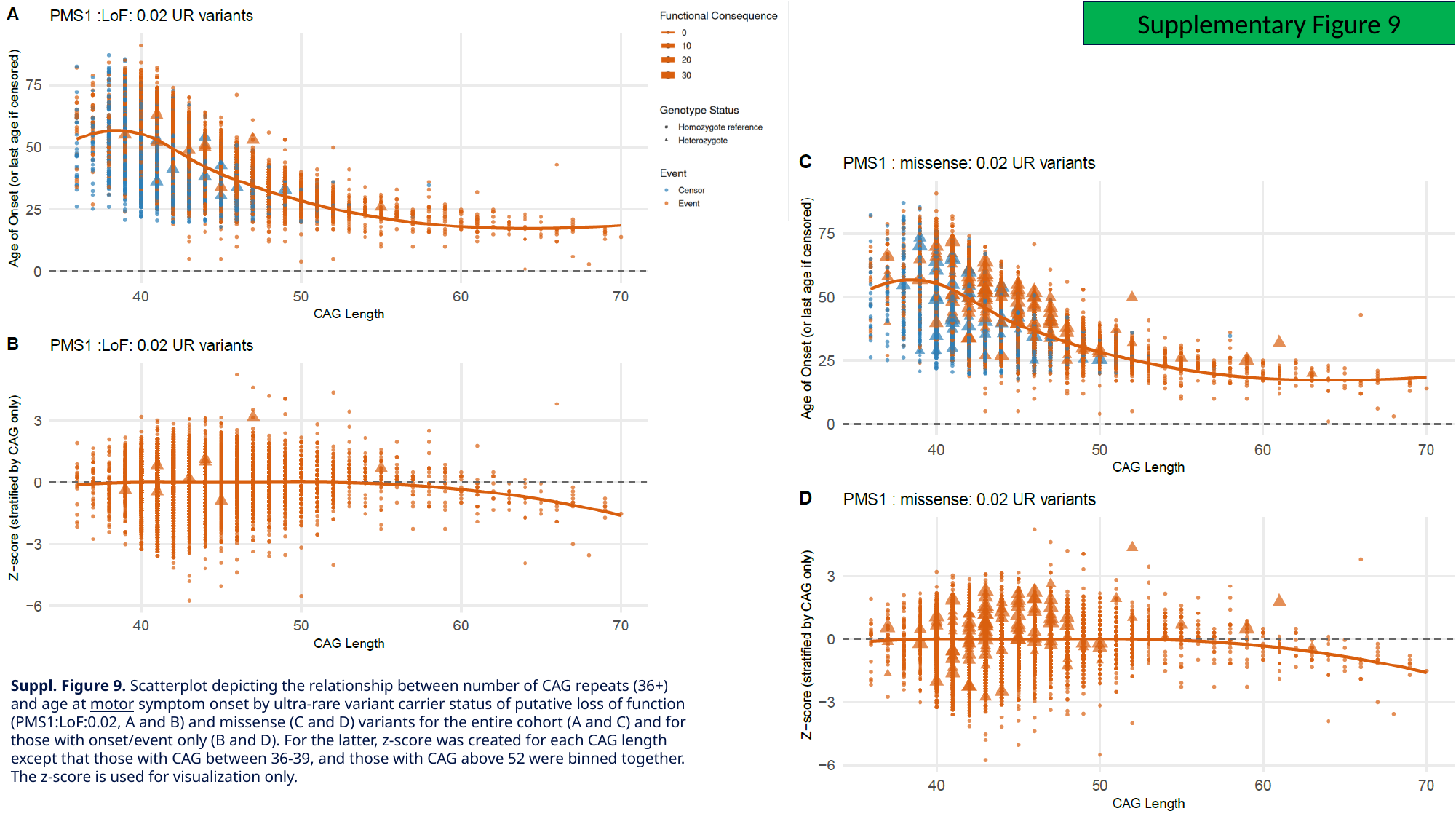

Supplementary Figure 9
Suppl. Figure 9. Scatterplot depicting the relationship between number of CAG repeats (36+) and age at motor symptom onset by ultra-rare variant carrier status of putative loss of function (PMS1:LoF:0.02, A and B) and missense (C and D) variants for the entire cohort (A and C) and for those with onset/event only (B and D). For the latter, z-score was created for each CAG length except that those with CAG between 36-39, and those with CAG above 52 were binned together. The z-score is used for visualization only.

### Slide 13
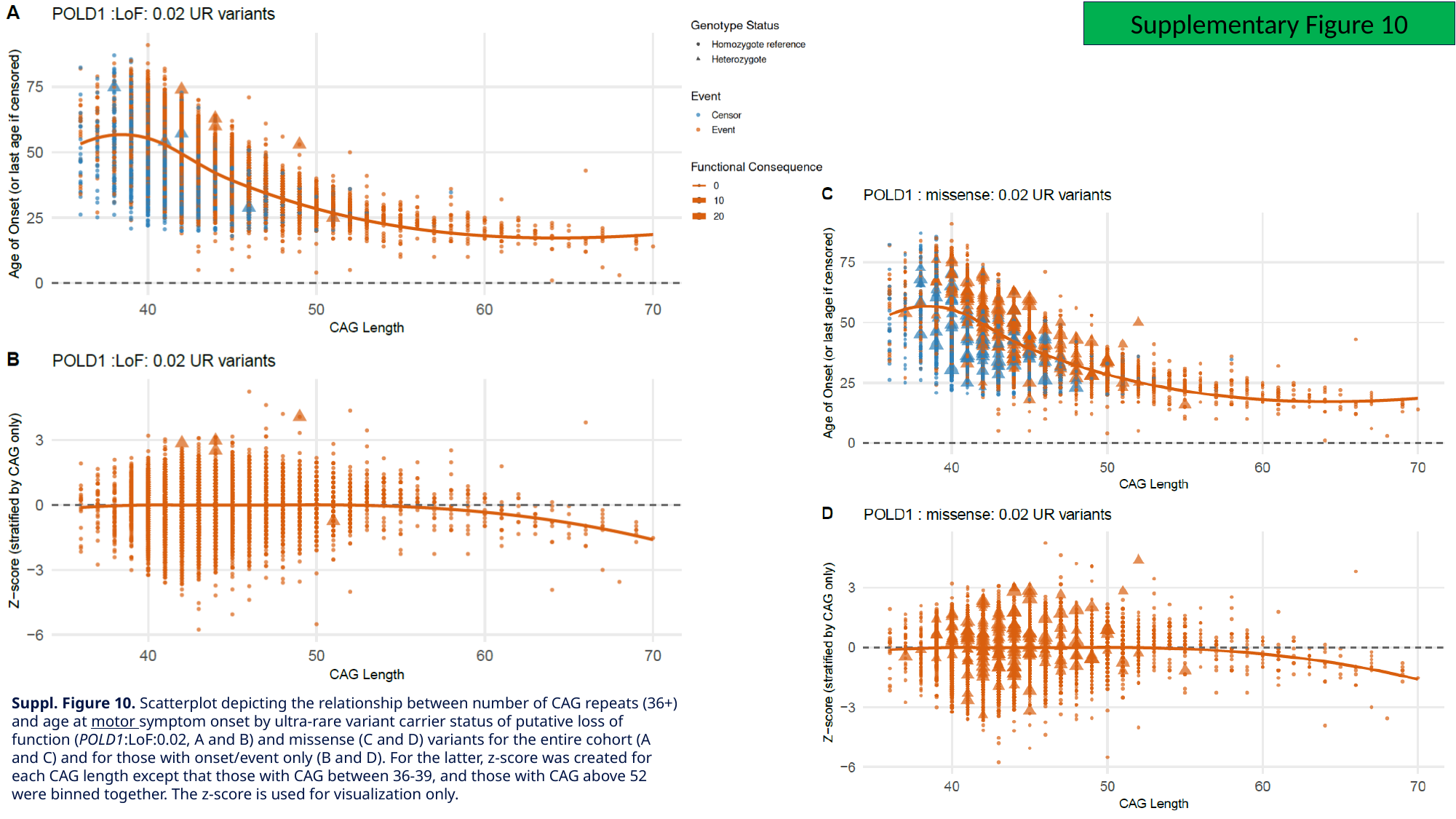

Supplementary Figure 10
Suppl. Figure 10. Scatterplot depicting the relationship between number of CAG repeats (36+) and age at motor symptom onset by ultra-rare variant carrier status of putative loss of function (POLD1:LoF:0.02, A and B) and missense (C and D) variants for the entire cohort (A and C) and for those with onset/event only (B and D). For the latter, z-score was created for each CAG length except that those with CAG between 36-39, and those with CAG above 52 were binned together. The z-score is used for visualization only.

### Slide 14
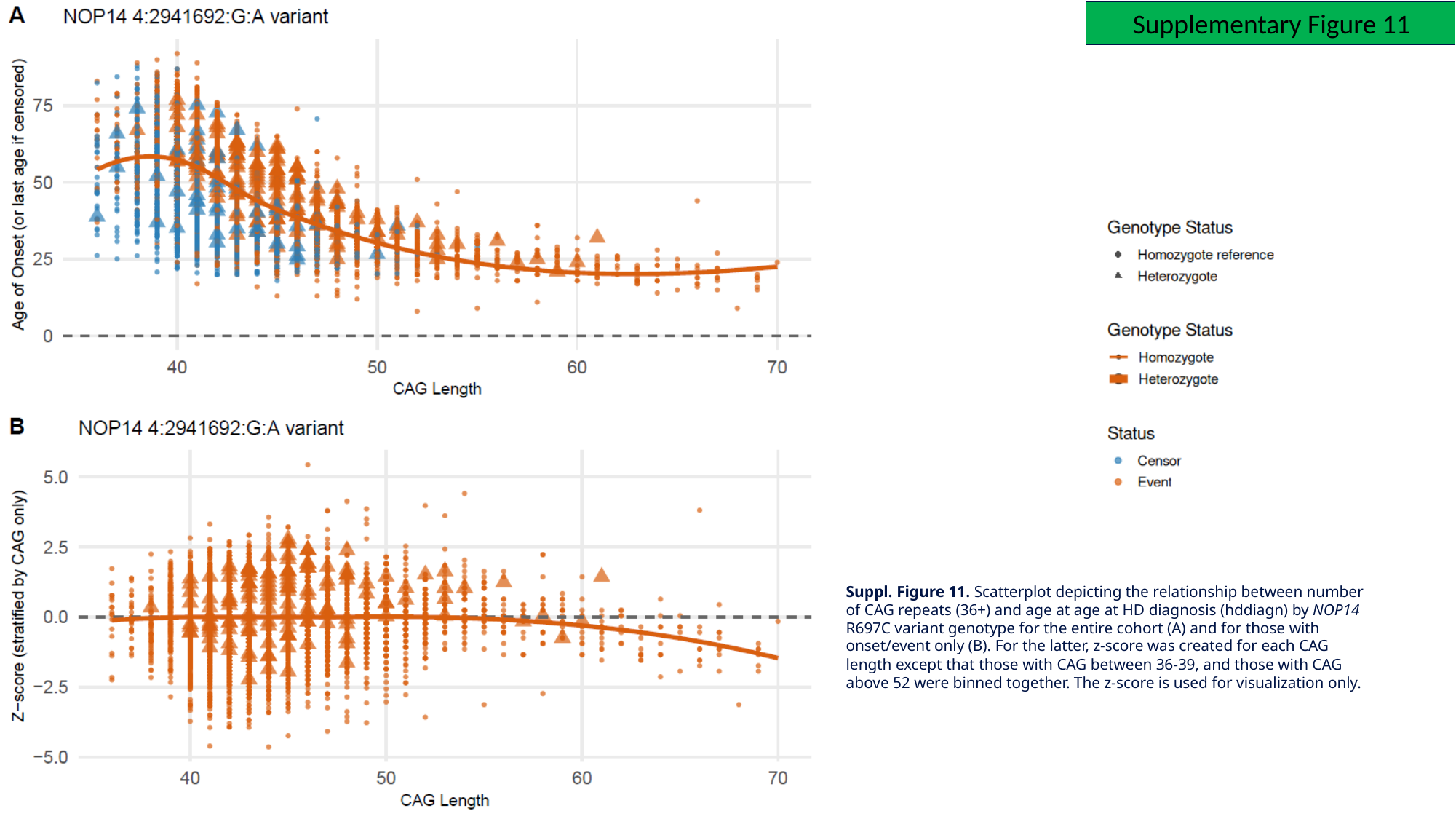

Supplementary Figure 11
Suppl. Figure 11. Scatterplot depicting the relationship between number of CAG repeats (36+) and age at age at HD diagnosis (hddiagn) by NOP14 R697C variant genotype for the entire cohort (A) and for those with onset/event only (B). For the latter, z-score was created for each CAG length except that those with CAG between 36-39, and those with CAG above 52 were binned together. The z-score is used for visualization only.

### Slide 15
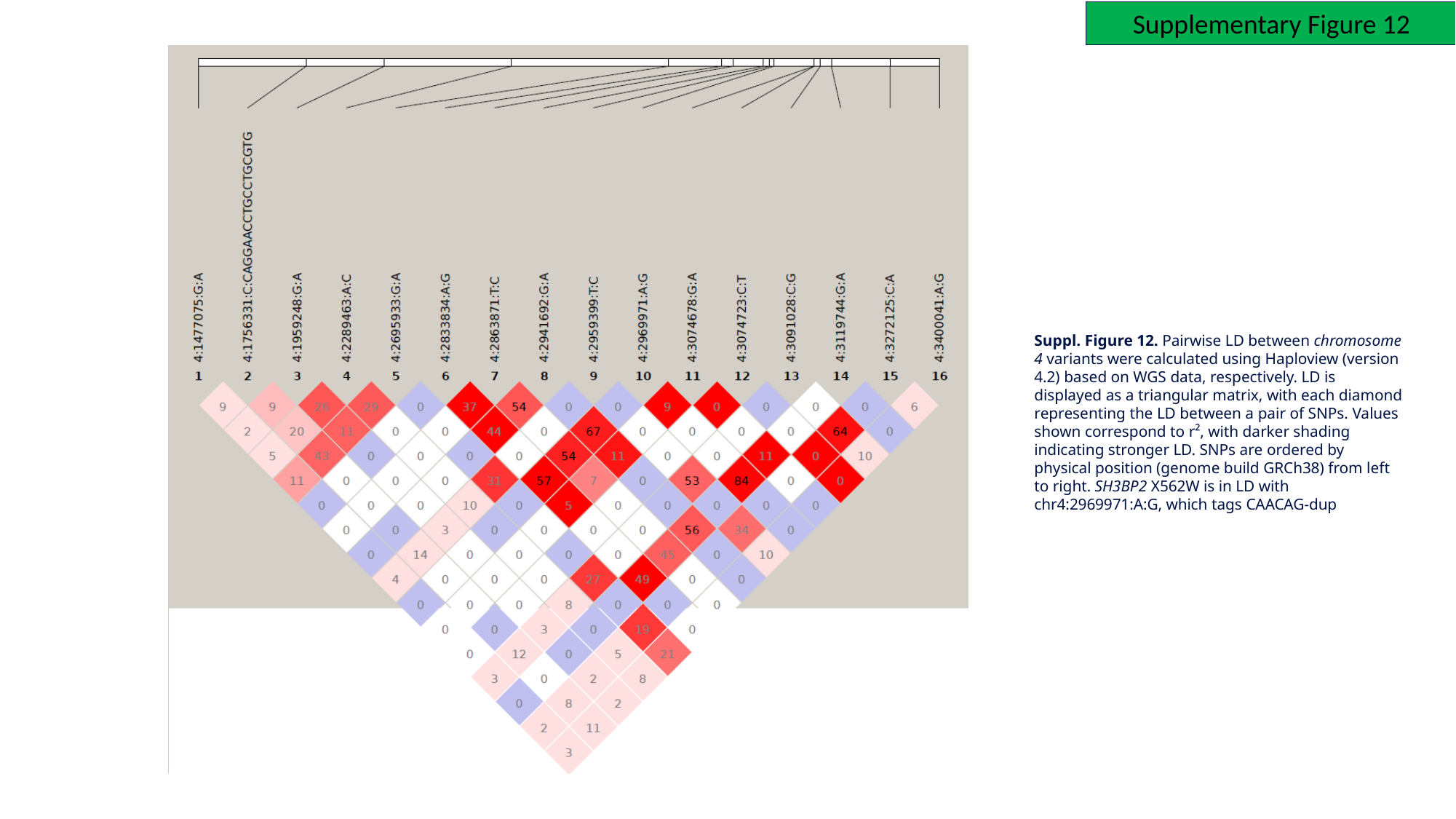

Supplementary Figure 12
Suppl. Figure 12. Pairwise LD between chromosome 4 variants were calculated using Haploview (version 4.2) based on WGS data, respectively. LD is displayed as a triangular matrix, with each diamond representing the LD between a pair of SNPs. Values shown correspond to r², with darker shading indicating stronger LD. SNPs are ordered by physical position (genome build GRCh38) from left to right. SH3BP2 X562W is in LD with chr4:2969971:A:G, which tags CAACAG-dup

### Slide 16
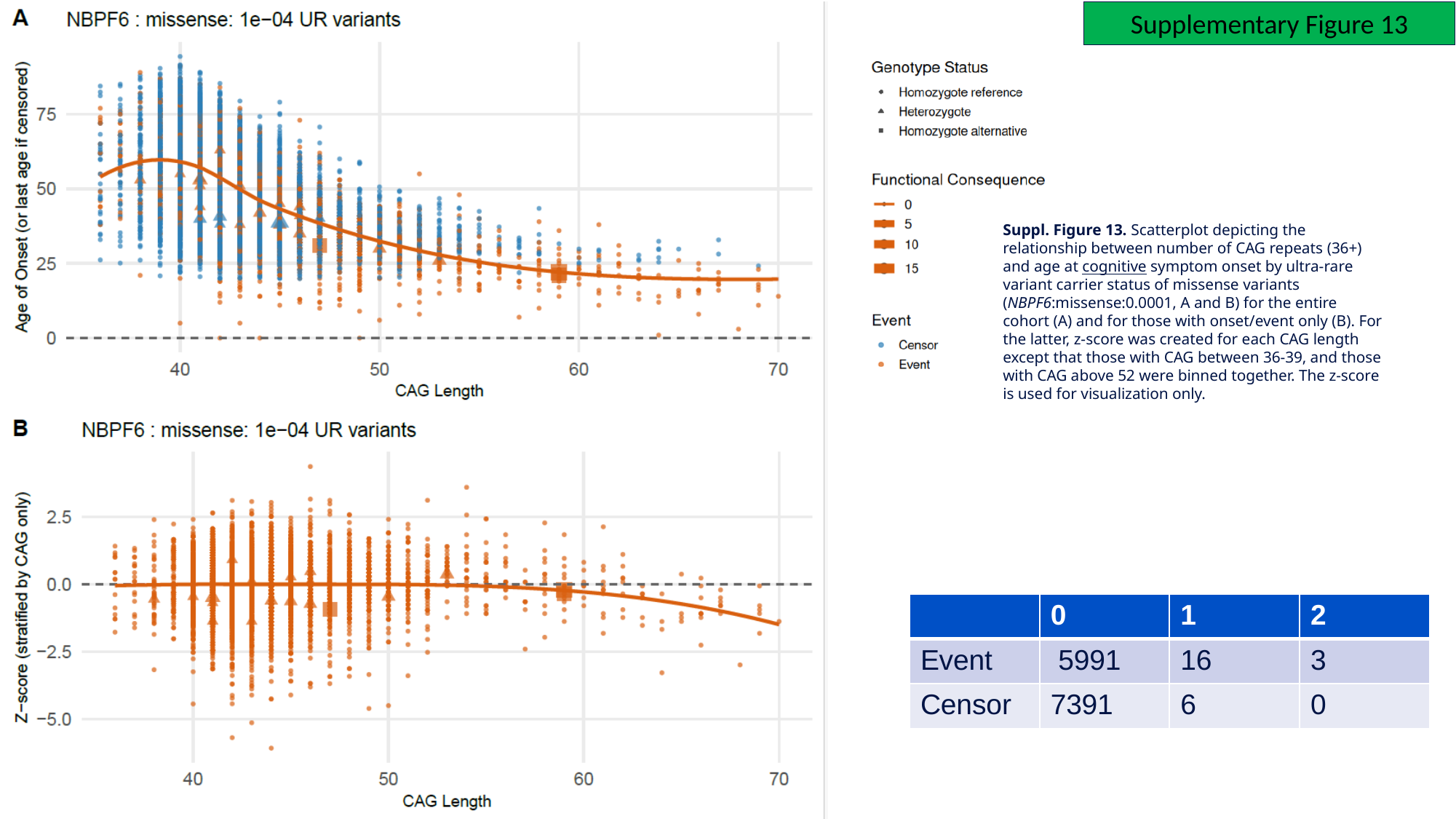

Supplementary Figure 13
Suppl. Figure 13. Scatterplot depicting the relationship between number of CAG repeats (36+) and age at cognitive symptom onset by ultra-rare variant carrier status of missense variants (NBPF6:missense:0.0001, A and B) for the entire cohort (A) and for those with onset/event only (B). For the latter, z-score was created for each CAG length except that those with CAG between 36-39, and those with CAG above 52 were binned together. The z-score is used for visualization only.
| | 0 | 1 | 2 |
| --- | --- | --- | --- |
| Event | 5991 | 16 | 3 |
| Censor | 7391 | 6 | 0 |

### Slide 17
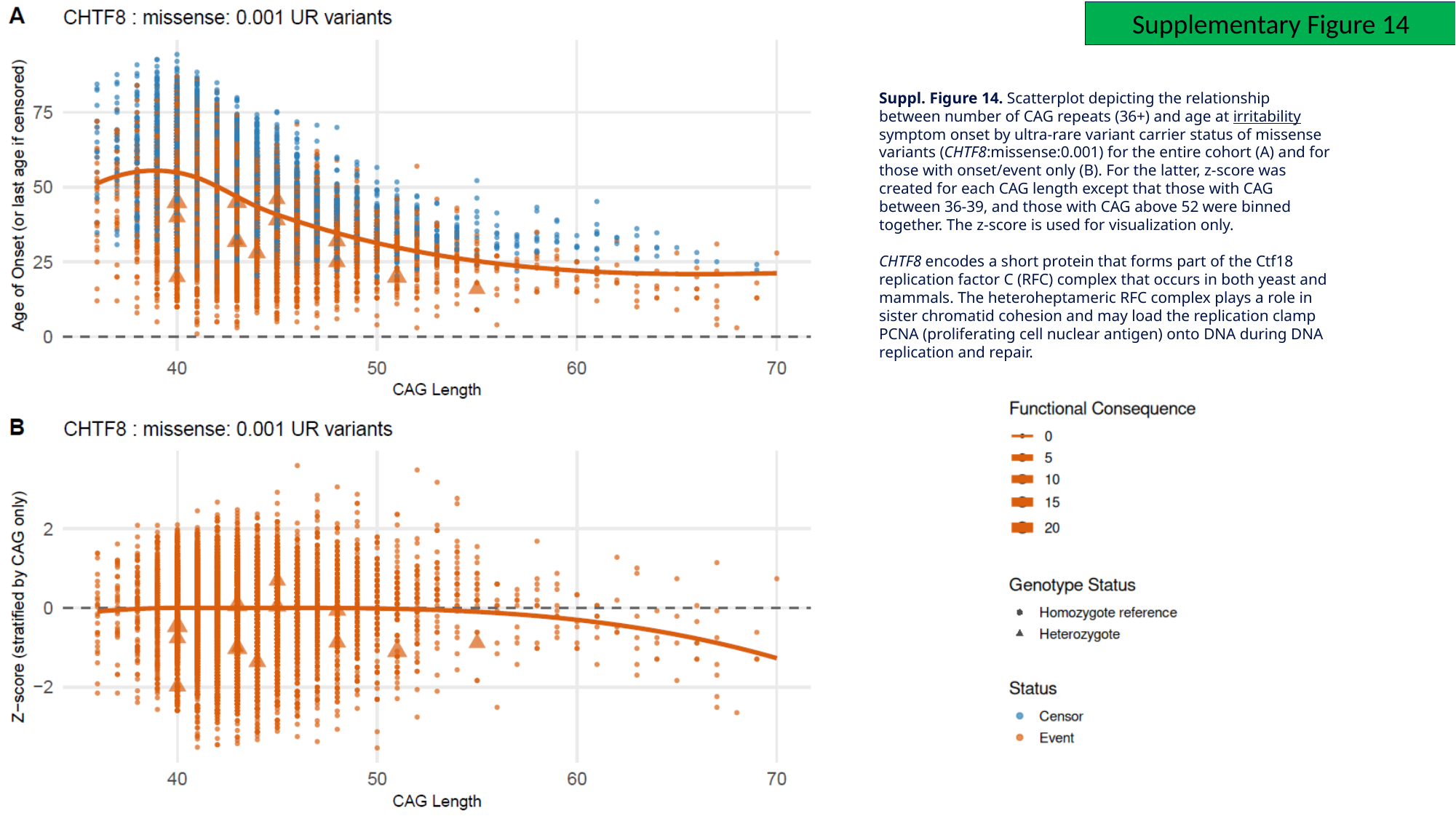

Supplementary Figure 14
Suppl. Figure 14. Scatterplot depicting the relationship between number of CAG repeats (36+) and age at irritability symptom onset by ultra-rare variant carrier status of missense variants (CHTF8:missense:0.001) for the entire cohort (A) and for those with onset/event only (B). For the latter, z-score was created for each CAG length except that those with CAG between 36-39, and those with CAG above 52 were binned together. The z-score is used for visualization only.
CHTF8 encodes a short protein that forms part of the Ctf18 replication factor C (RFC) complex that occurs in both yeast and mammals. The heteroheptameric RFC complex plays a role in sister chromatid cohesion and may load the replication clamp PCNA (proliferating cell nuclear antigen) onto DNA during DNA replication and repair.

### Slide 18
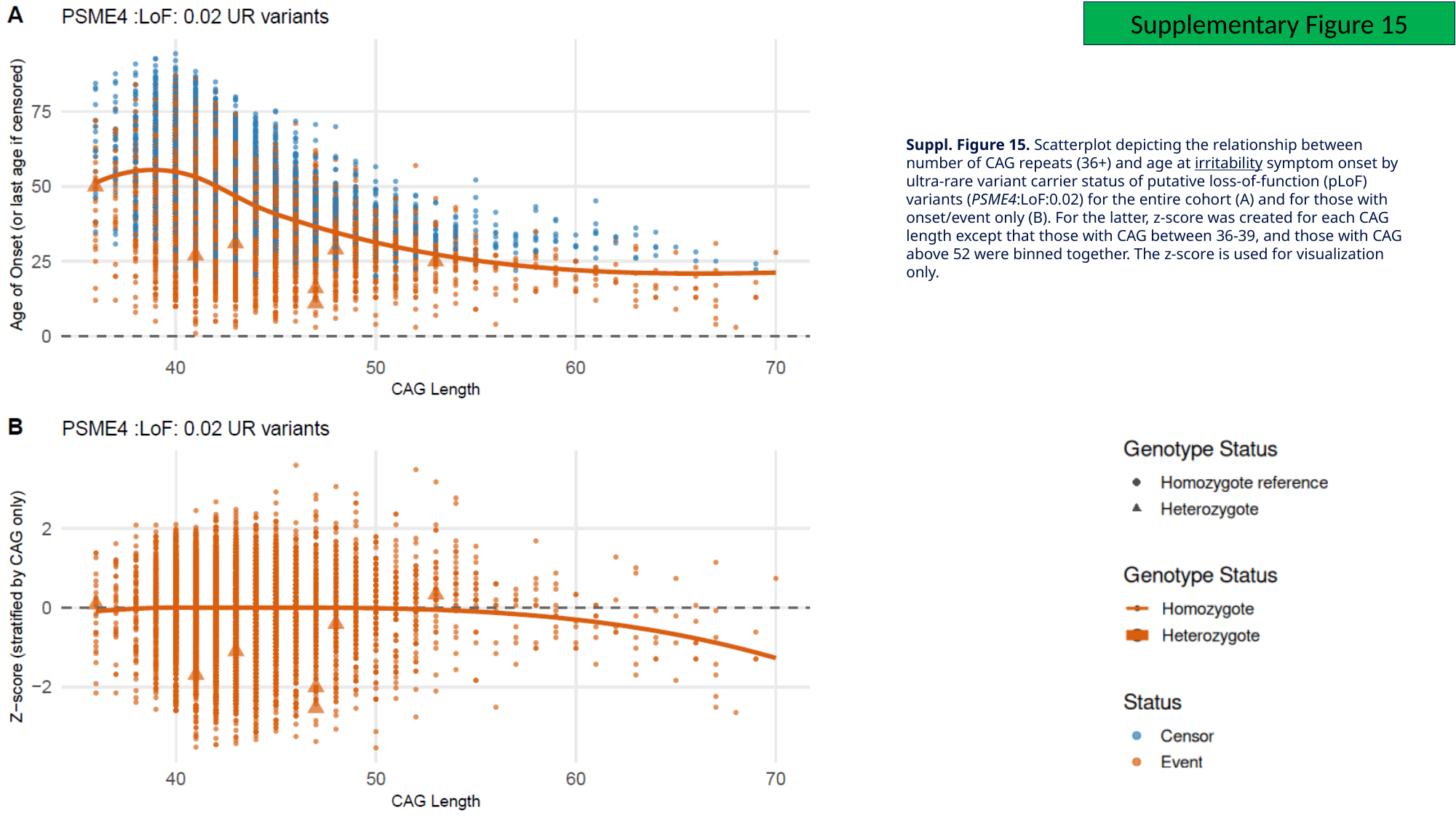

Supplementary Figure 15
Suppl. Figure 15. Scatterplot depicting the relationship between number of CAG repeats (36+) and age at irritability symptom onset by ultra-rare variant carrier status of putative loss-of-function (pLoF) variants (PSME4:LoF:0.02) for the entire cohort (A) and for those with onset/event only (B). For the latter, z-score was created for each CAG length except that those with CAG between 36-39, and those with CAG above 52 were binned together. The z-score is used for visualization only.

### Slide 19
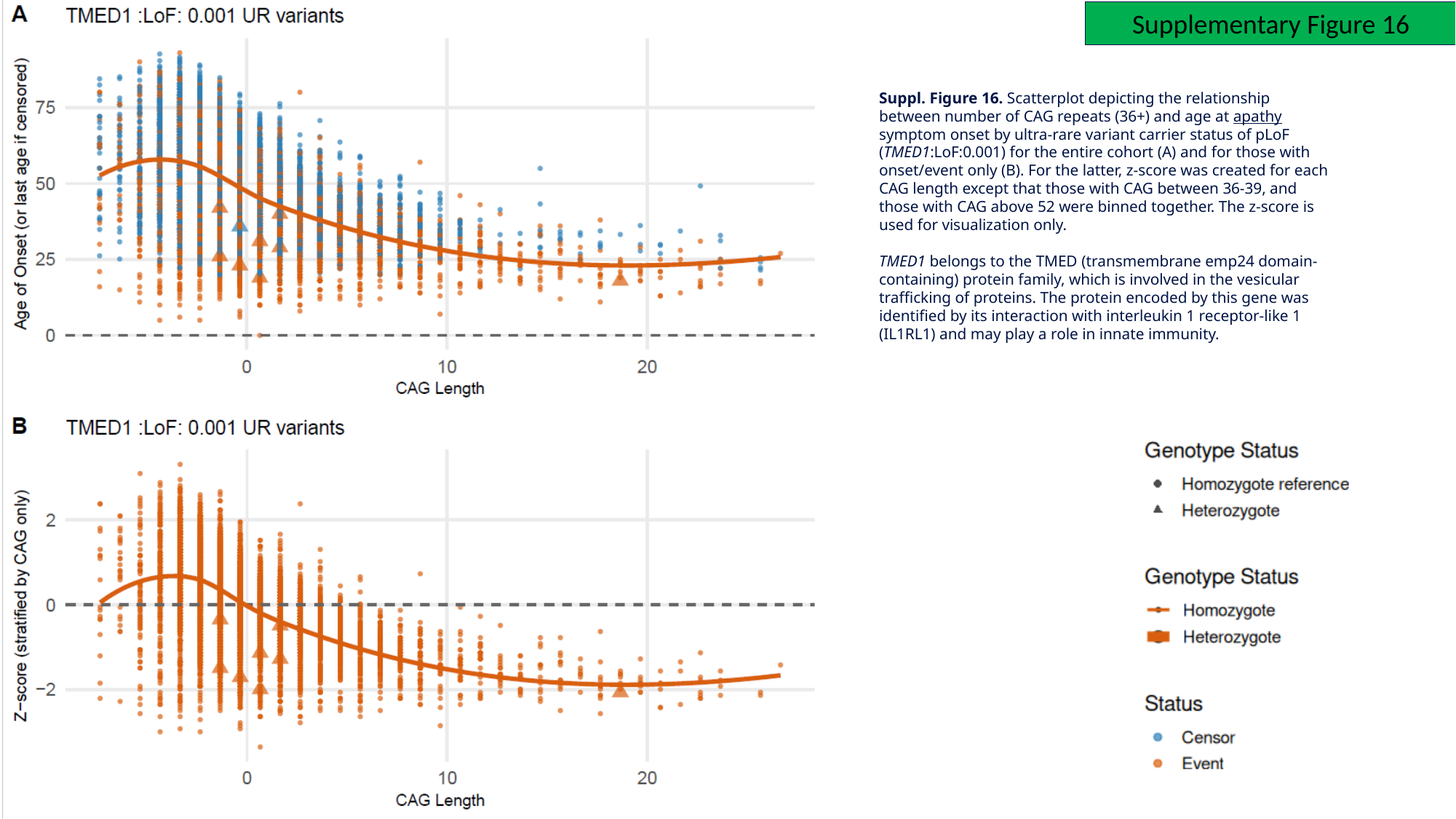

Supplementary Figure 16
Suppl. Figure 16. Scatterplot depicting the relationship between number of CAG repeats (36+) and age at apathy symptom onset by ultra-rare variant carrier status of pLoF (TMED1:LoF:0.001) for the entire cohort (A) and for those with onset/event only (B). For the latter, z-score was created for each CAG length except that those with CAG between 36-39, and those with CAG above 52 were binned together. The z-score is used for visualization only.
TMED1 belongs to the TMED (transmembrane emp24 domain-containing) protein family, which is involved in the vesicular trafficking of proteins. The protein encoded by this gene was identified by its interaction with interleukin 1 receptor-like 1 (IL1RL1) and may play a role in innate immunity.

### Slide 20
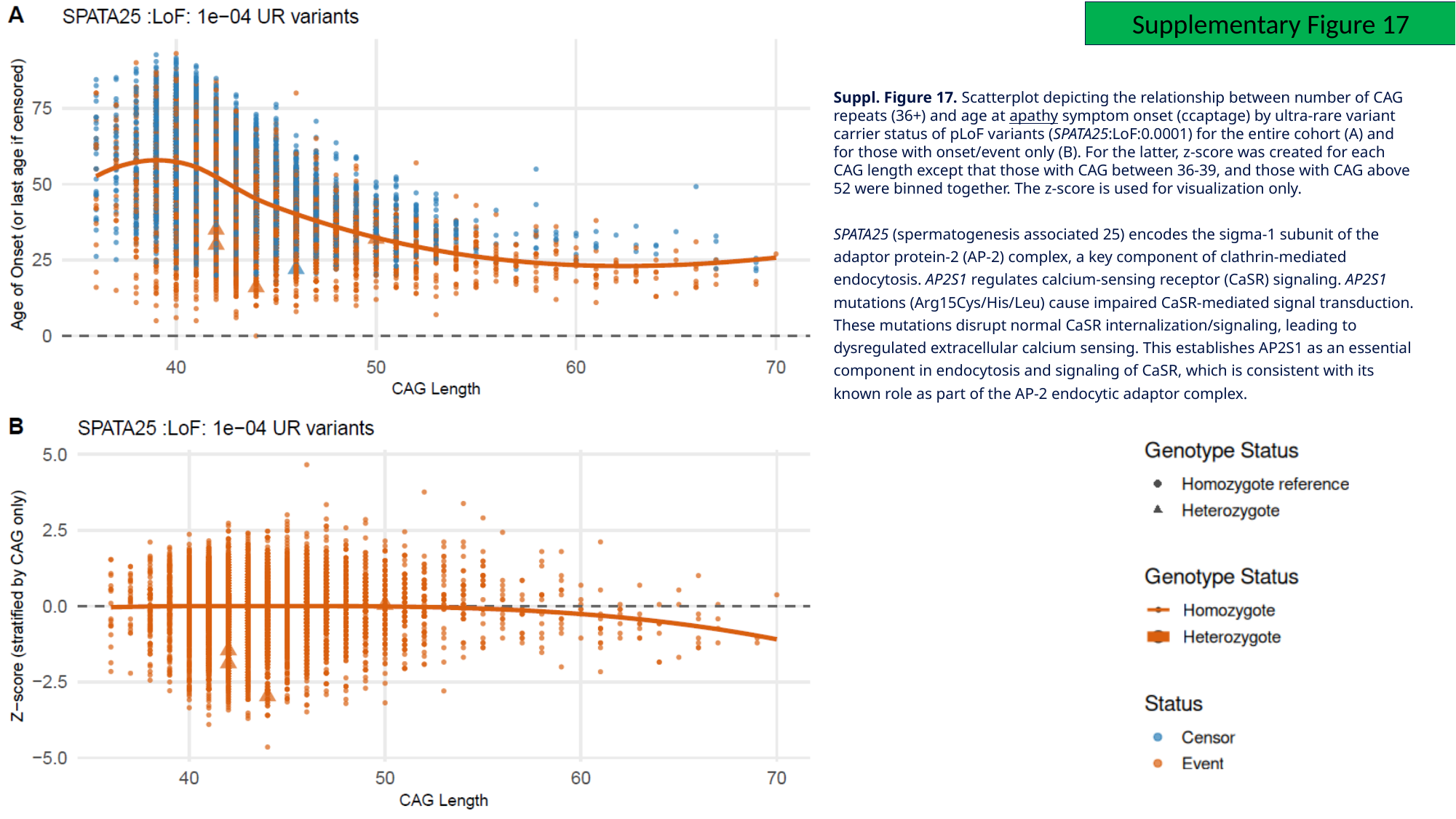

Supplementary Figure 17
Suppl. Figure 17. Scatterplot depicting the relationship between number of CAG repeats (36+) and age at apathy symptom onset (ccaptage) by ultra-rare variant carrier status of pLoF variants (SPATA25:LoF:0.0001) for the entire cohort (A) and for those with onset/event only (B). For the latter, z-score was created for each CAG length except that those with CAG between 36-39, and those with CAG above 52 were binned together. The z-score is used for visualization only.
SPATA25 (spermatogenesis associated 25) encodes the sigma‑1 subunit of the adaptor protein‑2 (AP‑2) complex, a key component of clathrin‑mediated endocytosis. AP2S1 regulates calcium‑sensing receptor (CaSR) signaling. AP2S1 mutations (Arg15Cys/His/Leu) cause impaired CaSR‑mediated signal transduction. These mutations disrupt normal CaSR internalization/signaling, leading to dysregulated extracellular calcium sensing. This establishes AP2S1 as an essential component in endocytosis and signaling of CaSR, which is consistent with its known role as part of the AP‑2 endocytic adaptor complex.

### Slide 21
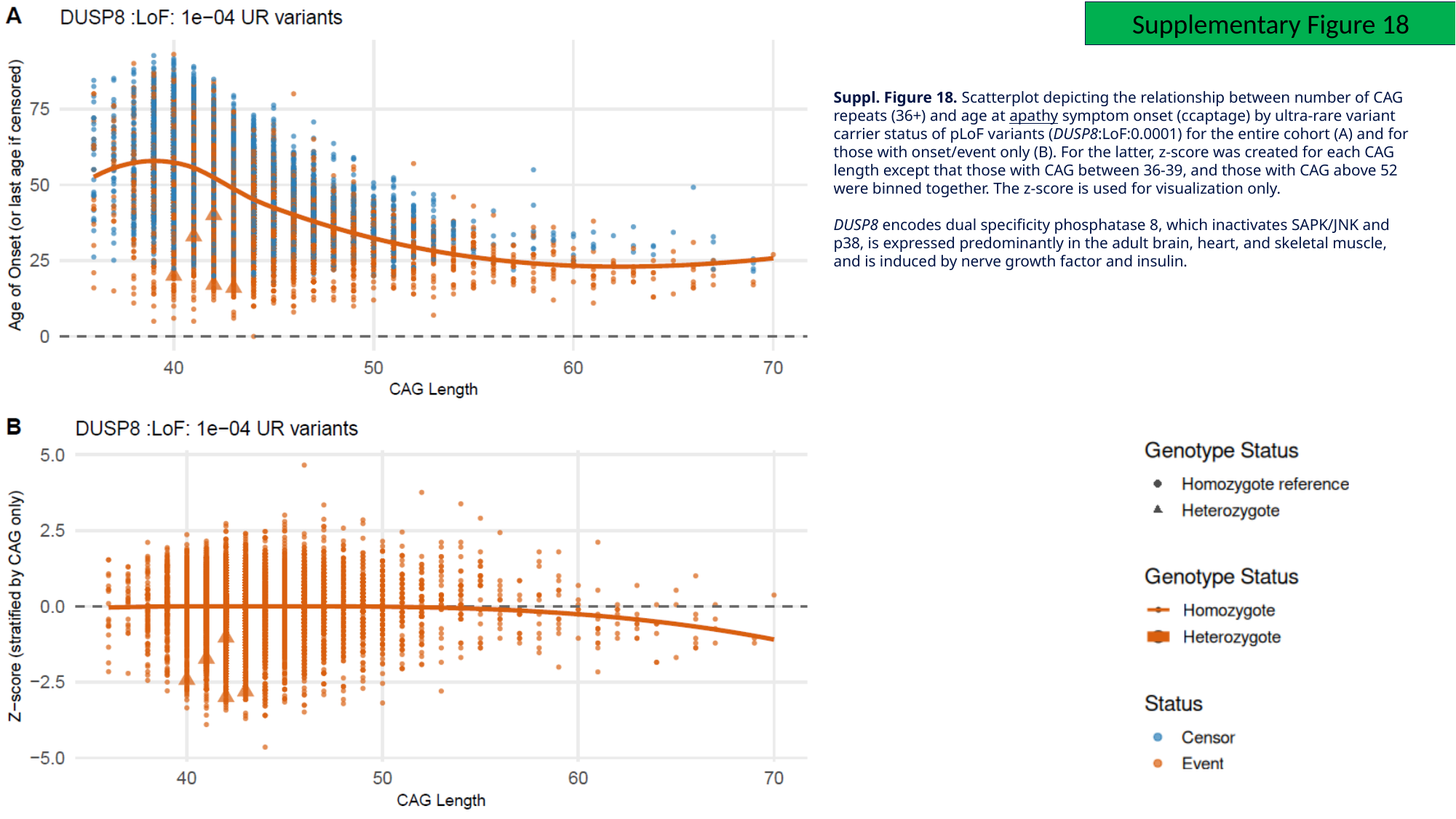

Supplementary Figure 18
Suppl. Figure 18. Scatterplot depicting the relationship between number of CAG repeats (36+) and age at apathy symptom onset (ccaptage) by ultra-rare variant carrier status of pLoF variants (DUSP8:LoF:0.0001) for the entire cohort (A) and for those with onset/event only (B). For the latter, z-score was created for each CAG length except that those with CAG between 36-39, and those with CAG above 52 were binned together. The z-score is used for visualization only.
DUSP8 encodes dual specificity phosphatase 8, which inactivates SAPK/JNK and p38, is expressed predominantly in the adult brain, heart, and skeletal muscle, and is induced by nerve growth factor and insulin.

### Slide 22
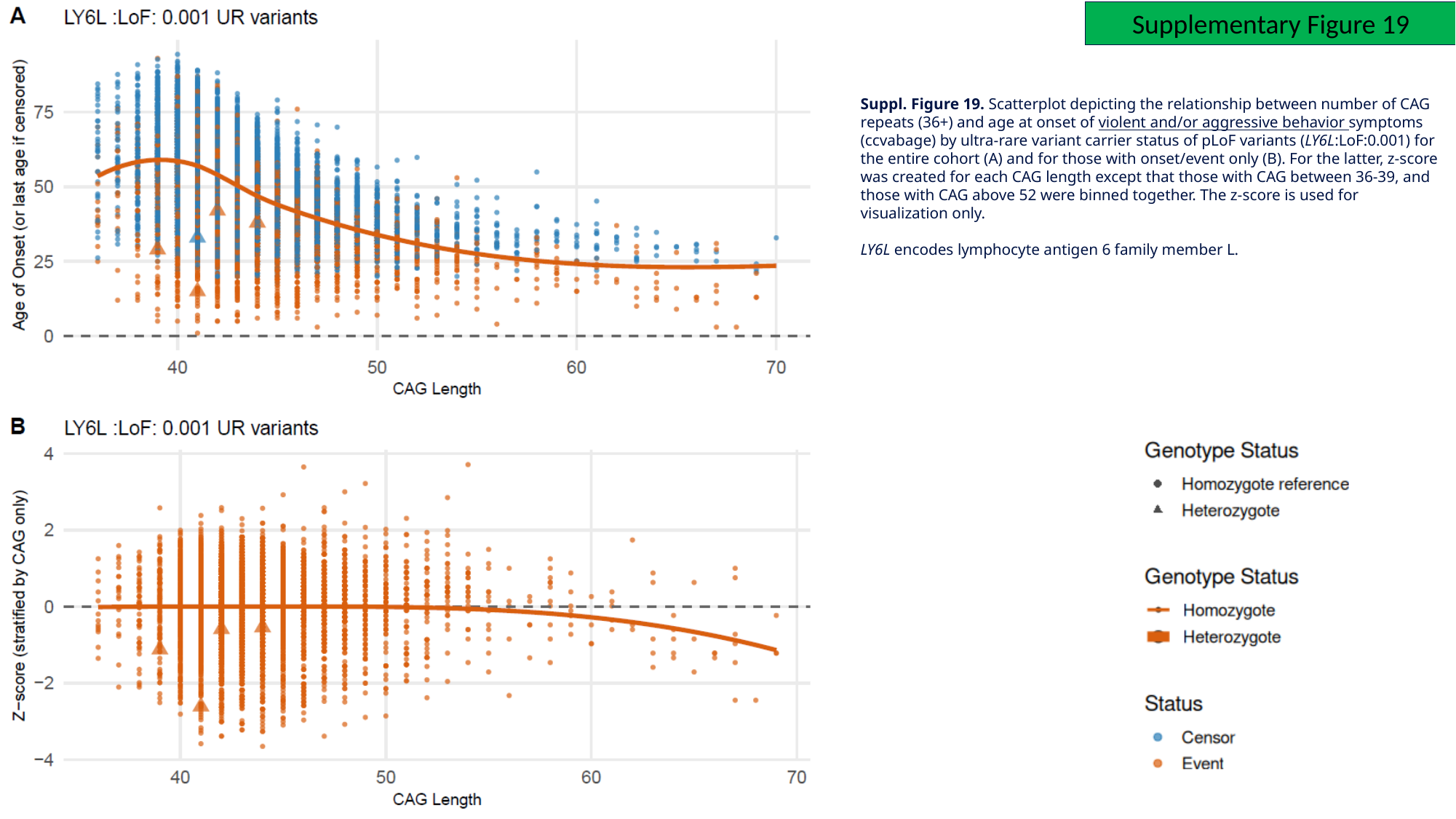

Supplementary Figure 19
Suppl. Figure 19. Scatterplot depicting the relationship between number of CAG repeats (36+) and age at onset of violent and/or aggressive behavior symptoms (ccvabage) by ultra-rare variant carrier status of pLoF variants (LY6L:LoF:0.001) for the entire cohort (A) and for those with onset/event only (B). For the latter, z-score was created for each CAG length except that those with CAG between 36-39, and those with CAG above 52 were binned together. The z-score is used for visualization only.
LY6L encodes lymphocyte antigen 6 family member L.

### Slide 23
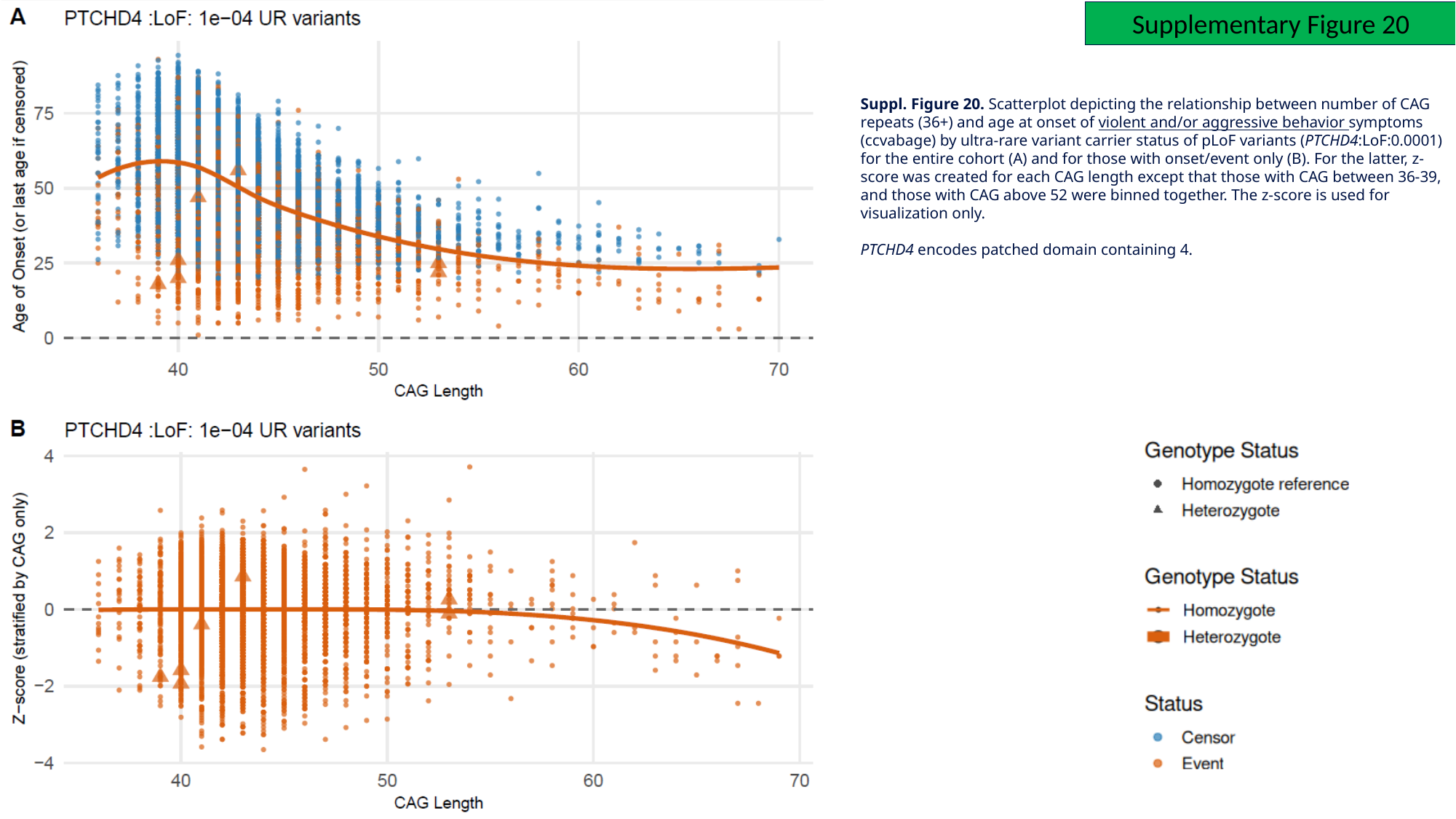

Supplementary Figure 20
Suppl. Figure 20. Scatterplot depicting the relationship between number of CAG repeats (36+) and age at onset of violent and/or aggressive behavior symptoms (ccvabage) by ultra-rare variant carrier status of pLoF variants (PTCHD4:LoF:0.0001) for the entire cohort (A) and for those with onset/event only (B). For the latter, z-score was created for each CAG length except that those with CAG between 36-39, and those with CAG above 52 were binned together. The z-score is used for visualization only.
PTCHD4 encodes patched domain containing 4.

### Slide 24
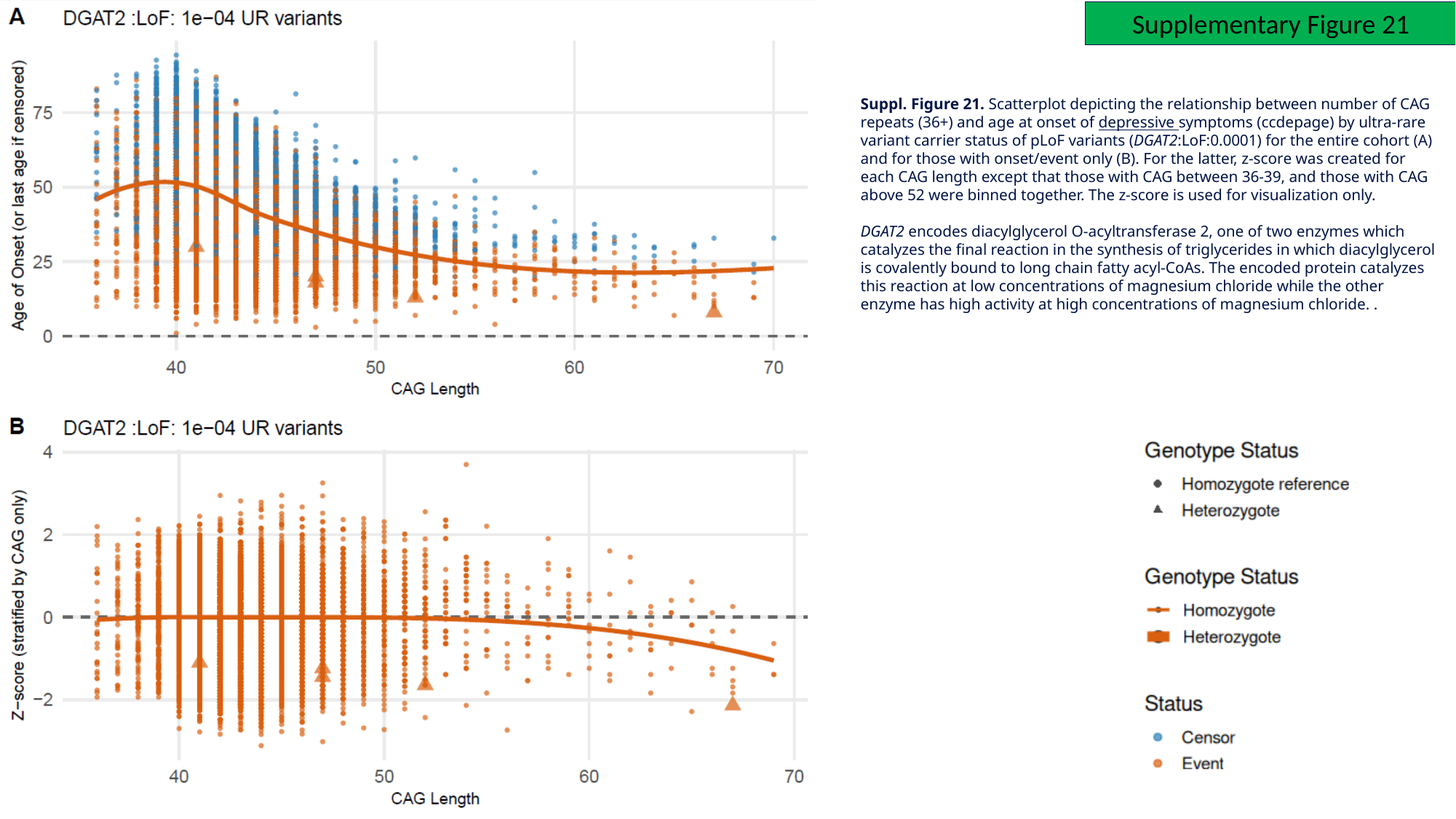

Supplementary Figure 21
Suppl. Figure 21. Scatterplot depicting the relationship between number of CAG repeats (36+) and age at onset of depressive symptoms (ccdepage) by ultra-rare variant carrier status of pLoF variants (DGAT2:LoF:0.0001) for the entire cohort (A) and for those with onset/event only (B). For the latter, z-score was created for each CAG length except that those with CAG between 36-39, and those with CAG above 52 were binned together. The z-score is used for visualization only.
DGAT2 encodes diacylglycerol O-acyltransferase 2, one of two enzymes which catalyzes the final reaction in the synthesis of triglycerides in which diacylglycerol is covalently bound to long chain fatty acyl-CoAs. The encoded protein catalyzes this reaction at low concentrations of magnesium chloride while the other enzyme has high activity at high concentrations of magnesium chloride. .

### Slide 25
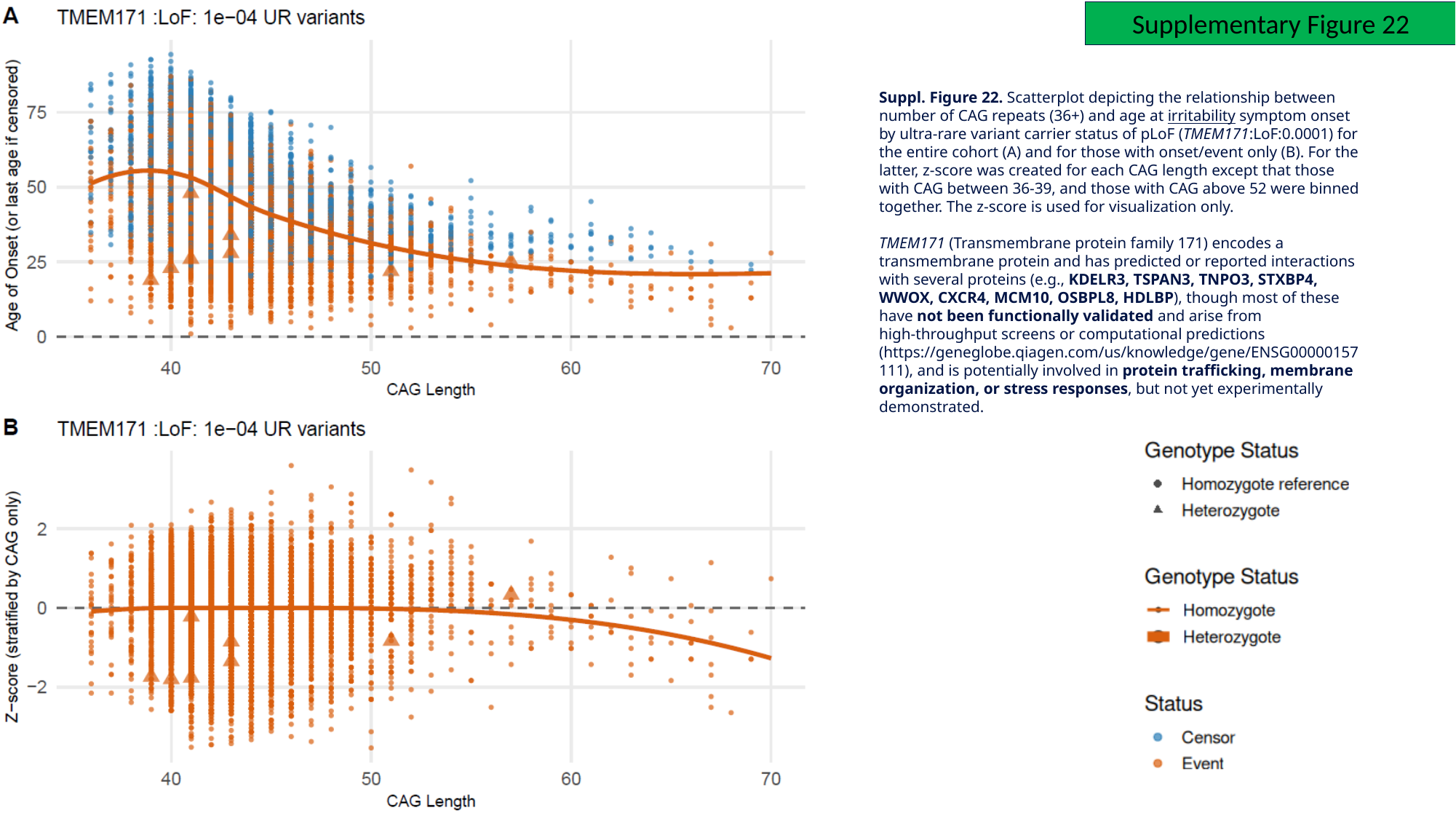

Supplementary Figure 22
Suppl. Figure 22. Scatterplot depicting the relationship between number of CAG repeats (36+) and age at irritability symptom onset by ultra-rare variant carrier status of pLoF (TMEM171:LoF:0.0001) for the entire cohort (A) and for those with onset/event only (B). For the latter, z-score was created for each CAG length except that those with CAG between 36-39, and those with CAG above 52 were binned together. The z-score is used for visualization only.
TMEM171 (Transmembrane protein family 171) encodes a transmembrane protein and has predicted or reported interactions with several proteins (e.g., KDELR3, TSPAN3, TNPO3, STXBP4, WWOX, CXCR4, MCM10, OSBPL8, HDLBP), though most of these have not been functionally validated and arise from high‑throughput screens or computational predictions (https://geneglobe.qiagen.com/us/knowledge/gene/ENSG00000157111), and is potentially involved in protein trafficking, membrane organization, or stress responses, but not yet experimentally demonstrated.

### Slide 26
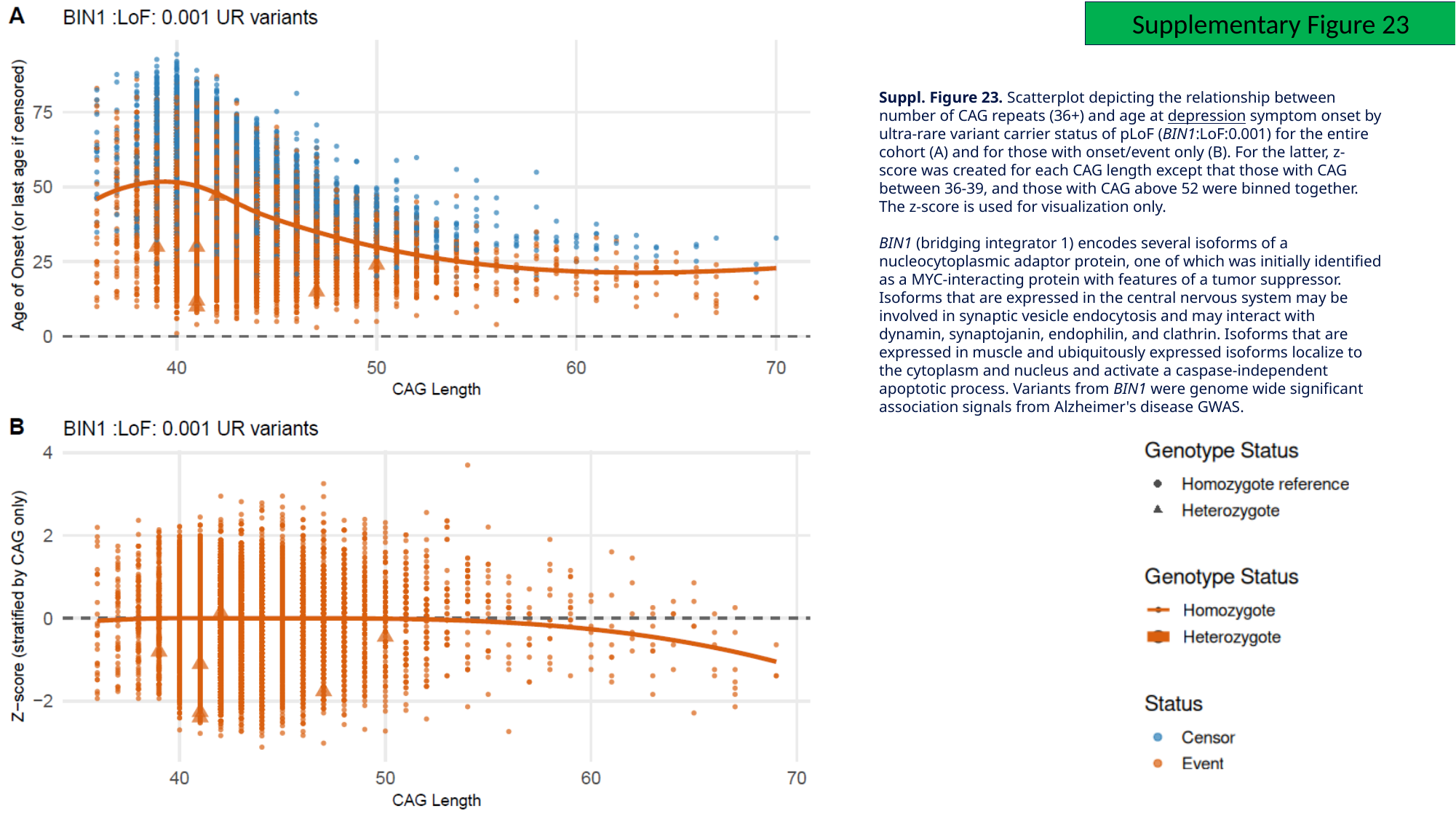

Supplementary Figure 23
Suppl. Figure 23. Scatterplot depicting the relationship between number of CAG repeats (36+) and age at depression symptom onset by ultra-rare variant carrier status of pLoF (BIN1:LoF:0.001) for the entire cohort (A) and for those with onset/event only (B). For the latter, z-score was created for each CAG length except that those with CAG between 36-39, and those with CAG above 52 were binned together. The z-score is used for visualization only.
BIN1 (bridging integrator 1) encodes several isoforms of a nucleocytoplasmic adaptor protein, one of which was initially identified as a MYC-interacting protein with features of a tumor suppressor. Isoforms that are expressed in the central nervous system may be involved in synaptic vesicle endocytosis and may interact with dynamin, synaptojanin, endophilin, and clathrin. Isoforms that are expressed in muscle and ubiquitously expressed isoforms localize to the cytoplasm and nucleus and activate a caspase-independent apoptotic process. Variants from BIN1 were genome wide significant association signals from Alzheimer's disease GWAS.

### Slide 27
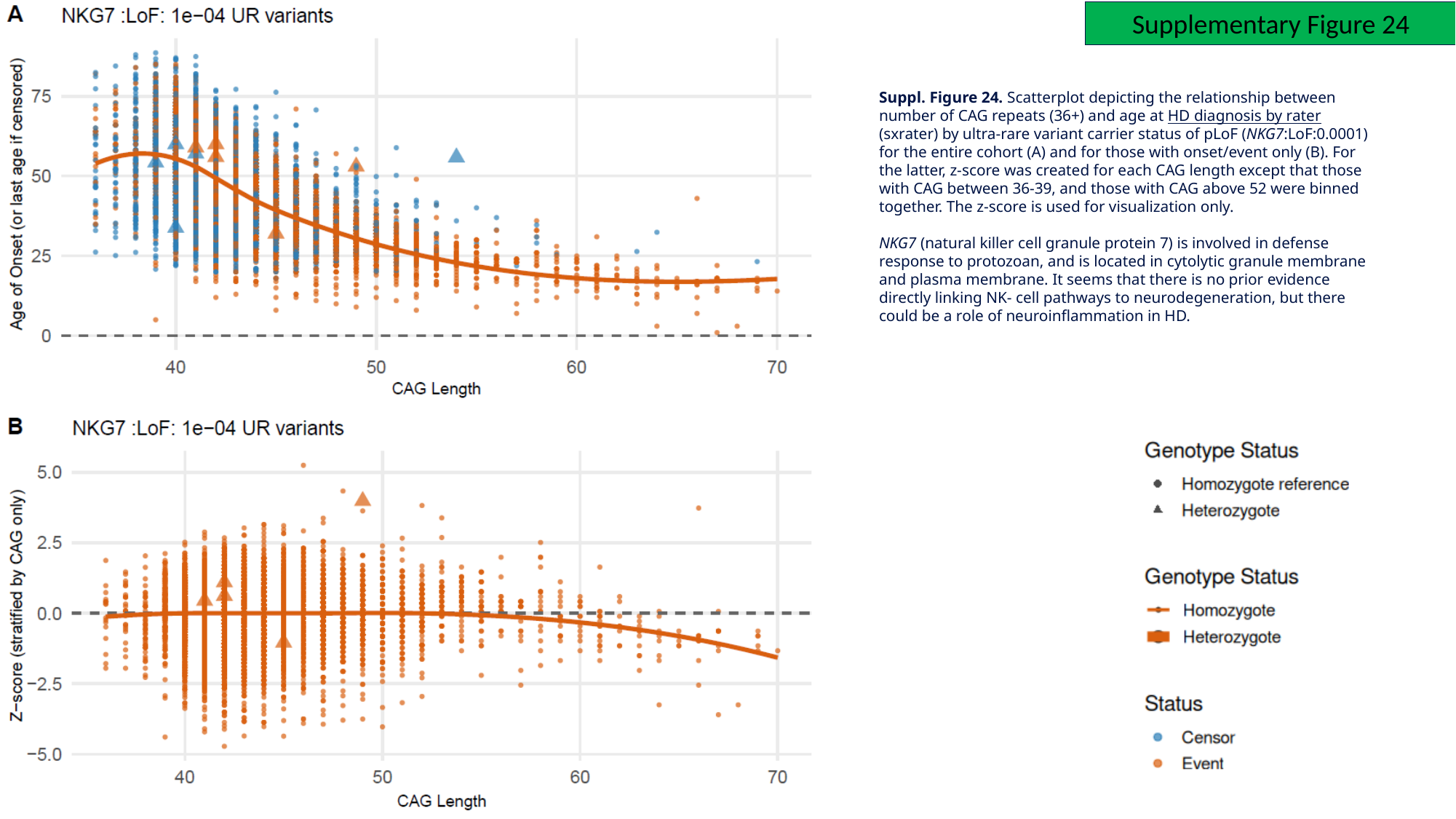

Supplementary Figure 24
Suppl. Figure 24. Scatterplot depicting the relationship between number of CAG repeats (36+) and age at HD diagnosis by rater (sxrater) by ultra-rare variant carrier status of pLoF (NKG7:LoF:0.0001) for the entire cohort (A) and for those with onset/event only (B). For the latter, z-score was created for each CAG length except that those with CAG between 36-39, and those with CAG above 52 were binned together. The z-score is used for visualization only.
NKG7 (natural killer cell granule protein 7) is involved in defense response to protozoan, and is located in cytolytic granule membrane and plasma membrane. It seems that there is no prior evidence directly linking NK- cell pathways to neurodegeneration, but there could be a role of neuroinflammation in HD.

### Slide 28
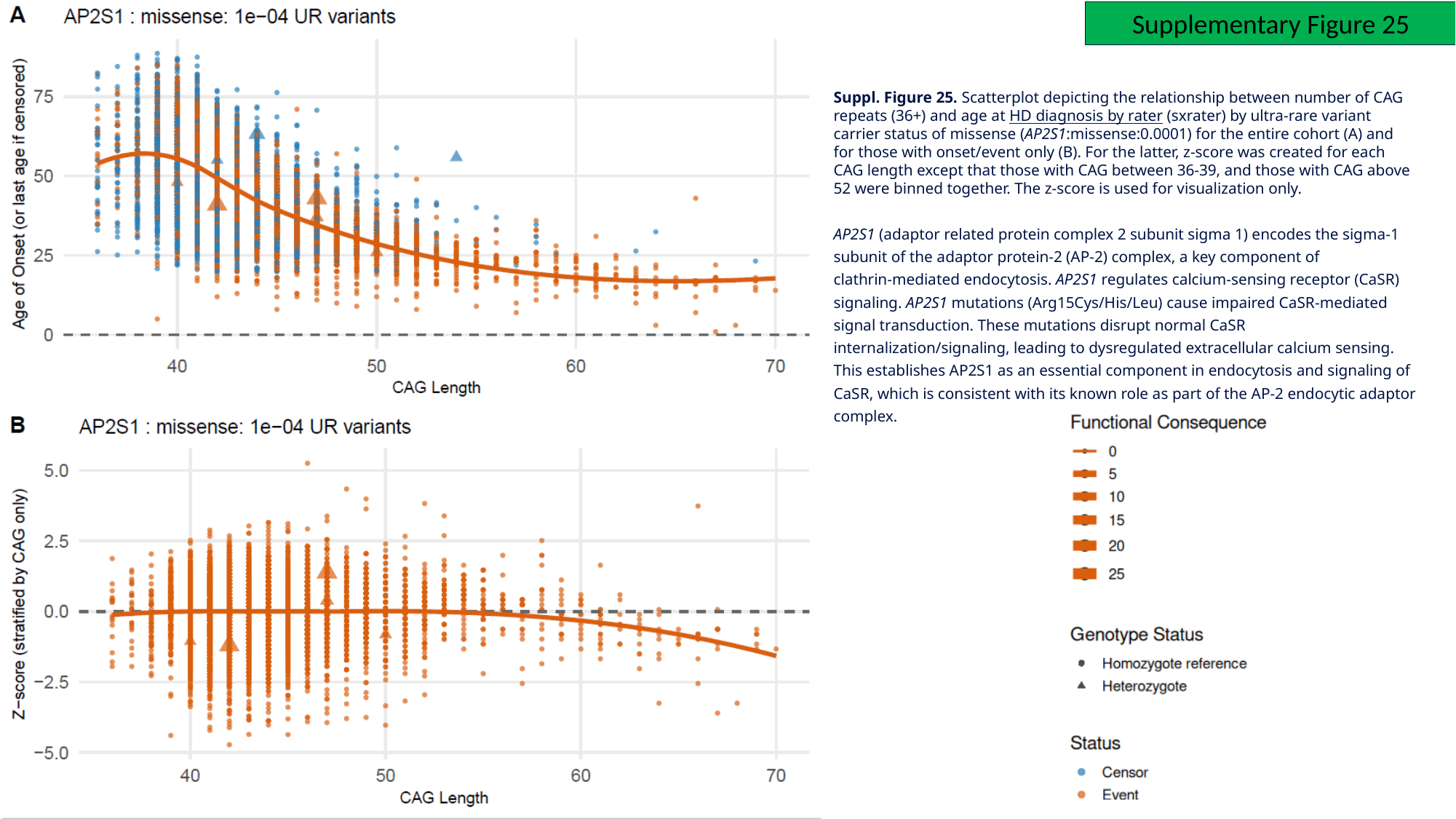

Supplementary Figure 25
Suppl. Figure 25. Scatterplot depicting the relationship between number of CAG repeats (36+) and age at HD diagnosis by rater (sxrater) by ultra-rare variant carrier status of missense (AP2S1:missense:0.0001) for the entire cohort (A) and for those with onset/event only (B). For the latter, z-score was created for each CAG length except that those with CAG between 36-39, and those with CAG above 52 were binned together. The z-score is used for visualization only.
AP2S1 (adaptor related protein complex 2 subunit sigma 1) encodes the sigma‑1 subunit of the adaptor protein‑2 (AP‑2) complex, a key component of clathrin‑mediated endocytosis. AP2S1 regulates calcium‑sensing receptor (CaSR) signaling. AP2S1 mutations (Arg15Cys/His/Leu) cause impaired CaSR‑mediated signal transduction. These mutations disrupt normal CaSR internalization/signaling, leading to dysregulated extracellular calcium sensing. This establishes AP2S1 as an essential component in endocytosis and signaling of CaSR, which is consistent with its known role as part of the AP‑2 endocytic adaptor complex.

### Slide 29
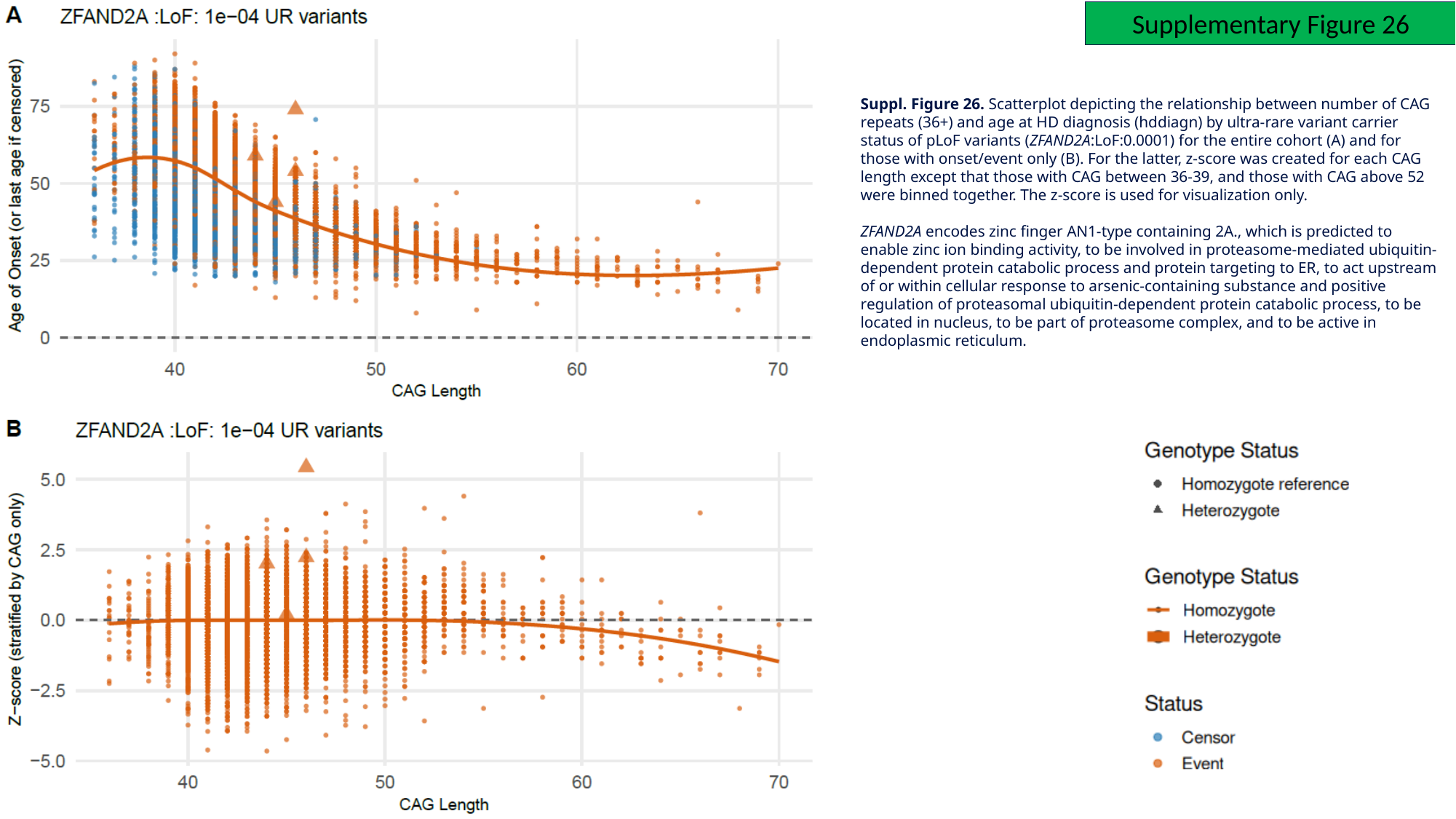

Supplementary Figure 26
Suppl. Figure 26. Scatterplot depicting the relationship between number of CAG repeats (36+) and age at HD diagnosis (hddiagn) by ultra-rare variant carrier status of pLoF variants (ZFAND2A:LoF:0.0001) for the entire cohort (A) and for those with onset/event only (B). For the latter, z-score was created for each CAG length except that those with CAG between 36-39, and those with CAG above 52 were binned together. The z-score is used for visualization only.
ZFAND2A encodes zinc finger AN1-type containing 2A., which is predicted to enable zinc ion binding activity, to be involved in proteasome-mediated ubiquitin-dependent protein catabolic process and protein targeting to ER, to act upstream of or within cellular response to arsenic-containing substance and positive regulation of proteasomal ubiquitin-dependent protein catabolic process, to be located in nucleus, to be part of proteasome complex, and to be active in endoplasmic reticulum.

### Slide 30
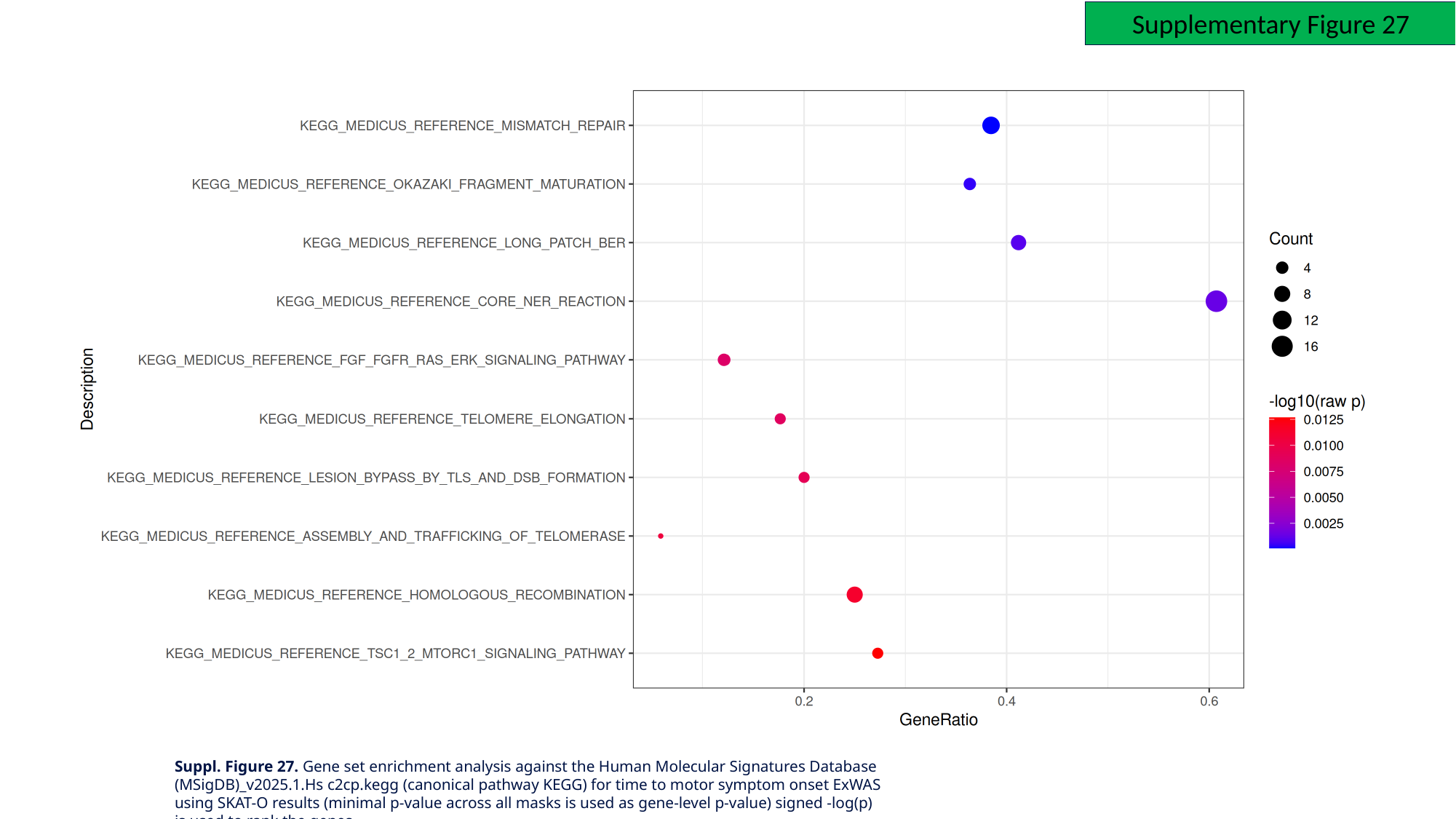

Supplementary Figure 27
Suppl. Figure 27. Gene set enrichment analysis against the Human Molecular Signatures Database (MSigDB)_v2025.1.Hs c2cp.kegg (canonical pathway KEGG) for time to motor symptom onset ExWAS using SKAT-O results (minimal p-value across all masks is used as gene-level p-value) signed -log(p) is used to rank the genes
